# Medical and non-medical out-of-pocket cost and time burden of COVID-19 illness in U.S. children and adults: a cross-sectional survey study

**DOI:** 10.64898/2026.10.14.26365534

**Authors:** Angela M. Rose, Aaron M. Scherer, Amber M. Gedlinske, Andrew M. Parker, Courtney A. Gidengil, Natoshia M. Askelson, Megan C. Lindley, Acham Gebremariam, Jamison Pike, Lisa A. Prosser

## Abstract

**Background:** While health system costs of COVID-19 are well documented, evidence on out-of-pocket costs to patients and families, including non-medical and time burden, is limited. This study aimed to estimate out-of-pocket medical, non-medical, and time burden of COVID-19 illness directly from patients and caregivers.

**Methods:** Two cross-sectional surveys were fielded to national nonprobability samples of U.S. adults in Fall 2023: a personal burden survey (adult sample) and a caregiver burden survey (child sample). Surveys collected data on care setting, visit duration including travel and wait time, and medical and non-medical out-of-pocket costs. After applying quality-based exclusion criteria, 409 adult sample and 52 child sample responses were analyzed.

**Results:** Over 87% of respondents were insured. Median out-of-pocket medical costs per visit were low, ranging from $5 (emergency department) to $10 (urgent care) in the adult sample, and from $0 (emergency department) to $75 (hospitalization) in the child sample. Median non-medical costs were $28 or less across care settings. Median time spent receiving care ranged from 90 minutes (primary care, child sample) to 300 minutes (emergency department, adult sample). Despite the relatively low reported out-of-pocket costs, COVID-related financial challenges were reported by 34% of the adult sample and 29% of the child sample, with the most common difficulty being paying non-medical bills (11% and 15%, respectively). Twelve percent in both groups reported problems paying COVID-19-related medical bills.

**Conclusions:** In this post-pandemic non-probability sample, individual out-of-pocket costs for COVID-19 illness were relatively modest. However, time burden was substantial and one-third of respondents reported financial hardship. These estimates, including non-medical and time burden not captured in administrative data, can support more robust economic evaluations of COVID-19 prevention and treatment interventions.

## Background

Despite annual decreases in COVID-19 hospitalization rates, COVID-19 remains a leading cause of respiratory virus-associated infection and hospitalization [1, 2]. There is extensive literature describing COVID-19 related healthcare utilization and associated costs to the health system [3-7], however limited evidence is available on the out-of-pocket costs of COVID-19 illness to families.

Healthcare costs are increasingly being shifted from insurers to consumers, via increased deductibles, co-pays, and co-insurance [8]. Among those with insurance, 9% of US families have reported problems paying medical bills in the past 12 months [9]. Therefore, measuring additional costs that are accrued outside of the health system are critical to estimating the full economic impact of an illness on families.

The purpose of this study was to provide estimates of out-of-pocket medical, non-medical, and time burden associated with COVID-19 illness. Previous estimates of out-of-pocket costs for COVID-19 illness [10-12] used administrative data to report medical co-pays, while this study collected data on costs directly from patients and their families. This study also includes costs not available from the administrative record such as time associated with receiving care or treatment and non-medical out-of-pocket expenses such as transportation, parking, and meals away from home, providing a broader estimate of the total family burden of illness. Estimates on these costs incurred at the family level can be used to conduct more robust economic analyses of COVID-19 prevention and treatment interventions.

## Methods

### Survey development

The two surveys were developed by the Healthcare and Public Perceptions of Immunizations (HaPPI) survey collaborative – a cooperative agreement between researchers at the Centers for Disease Control and Prevention (CDC), researchers at the University of Iowa, and RAND. Initial survey drafts were developed by adapting survey items from previous surveys assessing out-of-pocket costs for respiratory syncytial virus (RSV) illnesses, with additional items adapted from previous HaPPI surveys [13]. Survey drafts were reviewed by CDC subject matter experts for feedback and revised in response. One survey focused on the personal burden of the respondent’s most recent COVID-19 illness and the second focused on burden of caregiving for someone with COVID-19.

The surveys asked about the respondent’s or their family member’s/friend’s most recent COVID-19 infection. Respondents rated the severity of their COVID-19 illness (and Long COVID severity, if applicable) on a 6-point scale, with “No symptoms (1)” and “Very severe (6)” as scale anchors. Both surveys included detailed questions about the settings where medical care was received (e.g. primary care clinic, urgent care clinic, etc.), how long the medical visits took including travel and wait time, and the medical and non-medical (e.g. gas, parking, and meals) costs associated with receiving care for their first visit of that type. The survey also included questions about financial challenges more broadly associated with the most recent COVID-19 illness. Finally, the survey collected demographic information including COVID-19 vaccination status. A list of the topical sections in the survey and types of questions asked is available in Additional file 1.

The personal burden survey was fielded from September 8 to 17, 2023 and the caregiver burden survey was fielded from October 18 to 23, 2023, to national, nonprobability samples of U.S. adults recruited through a survey panel managed by Dynata [14].

Sampling quotas were used for respondent sex (1:1 female to male ratio) and the two largest racial/ethnic minority groups in the United States (20% minimum for Hispanic respondents, 15% minimum for non-Hispanic Black respondents) and were tracked via responses to survey items. Inclusion criteria included respondents (1) currently living in the United States; (2) aged 18 years or older; (3) having had a self-reported personal positive COVID-19 test or diagnosis (personal burden survey only); and (4) indicating that they had cared for a friend or family member who had self-reported COVID-19 (caregiver burden survey only).

The analysis for this manuscript included respondents to the personal burden survey (adult sample) and those respondents to the caregiver burden survey who reported on their experience caring for a child <18 years old with COVID-19 illness, as the responses of caregivers of a child can serve as a proxy for pediatric personal burden (child sample). Although not discussed in this manuscript, results for caregivers of adults aged ≥18 years are included in Additional file 2 (Tables S1–S4).

For our primary analysis, to improve data quality and mitigate recall bias, respondents were excluded from the data set if they reported: (1) >10 visits at a care setting (resulting exclusions: adult=18, child=2); (2) both a low severity COVID-19 illness and a hospitalization (adult=5, child=0); (3) that their most recent COVID-19 illness occurring >18 months prior to survey completion (adult=377; child=114); or (4) time spent receiving care >97.5^th^ percentile (adult=32, child=14) (Additional file 2: Table S5 and Figure S1). Speeders (those completing the survey significantly faster than the median completion time) and straightliners (those answering blocks of questions the same way when answers would be expected to differ) were also removed from the dataset by the survey vendor.

A secondary analysis was conducted with more restrictive criteria including: (1) excluding respondents reporting on a COVID-19 illness >12 months prior to survey completion, rather than >18 months (adult=544, child=130) or (2) having completed the survey in less than half the median survey duration (adult=55; child=9).

Analyses were conducted with the full sample that did not have any exclusions to improve data quality. Missing data were treated as missing; no imputation was performed.

### Analysis

Descriptive statistics were calculated for all three sample populations (primary, secondary and no-exclusion) as follows: number of healthcare visits, time spent receiving care, out-of-pocket medical costs, and non-medical costs by care setting for those reporting at least one visit at that setting. Healthcare visit time and travel time were summed to calculate total time spent receiving care.

To explore how costs and financial challenges differed by demographic and health characteristics, we stratified results by insurance status (insured vs uninsured), insurance type (Medicaid vs Other insurance), health condition (comorbid health conditions vs none), and Long COVID status (Long COVID vs no Long COVID).

We used the Strengthening the Reporting of Observational Studies in Epidemiology (STROBE) reporting recommendations for this cross-sectional study [15].

## Results

Eight hundred respondents completed the personal burden survey. Eight hundred respondents completed the caregiver burden survey, with 169 of these reporting care for a 0–17-year-old child who experienced COVID-19. After exclusions for the primary analysis, 409 responses were included from the personal burden survey (adult sample), and 52 responses were included from the caregiver burden survey (child sample) (Additional file 2: Table S5 and Figure S1).

More than 87% of respondents were insured with most receiving insurance through their employer (48% adult; 51% child), Medicare (36% adult), or Medicaid (23% adult; 35% child) (Table 1). Respondents reported mean COVID-19 illness severity level of 3.4 (adult) or 3.3 (child) out of 6. Over thirty-five percent of respondents in the adult sample reported experiencing Long COVID and 25% of the child sample reported that their child experienced Long COVID. Ever having received a COVID-19 vaccine varied by recipient: 80% adult, 56% child.

**Table 1.**
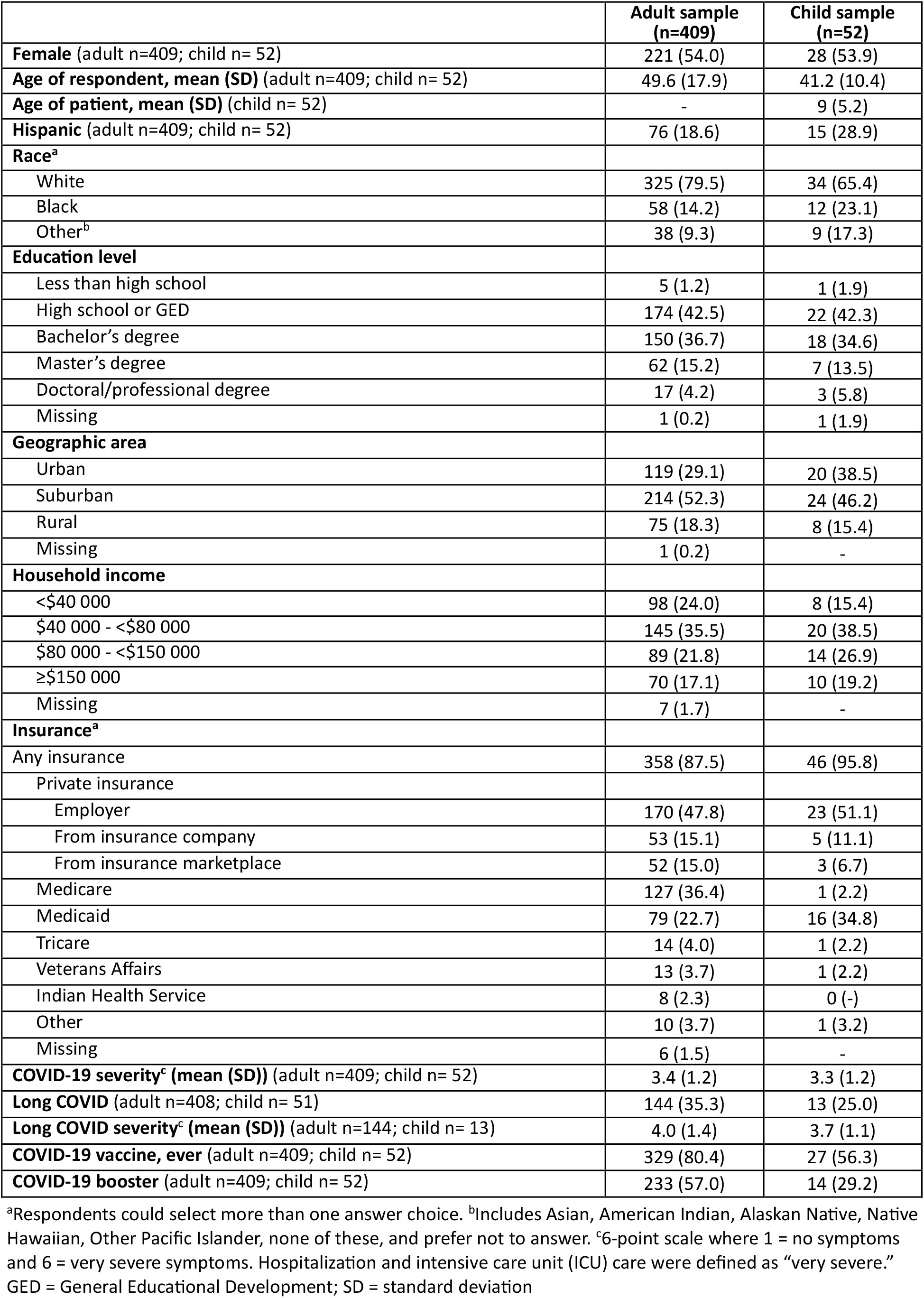
Respondent demographics, Fall 2023, US COVID-19 patients, n (%)

|  | Adult sample<br>(n=409) | Child sample<br>(n=52) |
| --- | --- | --- |
| **Female** (adult n=409; child n= 52) | 221 (54.0) | 28 (53.9) |
| **Age of respondent, mean (SD)** (adult n=409; child n= 52) | 49.6 (17.9) | 41.2 (10.4) |
| **Age of patient, mean (SD)** (child n= 52) | - | 9 (5.2) |
| **Hispanic** (adult n=409; child n= 52) | 76 (18.6) | 15 (28.9) |
| **Race<sup>a</sup>** |  |  |
| White | 325 (79.5) | 34 (65.4) |
| Black | 58 (14.2) | 12 (23.1) |
| Other<sup>b</sup> | 38 (9.3) | 9 (17.3) |
| **Education level** |  |  |
| Less than high school | 5 (1.2) | 1 (1.9) |
| High school or GED | 174 (42.5) | 22 (42.3) |
| Bachelor's degree | 150 (36.7) | 18 (34.6) |
| Master's degree | 62 (15.2) | 7 (13.5) |
| Doctoral/professional degree | 17 (4.2) | 3 (5.8) |
| Missing | 1 (0.2) | 1 (1.9) |
| **Geographic area** |  |  |
| Urban | 119 (29.1) | 20 (38.5) |
| Suburban | 214 (52.3) | 24 (46.2) |
| Rural | 75 (18.3) | 8 (15.4) |
| Missing | 1 (0.2) | - |
| **Household income** |  |  |
| <\$40 000 | 98 (24.0) | 8 (15.4) |
| \$40 000 - <\$80 000 | 145 (35.5) | 20 (38.5) |
| \$80 000 - <\$150 000 | 89 (21.8) | 14 (26.9) |
| ≥\$150 000 | 70 (17.1) | 10 (19.2) |
| Missing | 7 (1.7) | - |
| **Insurance<sup>a</sup>** |  |  |
| Any insurance | 358 (87.5) | 46 (95.8) |
| Private insurance |  |  |
| Employer | 170 (47.8) | 23 (51.1) |
| From insurance company | 53 (15.1) | 5 (11.1) |
| From insurance marketplace | 52 (15.0) | 3 (6.7) |
| Medicare | 127 (36.4) | 1 (2.2) |
| Medicaid | 79 (22.7) | 16 (34.8) |
| Tricare | 14 (4.0) | 1 (2.2) |
| Veterans Affairs | 13 (3.7) | 1 (2.2) |
| Indian Health Service | 8 (2.3) | 0 (-) |
| Other | 10 (3.7) | 1 (3.2) |
| Missing | 6 (1.5) | - |
| **COVID-19 severity<sup>c</sup> (mean (SD))** (adult n=409; child n= 52) | 3.4 (1.2) | 3.3 (1.2) |
| **Long COVID** (adult n=408; child n= 51) | 144 (35.3) | 13 (25.0) |
| **Long COVID severity<sup>c</sup> (mean (SD))** (adult n=144; child n= 13) | 4.0 (1.4) | 3.7 (1.1) |
| **COVID-19 vaccine, ever** (adult n=409; child n= 52) | 329 (80.4) | 27 (56.3) |
| **COVID-19 booster** (adult n=409; child n= 52) | 233 (57.0) | 14 (29.2) |

<sup>a</sup>Respondents could select more than one answer choice. <sup>b</sup>Includes Asian, American Indian, Alaskan Native, Native Hawaiian, Other Pacific Islander, none of these, and prefer not to answer. <sup>c</sup>6-point scale where 1 = no symptoms and 6 = very severe symptoms. Hospitalization and intensive care unit (ICU) care were defined as “very severe.” GED = General Educational Development; SD = standard deviation

Thirty-one percent of the adult sample had sought care from a primary care provider for their COVID-19 illness compared to 73% of the child sample (Figure 1). Urgent care visits (26% adult; 37% child) and emergency department visits (15% adult; 25% child) were also more frequently reported in the child sample than in the adult sample.

**Figure 1.** Respondents who received care for COVID-19 illness, by care setting, Fall 2023, US (%) Percentage of respondents in the adult sample (n=409) and child sample (n=52) reporting at least one visit in each care setting for their most recent COVID-19 illness.

Median (mean) out-of-pocket costs for medical care ranged from $5 ($103) for emergency department visits to $10 ($81) for urgent care visits for the adult sample and ranged from $0 ($81) for emergency department visits to $75 ($132) for hospitalizations for the child sample (Table 2). However, depending on the care setting, between 28% and 59% of the primary sample reported zero out-of-pocket costs for their medical care. Among the subset of respondents with non-zero out-of-pocket costs, mean and median costs were increased compared with all respondents reporting costs (Additional file 2: Table S6). Median (mean) non-medical costs were $28 ($115) or less across care settings in the adult and child samples (Table 2). Median (mean) time spent receiving care varied by care settings and ranged from 98 (182) minutes for primary care visits to 300 (388) minutes for emergency department visits in the adult sample and 90 (96) minutes for primary care visits to 240 (249) minutes for emergency department visits in the child sample.

**Table 2.**
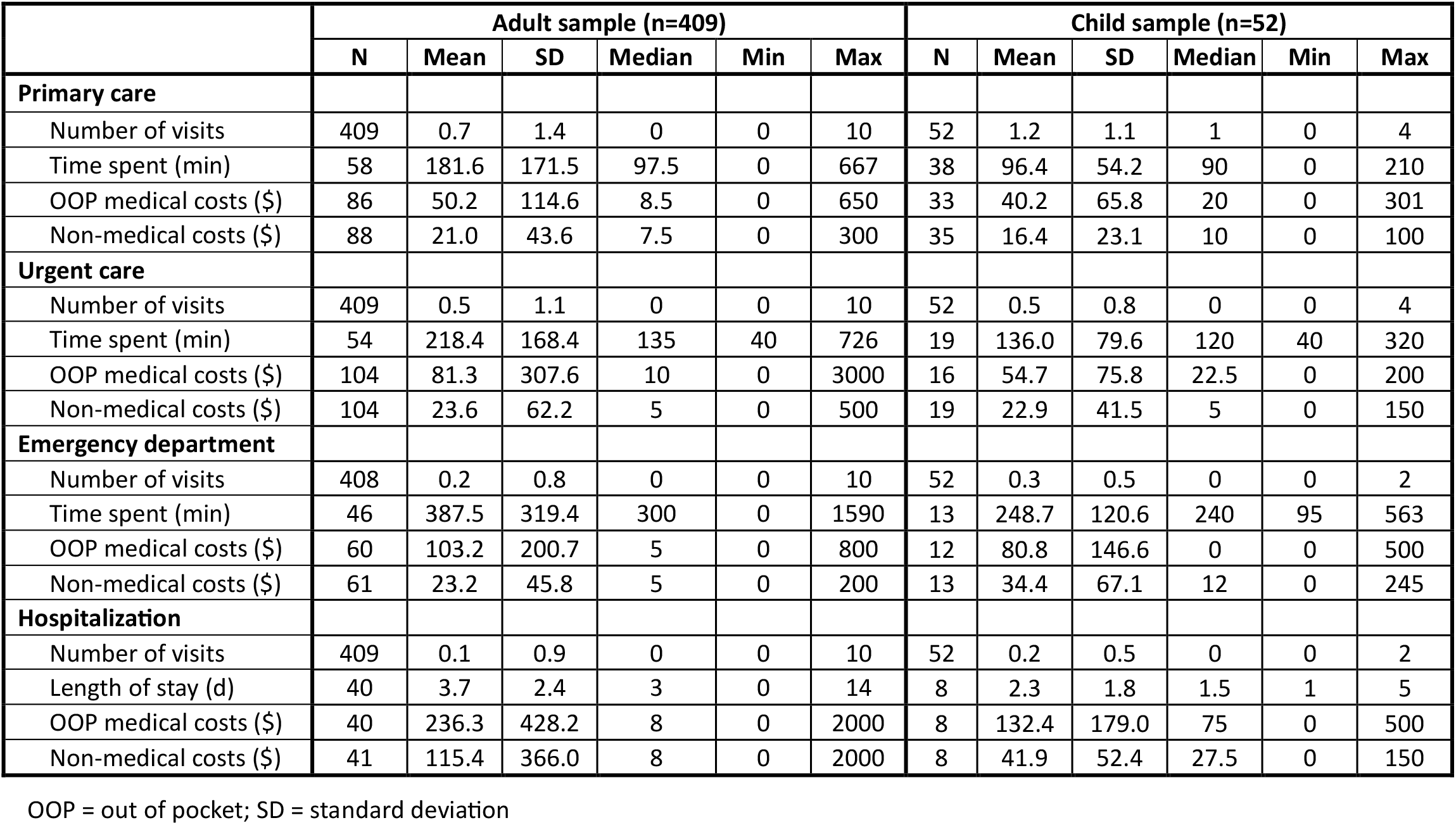
Visits, time spent, and out-of-pocket costs by care setting, Fall 2023, US COVID-19 patients.

Having any type of financial challenge as a result of the most recent COVID-19 illness was reported by 34% of the adult sample and 29% of the child sample (Table 3). The most frequently reported specific type of financial challenge was current problems paying for non-medical bills (11-15%). Twelve percent of respondents in both samples reported problems paying their COVID-19-related medical bills (when they had COVID-19, after they had COVID-19 or currently).

**Table 3.**
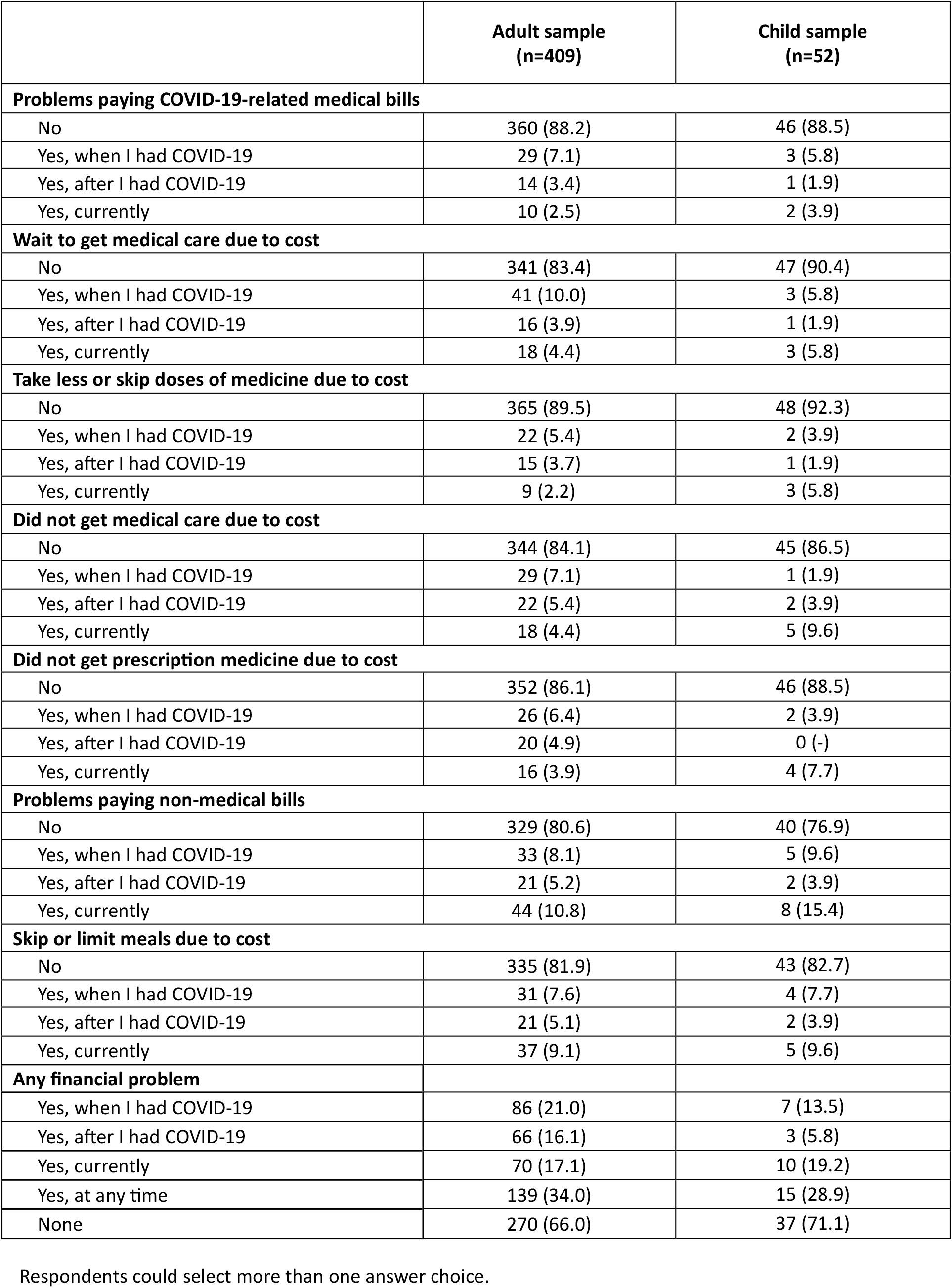
Financial challenges from most recent COVID-19 illness, Fall 2023, US COVID-19 patients, n (%)

|  | Adult sample (n=409) | Child sample (n=52) |
| --- | --- | --- |
| **Problems paying COVID-19-related medical bills** |  |  |
| No | 360 (88.2) | 46 (88.5) |
| Yes, when I had COVID-19 | 29 (7.1) | 3 (5.8) |
| Yes, after I had COVID-19 | 14 (3.4) | 1 (1.9) |
| Yes, currently | 10 (2.5) | 2 (3.9) |
| **Wait to get medical care due to cost** |  |  |
| No | 341 (83.4) | 47 (90.4) |
| Yes, when I had COVID-19 | 41 (10.0) | 3 (5.8) |
| Yes, after I had COVID-19 | 16 (3.9) | 1 (1.9) |
| Yes, currently | 18 (4.4) | 3 (5.8) |
| **Take less or skip doses of medicine due to cost** |  |  |
| No | 365 (89.5) | 48 (92.3) |
| Yes, when I had COVID-19 | 22 (5.4) | 2 (3.9) |
| Yes, after I had COVID-19 | 15 (3.7) | 1 (1.9) |
| Yes, currently | 9 (2.2) | 3 (5.8) |
| **Did not get medical care due to cost** |  |  |
| No | 344 (84.1) | 45 (86.5) |
| Yes, when I had COVID-19 | 29 (7.1) | 1 (1.9) |
| Yes, after I had COVID-19 | 22 (5.4) | 2 (3.9) |
| Yes, currently | 18 (4.4) | 5 (9.6) |
| **Did not get prescription medicine due to cost** |  |  |
| No | 352 (86.1) | 46 (88.5) |
| Yes, when I had COVID-19 | 26 (6.4) | 2 (3.9) |
| Yes, after I had COVID-19 | 20 (4.9) | 0 (-) |
| Yes, currently | 16 (3.9) | 4 (7.7) |
| **Problems paying non-medical bills** |  |  |
| No | 329 (80.6) | 40 (76.9) |
| Yes, when I had COVID-19 | 33 (8.1) | 5 (9.6) |
| Yes, after I had COVID-19 | 21 (5.2) | 2 (3.9) |
| Yes, currently | 44 (10.8) | 8 (15.4) |
| **Skip or limit meals due to cost** |  |  |
| No | 335 (81.9) | 43 (82.7) |
| Yes, when I had COVID-19 | 31 (7.6) | 4 (7.7) |
| Yes, after I had COVID-19 | 21 (5.1) | 2 (3.9) |
| Yes, currently | 37 (9.1) | 5 (9.6) |
| **Any financial problem** |  |  |
| Yes, when I had COVID-19 | 86 (21.0) | 7 (13.5) |
| Yes, after I had COVID-19 | 66 (16.1) | 3 (5.8) |
| Yes, currently | 70 (17.1) | 10 (19.2) |
| Yes, at any time | 139 (34.0) | 15 (28.9) |
| None | 270 (66.0) | 37 (71.1) |

Respondents could select more than one answer choice.

Stratified analyses showed that, in the adult sample, out-of-pocket costs were generally higher and financial challenges were more frequent in respondents who were uninsured, insured through Medicaid, had comorbid health conditions, or experienced Long COVID (Additional file 2: Tables S7–S10).

Analyses of the secondary population sample with more restrictive inclusion criteria and the population sample with no exclusions generated results that were generally similar to the primary analysis except for number of visits when comparing the primary analysis to the no exclusions analysis (Additional file 2: Tables S11–S14).

## Discussion

This study found relatively high rates of healthcare utilization with relatively low out-of-pocket costs in a sample of U.S. COVID-19 patients. Median out-of-pocket costs for medical care were low, ranging from $5 to $10 by care setting in the adult sample and $0 to $75 in the child sample. An important contribution of this study is measuring non-medical out-of-pocket costs. Median non-medical costs were also low at $30 or less across care settings in both the adult and child samples. However, median time spent receiving care ranged from 1.5 to 5 hours and varied by care setting. Financial challenges resulting from the COVID-19 illness were reported in about one third of this sample.

The estimated out-of-pocket medical costs in our study were substantially less than those found in previous studies [10-12], but major methodological differences could account for this. The previous studies had significantly older samples than ours (average age of 67 or older vs. 50) and reported total out-of-pocket medical spending for 3-12 month periods while our study focused on costs from a single healthcare encounter. We also had a substantially lower average length of hospital stay, 3 days versus the 7 days found in Thunell et al. [12], which could be due to increasing immunity in the population and changes in treatment practices over time, among other factors. Our study also relied on self-report rather than administrative data, which could also contribute to differences in estimates. Although administrative data may be a less biased source for medical costs, utilizing self-reports allowed us to measure non-medical and time burden of COVID-19 illness, which cannot be captured through administrative data.

Although out-of-pocket costs at the individual level were relatively low (but highly skewed) in the sampled population, it is important to note that costs of a prevalent infectious disease can represent a substantial cumulative cost on a population level. In addition, COVID-19 treatment costs represented burden for a portion of the sample as 12% of the sample reported problems paying for COVID-19-related medical bills and about 15% avoided getting medical care due to the costs.

Limitations of this analysis include that data was collected from a convenience sample that may not represent the US population. The study was conducted using online, opt-in survey panels which have known data quality risks. In an effort to remove low quality data, multiple exclusion criteria were included in the analysis. However, these exclusions limited the sample size and may have altered the representativeness of the sample. In addition, removing responses with very high care times may have biased burden estimates downward. Small sample sizes in the stratified analyses, particularly for the child sample, may limit the interpretability of the data. It was not possible to identify costs of Long COVID compared to acute COVID, although because this study collected costs on the initial visit for COVID-related illness not ongoing care As more data on Long COVID become available, this may be an important cost driver of COVID-related illness, for both health system and out-of-pocket costs, and should be captured in future studies.

## Conclusions

Our study adds information about time spent receiving care and non-medical costs such as gas, parking, and other travel costs, and meals away from home which are important to capturing the total burden of receiving medical care for an illness on families. These values can be used to inform economic analyses of the burden of COVID-19 and interventions to prevent COVID-19 illness.

## Data Availability

The dataset supporting the conclusions of this article is available in the OSF repository, https://osf.io/zvw28/files/osfstorage

## Abbreviations

CDC: Centers for Disease Control and Prevention
COVID-19: Coronavirus disease 2019
GED: General Educational Development
HaPPI: Healthcare and Public Perceptions of Immunizations
ICU: Intensive care unit
OOP: Out-of-pocket
OSF: Open Science Framework
RSV: Respiratory syncytial virus
SD: Standard deviation
STROBE: Strengthening the Reporting of Observational Studies in Epidemiology

## Declarations

### Ethics approval and consent to participate

This activity was reviewed and approved by the University of Iowa Institutional Review Board #202104132 and the CDC and complied with applicable federal law and CDC policy.^1^

### Consent for publication

Not applicable.

### Availability of data and materials

The dataset supporting the conclusions of this article is available in the OSF repository, https://osf.io/zvw28/files/osfstorage [16]. The survey instruments are included in Additional file 1.

### Authors’ contributions

AMR designed the data analysis plan, contributed to the interpretation of the results, and drafted the manuscript.

AMS led the study design and contributed to data interpretation.

AMG supervised data collection.

AMP contributed to the study design and data interpretation.

CAG contributed to the study design and data interpretation.

NMA contributed to the study design and data interpretation.

MCL conceptualized the study and contributed to data interpretation. AG conducted the data analysis.

JP conceptualized the study, advised on survey design, and contributed to the data analysis plan and the interpretation of the results.

LAP oversaw the data analysis and contributed to the data analysis plan and the interpretation of the results.

All authors read, provided feedback on, and approved the final manuscript.

## Acknowledgements

Not applicable.

## Additional files

**Additional file 1 (.docx)**

*Survey instruments*. Full text of the COVID-19 Personal Burden Survey and COVID-19 Caregiver Burden Survey, including question wording, response options, and survey logic.

**Additional file 2 (.docx)**

*Supplementary tables and figure*. Tables S1–S4: results for caregivers of patients aged ≥18 years; Table S5 and Figure S1: exclusion criteria and number of respondents excluded; Table S6: costs among respondents reporting non-zero costs; Tables S7–S10: costs and financial challenges stratified by insurance status, insurance type, health conditions, and Long COVID status (primary and no-exclusion analyses); Tables S11–S14: comparison of the no-exclusion, primary, and secondary analyses.

## Footnotes

1 See e.g., 45 C.F.R. part 46.102(l)(2), 21 C.F.R. part 56; 42 U.S.C. §241(d); 5 U.S.C. §552a; 44 U.S.C. §3501 et seq.

