## Appendix 2 for "Medical and non-medical out-of-pocket cost and time burden of COVID-19 illness in U.S. children and adults: a cross-sectional survey study"

### Contents

COVID-19 Personal Burden Survey ........ 1

COVID-19 Caregiver Burden Caregiver Survey ........ 38

### COVID-19 Personal Burden Survey

---

***NOTES FOR READER***

- **BLOCK TITLES**: Labels for the question blocks used by the study team but not displayed in the survey.
- **VARIABLE NAMES**: Text in italics and underlined at the beginning of a question that were not displayed in the survey but exported in the data file.
- **RESPONSE VALUES**: Numbers in parentheses at the end of response options indicate the numerically coded response in the data file but were not displayed in the survey.
- **SURVEY LOGIC**: Conditions used to determine survey flow are italicized and highlighted with a specific color to indicate the type of survey flow (see below). Survey logic is included for the reader's benefit and was not displayed in the survey.
  - Red = Screen out logic
    - Respondent routed back to online panel website.
  - Green = Display logic
    - Survey question is displayed if certain conditions are met.
  - Orange = Skip logic
    - Respondents skip certain questions if certain conditions are met.
  - Gray = Page break
  - Italicized text in {*braces*} indicates text that was "piped" in text of a prior response or text that varied depending on a prior response.
- **OTHER NOTES**: Italicized text in [*brackets*] are other notes for the reader and were not displayed in the survey.
- **PIPED TEXT**
  - *LongCovid*: "Did" (or "did") OR "Do" (or "do")
  - *CovidStatus*: "most recent" OR "current"
  - vax_group: "vaccines" OR "vaccination"

---

**Introduction Block**

*We invite you to be in a research study being conducted by investigators from The University of Iowa with funding from the Centers for Disease Control and Prevention. The purpose of the study is to ask adults about important immunization-related issues to inform recommendations and policies.*

*We are inviting you to be in this study because Qualtrics has identified you as someone who is aged 18 years or older. Approximately 1200 respondents will take part in this online survey.*

*If you agree to participate, we would like you to complete a survey regarding your experiences with having COVID-19. Other than questions to determine your eligibility for the study, you are free to skip any questions that you prefer not to answer. It will take approximately 15 minutes to complete the survey.*

*We will keep the information you provide confidential, however federal regulatory agencies and the University of Iowa Institutional Review Board (a committee that reviews and approves research studies) may inspect and copy records pertaining to this research. We will not collect your name or information that can be used to identify you.*

*There are no known risks from being in this study, and you will not benefit personally. However, we hope that others may benefit in the future from what we learn because of this study.*

*There are no costs for being in this study and being in this study is up to you. If you do not wish to be in the study, you may close the browser window now or at any time before submitting the survey. Submitting the survey will indicate your willingness to be in the study. You will be compensated according to the terms of your panel agreement.*

*If you have any questions about the study itself or experience a research-related injury, please contact Dr. Aaron Scherer ( or 319.384.6180). If you have questions about the rights of research subjects, please contact the Human Subjects Office, 105 Hardin Library for the Health Sciences, 600 Newton Rd, The University of Iowa, Iowa City, IA 52242-1098, (319) 335-6564, or. To offer input about your experiences as a research subject or to speak to someone other than the research staff, call the Human Subjects Office at the number above.*

*Thank you very much for your consideration.*

*Sincerely,*

*Aaron Scherer, PhD*
*Assistant Professor of Internal Medicine*

*Page Break*

*consent* Please only click 'Yes' to indicate that you have read and understand the above information, are fully aware of the risks, your right to withdraw at any point, and desire of your own free will to participate in this research study.

- No (2)
- Yes (1)

*Participants screened out of survey if consent response was "*No*".*

*Page Break*

*quality We care about the quality of our data. For us to get the most accurate measures of your opinions, it is important that you thoughtfully provide your best answers to each question in this survey.*

Do you commit to thoughtfully provide your best answers to each question in this survey?

- I will provide my best answers (1)
- I will not provide my best answers (2)
- I can't promise either way (3)

*Participants screened out of survey if quality response was "*I will not provide my best answers*" or "*I can't promise either way*".*

*Page Break*

**Demographics Block**
*age* What is your age?

________________________________________________________________

*gender* Do you currently describe yourself as male, female, or transgender?
- ○ Male (1)
- ○ Female (2)
- ○ Transgender (3)
- ○ None of these (4)
- ○ Prefer not to say (5)

*ethnic* Do you consider yourself to be Hispanic or Latino/a?
- ○ Hispanic or Latino/a (1)
- ○ Not Hispanic or Latino/a (2)
- ○ None of these (3)
- ○ Prefer not to answer (4)

*race* What race or races do you consider yourself to be? *Select all that apply.*
- ▢ American Indian or Alaskan Native (1)
- ▢ Asian (2)
- ▢ Black or African American (3)
- ▢ Native Hawaiian or Other Pacific Islander (4)
- ▢ White (5)
- ▢ ⊗None of these (6)
- ▢ ⊗Prefer not to answer (7)

*covid_status* Have you ever tested positive for COVID-19 (using a rapid point-of-care test, self-test, or laboratory test) or been told by a doctor or other health care provider that you have or had COVID-19?
- ○ No (1)
- ○ Yes (2)
- ○ Unsure (3)
- ○ Prefer not to say (4)

*Page Break*

**Information About Most Recent COVID-19 Block**
*covid_date* When did you most recently test positive for (or were told you had) COVID-19?

| Month | Year |
|---|---|
| ▼ January (1 ... December (12) | ▼ 2020 (1 ... 2023 (4) |

[*A pulldown menu was displayed with the months in chronological order, and a pulldown menu was displayed with the years in chronological order*.]

*covid_severe* How severe were your symptoms when they were at their worst for your most recent COVID-19? Symptoms resulting in hospitalization or being admitted to the ICU would be considered "very severe."

| 1 | 2 | 3 | 4 | 5 | 6 |
|---|---|---|---|---|---|
| No symptoms | | | | | Very severe symptoms |

*covid_current* Do you currently have any COVID-19 symptoms?

| | |
|---|---|
| No (2) | Yes (1) |

*Page Break*

**Additional COVID-19 Information Block**
*covid_long* *{LongCovid}* you have any symptoms lasting 3 months or longer that you did not have prior to when you most recently had COVID-19 (in other words, *Long COVID symptoms*)?

*Long COVID symptoms may include: tiredness or fatigue, difficulty thinking or concentrating, forgetfulness, or memory problems (sometimes referred to as "brain fog"), difficulty breathing or shortness of breath, joint or muscle pain, fast-beating or pounding heart (also known as heart palpitations), chest pain, dizziness on standing, changes to your menstrual cycle, changes to taste/smell, or inability to exercise.*

No (2) Yes (1)

*covid_symptoms* How much do these Long COVID symptoms reduce your ability to carry out day-to-day activities compared with the time before you had COVID-19?

| 1 | 2 | 3 | 4 | 5 | 6 |
|---|---|---|---|---|---|
| Not at all | | | | | A lot |

*Page Break*

*Answer the following questions based on when you most recently had COVID-19.*

*covid_provider* When you most recently had COVID-19, were you seen by a healthcare provider, such as a doctor or nurse, at any of these locations (including telehealth/virtual visits) for your COVID-19?

| | No (1) | Yes (2) |
|---|---|---|
| **Primary Care Clinic** (1) | ○ | ○ |
| **Urgent Care Clinic** (2) | ○ | ○ |
| **Emergency Room (ER)** (3) | ○ | ○ |
| **Specialty Clinic** Examples: Pulmonology (lung) or cardiology (heart) clinic (4) | ○ | ○ |
| **In-Patient Rehabilitation Clinic** (5) | ○ | ○ |

*covid_hosp* Were you ever hospitalized as a result of your COVID-19 *when you most recently had COVID-19*?

No (2) Yes (1)

*covid_home* Did you receive any home health care services (example: in-home rehabilitation) as a result of your most recent COVID-19?

No (2) Yes (1)

*covid_meds* Did you take any over-the-counter (non-prescription) or prescription medicines to treat, or as a result of, your most recent COVID-19? *(Examples: cold/cough medicine, decongestants, fever/pain reducers, inhalers or nebulizers, antibiotics for complications of COVID-19. Do not include medications taken during a hospitalization.)*

No (2) Yes (1)

*Page Break*

*Primary Care Visits Block was displayed if the covid_provider response was* "Yes" *to* "Primary Care Clinic".

**Primary Care Visits Block**

*Now we are going to ask you about primary care (non-specialist) you received when you most recently had COVID-19, which could include care for long-term symptoms (Long COVID).*

*primary_num* How many times did you visit a primary care (non-specialist) clinic for your most recent COVID-19?

________________________________________________________________

*primary_virtual* How many of these visits were virtual?

________________________________________________________________

*Page Break*

*Answer the following questions based on when you most recently had COVID-19.*

*primary_date* When was the first time you went to a primary care (non-specialist) clinic for your most recent COVID-19?

| Month | Year |
|---|---|
| ▼ January (1 ... December (12) | ▼ 2020 (1 ... 2023 (4) |

[*A pulldown menu was displayed with the months in chronological order, and a pulldown menu was displayed with the years in chronological order*.]

*primary_travel* How long did it take you to travel to the primary care clinic?

_____ hours _____ minutes

*primary_wait* How much time did you spend at the clinic (including wait and visit times)?

_____ hours _____ minutes

*primary_last was displayed if the primary_num response was greater than one.*

*primary_last* When was the last time you went to a primary care clinic for your most recent COVID-19?

| Month | Year |
| --- | --- |
| ▼ January (1 ... December (12) | ▼ 2020 (1 ... 2023 (4) |

[*A pulldown menu was displayed with the months in chronological order, and a pulldown menu was displayed with the years in chronological order*.]

*Page Break*

*Now we are going to ask you about out-of-pocket costs you might have had related to your first primary care visit when you most recently had COVID-19.*

***Out-of-pocket costs** are payments you made that were not reimbursed (paid back) by an insurance company.*

*If you can't remember the exact details, please provide your best answer possible based on what you can remember.*

*primary_direct* How much did you pay out-of-pocket for direct medical costs - such as visit fees, copays, deductibles, lab fees, x-ray - related to the first time you went to a primary care clinic for your most recent COVID-19? *Round to the nearest $10 value.*

________________________________________________________________

*primary_non* How much did you pay for non-medical costs - such as gas, meals, hotel stays, and other similar costs - related to the first time you went to a primary care clinic for your most recent COVID-19? *Round to the nearest $10 value.*

________________________________________________________________

*primary_desc* Please describe any non-medical costs you had. *Type "None" if you did not have any non-medical costs.*

________________________________________________________________

*Page Break*

*Urgent Care Visits Block was displayed if the covid_provider response was* "Yes" *to* "Urgent Care Clinic".

**Urgent Care Visits Block**

*Now we are going to ask you about urgent care you received when you most recently had COVID-19, which could include care for long-term symptoms (Long COVID).*

*urgent_num* How many times did you visit an urgent care clinic for your most recent COVID-19?

________________________________________________________________

*Page Break*

*Answer the following questions based on when you most recently had COVID-19. If you can't remember the exact details, please provide your best answer possible based on what you can remember.*

*urgent_date* When was the first time you went to an urgent care clinic for your most recent COVID-19?

| Month | Year |
|---|---|
| ▼ January (1 ... December (12) | ▼ 2020 (1 ... 2023 (4) |

[*A pulldown menu was displayed with the months in chronological order, and a pulldown menu was displayed with the years in chronological order*.]

*urgent_travel* How long did it take you to travel to the urgent care clinic?

_____ hours _____ minutes

*urgent_wait* How much time did you spend at the clinic (including wait and visit times)?

_____ hours _____ minutes

*urgent_last was displayed if the urgent_num response was greater than one.*

*urgent_last* When was the last time you went to an urgent care clinic for your most recent COVID-19?

| Month | Year |
|---|---|
| ▼ January (1 ... December (12) | ▼ 2020 (1 ... 2023 (4) |

[*A pulldown menu was displayed with the months in chronological order, and a pulldown menu was displayed with the years in chronological order*.]

*Page Break*

*Now we are going to ask you about out-of-pocket costs you might have had related to your first urgent care visit when you most recently had COVID-19.*

***Out-of-pocket costs** are payments you made that were not reimbursed (paid back) by an insurance company.*

*If you can't remember the exact details, please provide your best answer possible based on what you can remember.*

*urgent_direct* How much did you pay out-of-pocket for direct medical costs - such as visit fees, copays, deductibles, lab fees, x-ray - related to the first time you went to an urgent care clinic for your most recent COVID-19? *Round to the nearest $10 value.*

________________________________________________________________

*urgent_non* How much did you pay for non-medical costs - such as gas, meals, hotel stays, and other similar costs - related to the first time you went to an urgent care clinic for your most recent COVID-19? *Round to the nearest $10 value.*

________________________________________________________________

*urgent_desc* Please describe any non-medical costs you had. *Type "None" if you did not have any non-medical costs.*

________________________________________________________________

*Page Break*

*Hospitalizations Block was displayed if the covid_hosp response was* "Yes".

**Hospitalizations Block**

*Now we are going to ask you about hospitalizations you experienced when you most recently had COVID-19, which could include care for long-term symptoms (Long COVID).*

*hosp_num* How many times were you hospitalized because of your most recent COVID-19?

________________________________________________________________

*Page Break*

*Answer the following questions based on when you most recently had COVID-19. If you can't remember the exact details, please provide your best answer possible based on what you can remember.*

*hosp_date* When was the first time you were hospitalized for your most recent COVID-19?

| Month | Year |
|---|---|
| ▼ January (1 ... December (12) | ▼ 2020 (1 ... 2023 (4) |

[*A pulldown menu was displayed with the months in chronological order, and a pulldown menu was displayed with the years in chronological order*.]

*hosp_days* How many days did you stay at the hospital?

________________________________________

*hosp_last was displayed if the hosp_num response was greater than one.*

*hosp_last* When was the last time you were hospitalized for your most recent COVID-19?

| Month | Year |
|---|---|
| ▼ January (1 ... December (12) | ▼ 2020 (1 ... 2023 (4) |

[*A pulldown menu was displayed with the months in chronological order, and a pulldown menu was displayed with the years in chronological order*.]

*hosp_ldays was displayed if the hosp_num was greater than one.*

*hosp_ldays* How many days did you stay at the hospital?

________________________________________

*Now we are going to ask you about out-of-pocket costs you might have had related to the first time you were hospitalized as a result of your {CovidStatus} COVID-19.*

***Out-of-pocket costs** are payments you made that were not reimbursed (paid back) by an insurance company.*

*If you can't remember the exact details, please provide your best answer possible based on what you can remember.*

*hosp_direct* How much did you pay out-of-pocket for direct medical costs - such as visit fees, copays, deductibles, lab fees, x-ray - related to the first time you were hospitalized for your most recent COVID-19? *Round to the nearest $10 value.*

________________________________________

*hosp_non* How much did you pay for non-medical costs - such as gas, meals, hotel stays, and other similar costs - related to the first time you were hospitalized for your most recent COVID-19? *Round to the nearest $10 value.*

________________________________________

*hosp_desc* Please describe any non-medical costs you had. *Type "None" if you did not have any non-medical costs.*

________________________________________________________________

*Page Break*

*Emergency Room Visits Block was displayed if the covid_provider response was* "Yes" *to* "Emergency Room (ER)".

**Emergency Room Visits Block**

*Now we are going to ask you about visits to the emergency room (ER) when you most recently had COVID-19, which could include care for long-term symptoms (Long COVID).*

*Answer the following questions based on when you most recently had COVID-19.*

*er_num* How many times did you visit an emergency room (ER) for your most recent COVID-19?

________________________________________________________________

*Page Break*

*Answer the following questions based on when you most recently had COVID-19. If you can't remember the exact details, please provide your best answer possible based on what you can remember.*

*er_date* When was the first time you went to an emergency room for your most recent COVID-19?

| Month | Year |
|---|---|
| ▼ January (1 ... December (12) | ▼ 2020 (1 ... 2023 (4) |

[*A pulldown menu was displayed with the months in chronological order, and a pulldown menu was displayed with the years in chronological order*.]

*er_travel* How long did it take you to travel to the emergency room?

_____ hours _____ minutes

*er_wait* How much time did you spend at the emergency room (including wait and visit times)?

_____ hours _____ minutes

*er_last was displayed if the er_num response was greater than one.*

er_last When was the last time you went to an emergency room for your most recent COVID-19?

| Month | Year |
| --- | --- |
| ▼ January (1 ... December (12) | ▼ 2020 (1 ... 2023 (4) |

[*A pulldown menu was displayed with the months in chronological order, and a pulldown menu was displayed with the years in chronological order*.]

*Page Break*

*Now we are going to ask you about out-of-pocket costs you might have had related to your first emergency room visit when you most recently had COVID-19.*

***Out-of-pocket costs*** *are payments you made that were not reimbursed (paid back) by an insurance company.*

*If you can't remember the exact details, please provide your best answer possible based on what you can remember.*

er_direct How much did you pay out-of-pocket for direct medical costs - such as visit fees, copays, deductibles, lab fees, x-ray - related to the first time you went to an emergency room for your most recent COVID-19? *Round to the nearest $10 value.*

________________________________________________________________

er_non How much did you pay for non-medical costs - such as gas, meals, hotel stays, and other similar costs - related to the first time you went to an emergency room for your most recent COVID-19? *Round to the nearest $10 value.*

________________________________________________________________

er_desc Please describe any non-medical costs you had. *Type "None" if you did not have any non-medical costs.*

________________________________________________________________

*Page Break*

*Medical Specialist Visits Block was displayed if the covid_provider response was "Yes" to* "Specialty Clinic Examples: Pulmonology (lung) or cardiology (heart) clinic".

**Medical Specialist Visits Block**

*Now we are going to ask you about visits to a specialty care clinic (examples: pulmonology (lung) or cardiology (heart) clinic) you received when you most recently had COVID-19, which could include care for long-term symptoms (Long COVID).*

*spec_visits* Did you have any outpatient visits with any of the following specialists for your most recent COVID-19? *Select all that apply.*

- ☐ Infectious disease specialist (1)
- ☐ Pulmonologist (lung doctor) (2)
- ☐ Cardiologist (heart doctor) (3)
- ☐ Oncologist (cancer doctor) (4)
- ☐ Obstetrician/Gynecologist (OB/GYN) (5)
- ☐ Physical therapist (6)
- ☐ Occupational therapist (7)
- ☐ Other - please specify: (8) ________________________________________
- ☐ ⊗None of these (9)

*Page Break*

*Infectious Disease Intro Block and Infectious Disease Visits Block were displayed if the spec_type response was* "Infectious disease specialist".

**Infectious Disease Specialist Visits Block**

*Answer the following questions based on when you most recently had COVID-19.*

*id_num* How many times did you visit an infectious disease specialist clinic for your most recent COVID-19?

________________________________________________________________

*Page Break*

*Answer the following questions based on when you most recently had COVID-19. If you can't remember the exact details, please provide your best answer possible based on what you can remember.*

*id_date* When was the first time you went to an infectious disease specialist clinic for your most recent COVID-19?

| Month | Year |
| --- | --- |
| ▼ January (1 ... December (12) | ▼ 2020 (1 ... 2023 (4) |

[*A pulldown menu was displayed with the months in chronological order, and a pulldown menu was displayed with the years in chronological order*.]

*id_travel* How long did it take you to travel to the infectious disease specialist clinic?

_____ hours _____ minutes

*id_wait* How much time did you spend at the infectious disease specialist clinic (including wait and visit times)?

_____ hours _____ minutes

*id_last was displayed if the id_num response was greater than one.*

*id_last* When was the last time you went to an infectious disease specialist clinic for your most recent COVID-19?

| Month | Year |
| --- | --- |
| ▼ January (1 ... December (12) | ▼ 2020 (1 ... 2023 (4) |

[*A pulldown menu was displayed with the months in chronological order, and a pulldown menu was displayed with the years in chronological order*.]

*Now we are going to ask you about out-of-pocket costs you might have had related to your first infectious disease specialist clinic visit when you most recently had COVID-19.*

***Out-of-pocket costs** are payments you made that were not reimbursed (paid back) by an insurance company.*

*If you don't know or can't remember the exact details, please provide your best answer possible based on what you can remember.*

*id_direct* How much did you pay out-of-pocket for direct medical costs - such as visit fees, copays, deductibles, lab fees, x-ray - related to the first time you went to the infectious disease specialist clinic for your most recent COVID-19? *Round to the nearest $10 value.*

______________________________________________

*id_non* How much did you pay for non-medical costs - such as gas, meals, hotel stays, and other similar costs - related to the first time you went to the infectious disease specialist clinic for your most recent COVID-19? *Round to the nearest $10 value.*

________________________________________________________________

*id_desc* Please describe any non-medical costs you had. *Type "None" if you did not have any non-medical costs.*

________________________________________________________________

*Page Break*

*Pulmonology Intro Block and Pulmonology Visits Block were displayed if the spec_type response was* "Pulmonologist (lung doctor)".

**Pulmonology Visits Block**

*Answer the following questions based on when you most recently had COVID-19.*

*pulm_num* How many times did you visit a pulmonology clinic for your most recent COVID-19?

________________________________________________________________

*Page Break*

*Answer the following questions based on when you most recently had COVID-19. If you can't remember the exact details, please provide your best answer possible based on what you can remember.*

*pulm_date* When was the first time you went to a pulmonology clinic for your most recent COVID-19?

| Month | Year |
|---|---|
| ▼ January (1 ... December (12) | ▼ 2020 (1 ... 2023 (4) |

[*A pulldown menu was displayed with the months in chronological order, and a pulldown menu was displayed with the years in chronological order*.]

*pulm_travel* How long did it take you to travel to the pulmonology clinic?

_____ hours _____ minutes

*pulm_wait* How much time did you spend at the pulmonology clinic (including wait and visit times)?

_____ hours _____ minutes

*pulm_last was displayed if the pulm_num response was greater than one.*
*pulm_last* When was the last time you went to a pulmonology clinic for your most recent COVID-19?

| Month | Year |
| --- | --- |
| ▼ January (1 ... December (12) | ▼ 2020 (1 ... 2023 (4) |

[*A pulldown menu was displayed with the months in chronological order, and a pulldown menu was displayed with the years in chronological order*.]

*Now we are going to ask you about out-of-pocket costs you might have had related to your first pulmonology clinic visit when you most recently had COVID-19.*

***Out-of-pocket costs** are payments you made that were not reimbursed (paid back) by an insurance company.*

*If you don't know or can't remember the exact details, please provide your best answer possible based on what you can remember.*

*pulm_direct* How much did you pay out-of-pocket for direct medical costs - such as visit fees, copays, deductibles, lab fees, x-ray - related to the first time you went to the pulmonology clinic for your most recent COVID-19? *Round to the nearest $10 value.*

________________________________________________________________

*pulm_non* How much did you pay for non-medical costs - such as gas, meals, hotel stays, and other similar costs - related to the first time you went to the pulmonology clinic for your most recent COVID-19? *Round to the nearest $10 value.*

________________________________________________________________

*pulm_desc* Please describe any non-medical costs you had. *Type "None" if you did not have any non-medical costs.*

________________________________________________________________

*Page Break*

*Cardiology Intro Block and Cardiology Visits Block were displayed if the spec_type response was* "Cardiologist (heart doctor)".

**Cardiology Visits Block**
*Answer the following questions based on when you most recently had COVID-19.*

*card_num* How many times did you visit a cardiology clinic for your most recent COVID-19?

________________________________________________________________

*Page Break*

*Answer the following questions based on when you most recently had COVID-19. If you can't remember the exact details, please provide your best answer possible based on what you can remember.*

*card_date* When was the first time you went to a cardiology clinic for your most recent COVID-19?

| Month | Year |
|---|---|
| ▼ January (1 ... December (12) | ▼ 2020 (1 ... 2023 (4) |

[*A pulldown menu was displayed with the months in chronological order, and a pulldown menu was displayed with the years in chronological order*.]

*card_travel* How long did it take you to travel to the cardiology clinic?

_____ hours _____ minutes

*card_wait* How much time did you spend at the cardiology clinic (including wait and visit times)?

_____ hours _____ minutes

*card_last was displayed if the card_num response was greater than one.*

*card_last* When was the last time you went to a cardiology clinic for your most recent COVID-19?

| Month | Year |
|---|---|
| ▼ January (1 ... December (12) | ▼ 2020 (1 ... 2023 (4) |

[*A pulldown menu was displayed with the months in chronological order, and a pulldown menu was displayed with the years in chronological order*.]

*Now we are going to ask you about out-of-pocket costs you might have had related to your first pulmonology clinic visit when you most recently had COVID-19.*

***Out-of-pocket costs** are payments you made that were not reimbursed (paid back) by an insurance company.*

*If you don't know or can't remember the exact details, please provide your best answer possible based on what you can remember.*

*card_direct* How much did you pay out-of-pocket for direct medical costs - such as visit fees, copays, deductibles, lab fees, x-ray - related to the first time you went to the cardiology clinic for your most recent COVID-19? *Round to the nearest $10 value.*

________________________________________________________________

*card_non* How much did you pay for non-medical costs - such as gas, meals, hotel stays, and other similar costs - related to the first time you went to the cardiology clinic for your most recent COVID-19? *Round to the nearest $10 value.*

________________________________________________________________

*card_desc* Please describe any non-medical costs you had. *Type "None" if you did not have any non-medical costs.*

________________________________________________________________

*Page Break*

*Oncology Intro Block and Oncology Visits Block were displayed if the spec_type response was* "Oncologist (cancer doctor)".

**Oncology Visits Block**

*Answer the following questions based on when you most recently had COVID-19.*

*onc_num* How many times did you visit an oncology clinic for your most recent COVID-19?

________________________________________________________________

*Page Break*

*Answer the following questions based on when you most recently had COVID-19. If you can't remember the exact details, please provide your best answer possible based on what you can remember.*

*onc_date* When was the first time you went to an oncology clinic for your most recent COVID-19?

| Month | Year |
| --- | --- |
| ▼ January (1 ... December (12) | ▼ 2020 (1 ... 2023 (4) |

[*A pulldown menu was displayed with the months in chronological order, and a pulldown menu was displayed with the years in chronological order*.]

*onc_travel* How long did it take you to travel to the oncology clinic?

_____ hours _____ minutes

*onc_wait* How much time did you spend at the oncology clinic (including wait and visit times)?

_____ hours _____ minutes

*onc_last was displayed if the onc_num was greater than one.*

*onc_last* When was the last time you went to an oncology clinic for your most recent COVID-19?

| Month | Year |
| --- | --- |
| ▼ January (1 ... December (12) | ▼ 2020 (1 ... 2023 (4) |

[*A pulldown menu was displayed with the months in chronological order, and a pulldown menu was displayed with the years in chronological order*.]

*Now we are going to ask you about out-of-pocket costs you might have had related to your first oncology clinic visit when you most recently had COVID-19.*

***Out-of-pocket costs** are payments you made that were not reimbursed (paid back) by an insurance company.*

*If you don't know or can't remember the exact details, please provide your best answer possible based on what you can remember.*

*onc_direct* How much did you pay out-of-pocket for direct medical costs - such as visit fees, copays, deductibles, lab fees, x-ray - related to the first time you went to the oncology clinic for your most recent COVID-19? *Round to the nearest $10 value.*

______________________________________________

*onc_non* How much did you pay for non-medical costs - such as gas, meals, hotel stays, and other similar costs - related to the first time you went to the oncology clinic for your most recent COVID-19? *Round to the nearest $10 value.*

________________________________________________________________

*onc_desc* Please describe any non-medical costs you had. *Type "None" if you did not have any non-medical costs.*

________________________________________________________________

*Page Break*

*OB/GYN Intro Block and OB/GYN Visits Block were displayed if the spec_type response was* “Obstetrician/Gynecologist (OB/GYN)”.

**OB/GYN Visits Block**

*Answer the following questions based on when you most recently had COVID-19.*

*ob_num* How many times did you visit an obstetrics/gynecology clinic for your most recent COVID-19?

________________________________________________________________

*Page Break*

*Answer the following questions based on when you most recently had COVID-19. If you can’t remember the exact details, please provide your best answer possible based on what you can remember.*

*ob_date* When was the first time you went to an obstetrics/gynecology clinic for your most recent COVID-19?

| Month | Year |
|---|---|
| ▼ January (1 ... December (12) | ▼ 2020 (1 ... 2023 (4) |

[*A pulldown menu was displayed with the months in chronological order, and a pulldown menu was displayed with the years in chronological order*.]

*ob_travel* How long did it take you to travel to the obstetrics/gynecology clinic?

_____ hours _____ minutes

*ob_wait* How much time did you spend at the obstetrics/gynecology clinic (including wait and visit times)?

_____ hours _____ minutes

*ob_last was displayed if the ob_num response was greater than one.*

*ob_last* When was the last time you went to an obstetrics/gynecology clinic for your most recent COVID-19?

| Month | Year |
| --- | --- |
| ▼ January (1 ... December (12) | ▼ 2020 (1 ... 2023 (4) |

[*A pulldown menu was displayed with the months in chronological order, and a pulldown menu was displayed with the years in chronological order*.]

*Now we are going to ask you about out-of-pocket costs you might have had related to your first obstetrics/gynecology clinic visit when you most recently had COVID-19.*

***Out-of-pocket costs** are payments you made that were not reimbursed (paid back) by an insurance company.*

*If you don't know or can't remember the exact details, please provide your best answer possible based on what you can remember.*

*ob_direct* How much did you pay out-of-pocket for direct medical costs - such as visit fees, copays, deductibles, lab fees, x-ray - related to the first time you went to the obstetrics/gynecology clinic for your most recent COVID-19? *Round to the nearest $10 value.*

________________________________________________________________

*ob_non* How much did you pay for non-medical costs - such as gas, meals, hotel stays, and other similar costs - related to the first time you went to the obstetrics/gynecology clinic for your most recent COVID-19? *Round to the nearest $10 value.*

________________________________________________________________

*ob_desc* Please describe any non-medical costs you had. *Type "None" if you did not have any non-medical costs.*

________________________________________________________________

*Page Break*

*Physical Therapy Intro Block and Physical Therapy Visits Block were displayed if the spec_type response was* "Physical therapist".

**Physical Therapy Visits Block**

*Answer the following questions based on when you most recently had COVID-19.*

*pt_num* How many times did you visit a physical therapy clinic for your most recent COVID-19?

________________________________________________________________

*Answer the following questions based on when you most recently had COVID-19. If you can't remember the exact details, please provide your best answer possible based on what you can remember.*

*pt_date* When was the first time you went to a physical therapy clinic for your most recent COVID-19?

| Month | Year |
| --- | --- |
| ▼ January (1 ... December (12) | ▼ 2020 (1 ... 2023 (4) |

[*A pulldown menu was displayed with the months in chronological order, and a pulldown menu was displayed with the years in chronological order*.]

*pt_travel* How long did it take you to travel to the physical therapy clinic?

_____ hours _____ minutes

*pt_wait* How much time did you spend at the physical therapy clinic (including wait and visit times)?

_____ hours _____ minutes

*pt_last was displayed if the pt_num response was greater than one.*

*pt_last* When was the last time you went to a physical therapy clinic for your most recent COVID-19?

| Month | Year |
| --- | --- |
| ▼ January (1 ... December (12) | ▼ 2020 (1 ... 2023 (4) |

[*A pulldown menu was displayed with the months in chronological order, and a pulldown menu was displayed with the years in chronological order*.]

*Now we are going to ask you about out-of-pocket costs you might have had related to your first physical therapy clinic visit when you most recently had COVID-19.*

***Out-of-pocket costs** are payments you made that were not reimbursed (paid back) by an insurance company.*

*If you don't know or can't remember the exact details, please provide your best answer possible based on what you can remember.*

*pt_direct* How much did you pay out-of-pocket for direct medical costs - such as visit fees, copays, deductibles, lab fees, x-ray - related to the first time you went to the physical therapy clinic for your most recent COVID-19? *Round to the nearest $10 value.*

________________________________________________________________

*pt_non* How much did you pay for non-medical costs - such as gas, meals, hotel stays, and other similar costs - related to the first time you went to the physical therapy clinic for your most recent COVID-19? *Round to the nearest $10 value.*

________________________________________________________________

*pt_desc* Please describe any non-medical costs you had. *Type "None" if you did not have any non-medical costs.*

________________________________________________________________

*Page Break*

*Occupational Therapy Intro Block and Occupational Therapy Visits Block were displayed if the spec_type response was* "Occupational therapist".

**Occupational Therapy Visits Block**

*Answer the following questions based on when you most recently had COVID-19.*

*ot_num* How many times did you visit an occupational therapy clinic for your most recent COVID-19?

________________________________________________________________

*Page Break*

*Answer the following questions based on when you most recently had COVID-19. If you can't remember the exact details, please provide your best answer possible based on what you can remember.*

*ot_date* When was the first time you went to an occupational therapy clinic for your most recent COVID-19?

| Month | Year |
|---|---|
| ▼ January (1 ... December (12) | ▼ 2020 (1 ... 2023 (4) |

[*A pulldown menu was displayed with the months in chronological order, and a pulldown menu was displayed with the years in chronological order*.]

*ot_travel* How long did it take you to travel to the occupational therapy clinic?

_____ hours _____ minutes

*ot_wait* How much time did you spend at the occupational therapy clinic (including wait and visit times)?

_____ hours _____ minutes

*ot_last was displayed if the ot_num response was greater than one.*

*ot_last* When was the last time you went to an occupational therapy clinic for your most recent COVID-19?

| Month | Year |
|---|---|
| ▼ January (1 ... December (12) | ▼ 2020 (1 ... 2023 (4) |

[*A pulldown menu was displayed with the months in chronological order, and a pulldown menu was displayed with the years in chronological order*.]

*Now we are going to ask you about out-of-pocket costs you might have had related to your first occupational therapy clinic visit when you most recently had COVID-19.*

***Out-of-pocket costs** are payments you made that were not reimbursed (paid back) by an insurance company.*

*If you don't know or can't remember the exact details, please provide your best answer possible based on what you can remember.*

*ot_direct* How much did you pay out-of-pocket for direct medical costs - such as visit fees, copays, deductibles, lab fees, x-ray - related to the first time you went to the occupational therapy clinic for your most recent COVID-19? *Round to the nearest $10 value.*

______________________________________________

*ot_non* How much did you pay for non-medical costs - such as gas, meals, hotel stays, and other similar costs - related to the first time you went to the occupational therapy clinic for your most recent COVID-19? *Round to the nearest $10 value.*

________________________________________________________________

*ot_desc* Please describe any non-medical costs you had. *Type "None" if you did not have any non-medical costs.*

________________________________________________________________

*Page Break*

*Other Visits Intro Block and Other Visits Block were displayed if the spec_type response was* "Other - please specify:".

**Other Specialty Visits Block**

*Answer the following questions based on when you most recently had COVID-19.*

*other_num* How many times did you visit the {*spec_visits Other response}* clinic for your most recent COVID-19?

________________________________________________________________

*Page Break*

*Answer the following questions based on when you most recently had COVID-19. If you can't remember the exact details, please provide your best answer possible based on what you can remember.*

*other_date* When was the first time you went to the {*spec_visits Other response}* clinic for your most recent COVID-19?

| Month | Year |
| --- | --- |
| ▼ January (1 ... December (12) | ▼ 2020 (1 ... 2023 (4) |

[*A pulldown menu was displayed with the months in chronological order, and a pulldown menu was displayed with the years in chronological order*.]

*other_travel* How long did it take you to travel to the {*spec_visits* *Other response}* clinic?

_____ hours _____ minutes

*other_wait* How much time did you spend at the {*spec_visits* *Other response}* clinic (including wait and visit times)?

_____ hours _____ minutes

*other_last was displayed if the other_num response was greater than one.*
*other_last* When was the last time you went to the {*spec_visits* *Other response}* clinic for your most recent COVID-19?

| Month | Year |
|---|---|
| ▼ January (1 ... December (12) | ▼ 2020 (1 ... 2023 (4) |

[*A pulldown menu was displayed with the months in chronological order, and a pulldown menu was displayed with the years in chronological order*.]

*Now we are going to ask you about out-of-pocket costs you might have had related to your first {spec_visits Other response} clinic visit when you most recently had COVID-19.*

***Out-of-pocket costs** are payments you made that were not reimbursed (paid back) by an insurance company.*

*If you don't know or can't remember the exact details, please provide your best answer possible based on what you can remember.*

*other_direct* How much did you pay out-of-pocket for direct medical costs - such as visit fees, copays, deductibles, lab fees, x-ray - related to the first time you went to the {*spec_visits* *Other response}* clinic for your most recent COVID-19? *Round to the nearest $10 value.*

________________________________________________________________

*other_non* How much did you pay for non-medical costs - such as gas, meals, hotel stays, and other similar costs - related to the first time you went to the {*spec_visits* *Other response}* clinic for your most recent COVID-19? *Round to the nearest $10 value.*

________________________________________________________________

*other_desc* Please describe any non-medical costs you had. *Type "None" if you did not have any non-medical costs.*

________________________________________________________________

*Page Break*

*In Patient Rehab Block was displayed if the covid_provider response was* "Yes" *to* "In-Patient Rehabilitation Clinic".

**Inpatient Rehab Block**

*Now we are going to ask you about inpatient rehabilitation you experienced when you most recently had COVID-19, which could include care for long-term symptoms (Long COVID).*

*Answer the following questions based on when you most recently had COVID-19.*

*inpatient_num* How many times were you admitted to an inpatient rehabilitation clinic as a result of your most recent COVID-19?

________________________________________________________________

*Page Break*

*Answer the following questions based on when you most recently had COVID-19. If you can't remember the exact details, please provide your best answer possible based on what you can remember.*

*inpatient_firstdate* When was the first time you were admitted to an inpatient rehabilitation clinic as a result of your most recent COVID-19?

| Month | Year |
| --- | --- |
| ▼ January (1 ... December (12) | ▼ 2020 (1 ... 2023 (4) |

[*A pulldown menu was displayed with the months in chronological order, and a pulldown menu was displayed with the years in chronological order*.]

*inpatient_firstdays* How many days were you at the inpatient rehabilitation clinic?

________________________________________________________________

*inpatient_lastdate was displayed if the inpatient_num was greater than one.*

*inpatient_lastdate* When was the last time you were admitted to an inpatient rehabilitation clinic as a result of your most recent COVID-19?

| Month | Year |
| --- | --- |
| ▼ January (1 ... December (12) | ▼ 2020 (1 ... 2023 (4) |

[*A pulldown menu was displayed with the months in chronological order, and a pulldown menu was displayed with the years in chronological order*.]

*inpatient_lastdays was displayed if the inpatient_num was greater than one.*

*inpatient_lastdays* How many days were you at the inpatient rehabilitation clinic?

________________________________________________________________

*Now we are going to ask you about out-of-pocket costs you might have had related to the first time you were admitted to an inpatient rehabilitation clinic as a result of your most recent COVID-19.*

***Out-of-pocket costs** are payments you made that were not reimbursed (paid back) by an insurance company.*

*If you can't remember the exact details, please provide your best answer possible based on what you can remember.*

*inpatient_direct* How much did you pay out-of-pocket for direct medical costs - such as visit fees, copays, deductibles, lab fees, x-ray - related to the first time you were admitted to an inpatient rehabilitation clinic as a result of your most recent COVID-19? *Round to the nearest $10 value.*

________________________________________________________________

*inpatient_non* How much did you pay for non-medical costs - such as gas, meals, hotel stays, and other similar costs - related to the first time you were admitted to an inpatient rehabilitation clinic as a result of your most recent COVID-19? *Round to the nearest $10 value.*

________________________________________________________________

*inpatient_desc* Please describe any non-medical costs you had. *Type "None" if you did not have any non-medical costs.*

________________________________________________________________

*Page Break*

*Home Health Care Services Block was displayed if the covid_home response was* "Yes".

**Home Health Care Services Block**

*Now we are going to ask you about home health care services you received related to your most recent COVID-19, which could include care for long-term symptoms (Long COVID). If you can't remember the exact details, please provide your best answer possible based on what you can remember.*

*home_first* When did you first start receiving home health care services (example: in-home rehabilitation) as a result of your most recent COVID-19?

| Month | Year |
|---|---|
| ▼ January (1 ... December (12) | ▼ 2020 (1 ... 2023 (4) |

[*A pulldown menu was displayed with the months in chronological order, and a pulldown menu was displayed with the years in chronological order*.]

*home_last* When was the last time you received home health care services as a result of your most recent COVID-19?

| Month | Year |
| --- | --- |
| ▼ January (1 ... December (12) | ▼ 2020 (1 ... 2023 (4) |

[*A pulldown menu was displayed with the months in chronological order, and a pulldown menu was displayed with the years in chronological order*.]

*home_days* How many days per week {*LongCovid*} you receive home health care services on average as a result of your most recent COVID-19?

________________________________________________________________

*home_hours* How many hours per day {*LongCovid*} you receive home health care services on average as a result of your most recent COVID-19?

________________________________________________________________

*Page Break*

*Now we are going to ask you about out-of-pocket costs you might have had related to home health care services you received as a result of your most recent COVID-19.*

***Out-of-pocket costs** are payments you made that were not reimbursed (paid back) by an insurance company.*

*If you can't remember the exact details, please provide your best answer possible based on what you can remember.*

*home_direct* How much did you pay out-of-pocket for direct medical costs - such as visit fees, equipment, copays, deductibles - related to home health care services you received as a result of your most recent COVID-19? *Round to the nearest $10 value.*

________________________________________________________________

*Page Break*

*Medications Block was displayed if the covid_meds response was* “Yes”.

**Medications Block**

*Now we are going to ask you about medicines you received related to your {CovidStatus} COVID-19, which could include medicines for long-term symptoms (Long COVID). If you can't remember the exact details, please provide your best answer possible based on what you can remember.*

*meds_prescription* How many types of prescription medicines did you take for you most recent COVID-19?

________________________________________________________________

*meds_pcost* How much did you pay out-of-pocket for these prescription medicines? *Round to the nearest $10 value.*

________________________________________________________________

*meds_otc* How many types of over-the-counter (non-prescription) medicines did you take for you most recent COVID-19?

________________________________________________________________

*meds_otccost* How much did you pay out-of-pocket for these over-the-counter medicines? *Round to the nearest $10 value.*

________________________________________________________________

*Page Break*

**Insurance and Reimbursement Block**

*Now we are going to ask you about any health insurance you may have had during {CovidStatus} COVID-19, which could include care for long-term symptoms (Long COVID).*

*ins_coverage* Did you have any health insurance or health coverage plans during your most recent COVID-19?

- ○ No (2)
- ○ Yes (1)
- ○ Unsure/I can't remember (3)

*Participants skipped to the Confidence Estimates Block if the ins_coverage response was NOT* “Yes”.

<u>ins_type</u> Were you covered by any of the following types of health insurance or health coverage plans during your most recent COVID-19? *Mark "No" or "Yes" for EACH type of insurance.*

| | No (1) | Yes (2) |
|---|---|---|
| Private insurance through a <u>current</u> or former employer or union (of you or another family member) (1) | ○ | ○ |
| Insurance purchased directly from an insurance company (2) | ○ | ○ |
| Insurance purchased directly from HealthCare.gov or a state Health Insurance Marketplace (3) | ○ | ○ |
| Medicare (for people 65 and older, or people with certain disabilities) (4) | ○ | ○ |
| Medicaid, Medical Assistance, or any kind of government-assistance plan for people with low incomes or a disability (5) | ○ | ○ |
| TRICARE or other military health care (6) | ○ | ○ |
| VA (7) | ○ | ○ |
| Indian Health Service (8) | ○ | ○ |
| Any other type of health insurance or health coverage plan - Specify: (9) | ○ | ○ |

<u>ins_reimburse</u> Were you partially or fully reimbursed (paid back) by a health insurance company or public assistance program for any payments you made for care, services, or medicines you received related to your <u>most recent</u> COVID-19?

- ○ No (1)
- ○ Yes (2)
- ○ Waiting to find out (3)
- ○ Don't know/Unsure (4)

*Participants skipped to the Confidence Estimates Block if the <u>ins_reimburse</u> response was NOT "Yes".*

<u>ins_amount</u> *If you can't remember the exact details for the following questions, please provide your best answer possible based on what you can remember.*

How much money were you reimbursed (paid back) by a health insurance company or public assistance program for any payments you made for care, services, or medicines you received related to your most recent COVID-19? *Round to the nearest $10 value.*

________________________________________________________________

*Page Break*

*ins_time* How much time did you spend talking to your insurance company about reimbursements or other issues related to your most recent COVID-19?

_____ hours _____ minutes

*Page Break*

**Confidence Estimates Block**
*Now we are going to ask you about how confident you were with some of the answers you provided in the previous sections.*

*conf_medical* Overall, how confident are you that the numbers you provided about medical costs are accurate?

| 1 | 2 | 3 | 4 | 5 |
|---|---|---|---|---|
| Not at all confident | | | | Very confident |

*conf_non* Overall, how confident are you that the numbers you provided about non-medical costs are accurate?

| 1 | 2 | 3 | 4 | 5 |
|---|---|---|---|---|
| Not at all confident | | | | Very confident |

*conf_travel* Overall, how confident are you that the numbers you provided about travel time are accurate?

| 1 | 2 | 3 | 4 | 5 |
|---|---|---|---|---|
| Not at all confident | | | | Very confident |

*conf_wait* Overall, how confident are you that the numbers you provided about time at the clinic (wait and visit times) are accurate?

| 1 | 2 | 3 | 4 | 5 |
|---|---|---|---|---|
| Not at all confident | | | | Very confident |

*Page Break*

**Financial Challenges Block**
*Now we are going to ask you about any challenges you may have had during most recent COVID-19 or as a result of it, which could include care for long-term symptoms (Long COVID).*

*Answer the following questions based on when you most recently had COVID-19.*

*bills_problem* Did you or anyone else in your household have problems paying, or were not able to pay, any COVID-related medical bills? *Select all that apply.*

- ☐ ⊗No (1)
- ☐ Yes, when I had COVID (2)
- ☐ Yes, after I had COVID (3)
- ☐ Yes, currently (4)

*bills_wait* Did you or anyone else in your household ever wait to get any medical care (COVID-related or otherwise) because of worry about the cost?

- ☐ ⊗No (1)
- ☐ Yes, when I had COVID (2)
- ☐ Yes, after I had COVID (3)
- ☐ Yes, currently (4)

*bills_meds* Did you or anyone else in your household ever take less or skip doses of medicine (COVID-related or otherwise) because of worry about the cost?

- ☐ ⊗No (1)
- ☐ Yes, when I had COVID (2)
- ☐ Yes, after I had COVID (3)
- ☐ Yes, currently (4)

*bills_care* Was there any time you or anyone else in your household needed medical care (COVID-related or otherwise) but did not get it because of worry about the cost?

- ☐ ⊗No (1)
- ☐ Yes, when I had COVID (2)
- ☐ Yes, after I had COVID (3)
- ☐ Yes, currently (4)

*bills_prescription* Was there any time you or anyone else in your household needed a prescription medicine (COVID-related or otherwise) but did not get it because of worry about the cost?

- ☐ ⊗No (1)
- ☐ Yes, when I had COVID (2)
- ☐ Yes, after I had COVID (3)
- ☐ Yes, currently (4)

*Page Break*

*bills_other* Did you or anyone else in your household have problems paying, or were not able to pay, any non-medical bills (examples: mortgage/rent; student loans; car loan; credit cards)? *Select all that apply.*

- ☐ ⊗No (1)
- ☐ Yes, when I had COVID (2)
- ☐ Yes, after I had COVID (3)
- ☐ Yes, currently (4)

*bills_meals* Did you or anyone else in your household ever not eat as much, or skip meals, because there wasn't enough money for food? *Select all that apply.*

- ☐ ⊗No (1)
- ☐ Yes, when I had COVID (2)
- ☐ Yes, after I had COVID (3)
- ☐ Yes, currently (4)

*covid_absent* How many days of school or work did you miss as a result of your most recent COVID-19?

________________________________________________________________

*Page Break*

**Vaccine History and Attitudes Block**
*Almost done! Now we are going to ask you about vaccines you may have received, and for some additional information about you.*

*self_flu* Over the last 3 years, how many times did you get an annual flu vaccine?

Never (1) Sometimes (2) Every year (3)

*self_covid* Have you ever received a COVID-19 vaccine?

No (2) Yes (1)

*self_booster* Have you received a COVID-19 booster shot since September 1, 2022?

No (2) Yes (1)

*vax_attitudes* How do you feel about {*vaxgroup*}, in general?

| 1 | 2 | 3 | 4 | 5 |
|---|---|---|---|---|
| Very negative | | | | Very positive |

*Page Break*

**Additional Demographics Block**
*state* Which state do you live in?

[*U.S. states and territories were listed in alphabetical order, with* District of Columbia *coded as* “9”, and “40” was skipped (i.e., Rhode Island *was coded as* “41” *instead of* “40”.]

▼ Alabama (1) ... Wyoming (51)

*rurality* How would you describe the area in which you live?

Urban (1) Suburban (2) Rural (3)

*educ* What is the highest level of schooling you have completed?
- Less than high school (1)
- High school (Diploma or GED) (2)
- Bachelor's degree (BS, BA, etc.) (3)
- Master's degree (MA, MPH, etc.) (4)
- Doctoral/Professional degree (PhD, MD, etc.) (5)

*income* Which of the following categories best describes your total household income, from all sources, before taxes?

- ○ Less than $20,000 (1)
- ○ $20,000 - $39,999 (2)
- ○ $40,000 - $59,999 (3)
- ○ $60,000 - $79,999 (4)
- ○ $80,000 - $99,999 (5)
- ○ $100,000 - $149,999 (6)
- ○ $150,000 - $199,999 (7)
- ○ $200,000 - $249,999 (8)
- ○ $250,000 - $299,999 (9)
- ○ $300,000 or more (10)
- ○ Prefer not to answer (11)

*income_people* Including yourself, how many people does that income support?

________________________________________________________________

*Page Break*

*health_conditions* Have you been diagnosed with any of the following conditions? *Select all that apply.*

- ▢ Chronic obstructive pulmonary disease (COPD) (1)
- ▢ Heart disease (2)
- ▢ Diabetes (3)
- ▢ Immunocompromised condition or taking medications that suppress the immune system (4)
- ▢ Kidney disease or need for dialysis (5)
- ▢ Liver disease (6)
- ▢ ⊗None of these (7)

*comments* Do you have any other comments or thoughts you would like to share with us regarding challenges you’ve experienced as a result of having COVID-19?

________________________________________________________________

### COVID-19 Caregiver Burden Caregiver Survey

***NOTES FOR READER***

- **BLOCK TITLES**: Labels for the question blocks used by the study team but not displayed in the survey.
- **VARIABLE NAMES**: Text in italics and underlined at the beginning of a question that were not displayed in the survey but exported in the data file.
- **RESPONSE VALUES**: Numbers in parentheses at the end of response options indicate the numerically coded response in the data file but were not displayed in the survey.
- **SURVEY LOGIC**: Conditions used to determine survey flow are italicized and highlighted with a specific color to indicate the type of survey flow (see below). Survey logic is included for the reader's benefit and was not displayed in the survey.
    - Red = Screen out logic
        - Respondent routed back to online panel website.
    - Green = Display logic
        - Survey question is displayed if certain conditions are met.
    - Orange = Skip logic
        - Respondents skip certain questions if certain conditions are met.
    - Gray = Page break
    - Italicized text in {*braces*} indicates text that was "piped" in text of a prior response or text that varied depending on a prior response.
- **OTHER NOTES**: Italicized text in [*brackets*] are other notes for the reader and were not displayed in the survey.
- **PIPED TEXT**
    - *LongCovid*: "Did" (or "did") OR "Does" (or "does")
    - *CovidStatus*: "most recent" OR "current"
    - care_person: Lowercase response option for care_person item
    - vax_group: "vaccines" OR "vaccination"

**Introduction Block**
*We invite you to be in a research study being conducted by investigators from The University of Iowa with funding from the Centers for Disease Control and Prevention. The purpose of the study is to ask adults about important immunization-related issues to inform recommendations and policies.*

*We are inviting you to be in this study because Qualtrics has identified you as someone who is aged 18 years or older. Approximately 1800 respondents will take part in this online survey.*

*If you agree to participate, we would like you to complete a survey regarding your experiences caring for someone who has had COVID-19. Other than questions to determine your eligibility for the study, you are free to skip any questions that you prefer not to answer. It will take approximately 15 minutes to complete the survey.*

*We will keep the information you provide confidential, however federal regulatory agencies and the University of Iowa Institutional Review Board (a committee that reviews and approves research studies) may inspect and copy records pertaining to this research. We will not collect your name or information that can be used to identify you.*

*There are no known risks from being in this study, and you will not benefit personally.  However, we hope that others may benefit in the future from what we learn because of this study.*

*There are no costs for being in this study and being in this study is up to you. If you do not wish to be in the study, you may close the browser window now or at any time before submitting the survey. Submitting the survey will indicate your willingness to be in the study. You will be compensated according to the terms of your panel agreement.*

*If you have any questions about the study itself or experience a research-related injury, please contact Dr. Aaron Scherer ( or 319.384.6180). If you have questions about the rights of research subjects, please contact the Human Subjects Office, 105 Hardin Library for the Health Sciences, 600 Newton Rd, The University of Iowa, Iowa City, IA  52242-1098, (319) 335-6564, or. To offer input about your experiences as a research subject or to speak to someone other than the research staff, call the Human Subjects Office at the number above.*

*Thank you very much for your consideration.*

*Sincerely,*

*Aaron Scherer, PhD*
*Assistant Professor of Internal Medicine*

*Page Break*

*consent* Please only click 'Yes' to indicate that you have read and understand the above information, are fully aware of the risks, your right to withdraw at any point, and desire of your own free will to participate in this research study.

- No (2)
- Yes (1)

*Participants screened out of survey if consent response was "No".*

*Page Break*

*quality We care about the quality of our data. For us to get the most accurate measures of your opinions, it is important that you thoughtfully provide your best answers to each question in this survey.*

Do you commit to thoughtfully provide your best answers to each question in this survey?

- I will provide my best answers (1)
- I will not provide my best answers (2)
- I can't promise either way (3)

*Participants screened out of survey if quality response was* "I will not provide my best answers" *or* "I can't promise either way".

*Page Break*

**Demographics Block**

*age* What is your age?

________________________________________________________________

*gender* Do you currently describe yourself as male, female, or transgender?

- ○ Male (1)
- ○ Female (2)
- ○ Transgender (3)
- ○ None of these (4)
- ○ Prefer not to say (5)

*ethnic* Do you consider yourself to be Hispanic or Latino/a?

- ○ Hispanic or Latino/a (1)
- ○ Not Hispanic or Latino/a (2)
- ○ None of these (3)
- ○ Prefer not to answer (4)

*race* What race or races do you consider yourself to be? *Select all that apply.*

- ▢ American Indian or Alaskan Native (1)
- ▢ Asian (2)
- ▢ Black or African American (3)
- ▢ Native Hawaiian or Other Pacific Islander (4)
- ▢ White (5)
- ▢ ⊗None of these (6)
- ▢ ⊗Prefer not to answer (7)

*covid_status* Have you ever been the primary caregiver of a friend or family member while they were sick with COVID-19? Do not include caregiving for someone with COVID-19 that you were paid to do (example: it was part of your job responsibilities).

- ○ No (1)
- ○ Yes (2)
- ○ Unsure (3)
- ○ Prefer not to say (4)

*Page Break*

**Caregiver Intro Block**

*care_num* How many friends or family members have you cared for who have been sick with COVID-19?

________________________________________________________________

*care_current* Are you currently taking care of a friend or family member who is sick with COVID-19?

No (2) Yes (1)

*Page Break*

*care_age* How old was the family member you most recently cared for who was sick with COVID-19?

________________________________________________________________

*care_person* How is the person you cared for related to you?

*They are my . . .*

- ○ Child (1)
- ○ Spouse/Partner (2)
- ○ Parent (3)
- ○ Grandparent (4)
- ○ Sibling (5)
- ○ Niece (6)
- ○ Nephew (7)
- ○ Aunt (8)
- ○ Uncle (9)
- ○ Other *(please write-in relationship)*: (10) ________________________________________________

*Page Break*

**Caregiving Information Block**
*care_severe* How severe were your *{care_person}*'s COVID-19 symptoms at their worst? Symptoms resulting in hospitalization or being admitted to the ICU would be considered "very severe."

| 1 | 2 | 3 | 4 | 5 | 6 |
|---|---|---|---|---|---|
| No symptoms | | | | | Very severe symptoms |

*care_current* Does your *{care_person}* currently have any COVID-19 symptoms?

No (2) Yes (1) Don’t know/Unsure (3)

*Page Break*

**Caregiver Long COVID Block**

*care_long* {*LongCovid*} your *{care_person}* have any symptoms that lasted 3 months or longer that they did not have prior to when they *most recently had* COVID-19 (in other words, *Long COVID symptoms*)?

*Long COVID symptoms may include: tiredness or fatigue, difficulty thinking or concentrating, forgetfulness, or memory problems (sometimes referred to as "brain fog"), difficulty breathing or shortness of breath, joint or muscle pain, fast-beating or pounding heart (also known as heart palpitations), chest pain, dizziness on standing, changes to your menstrual cycle, changes to taste/smell, or inability to exercise.*

- No (2)
- Yes (1)
- Unsure (3)

*Page Break*

*long_severe was displayed if the care_long response was* "Yes".

*long_severe* How much {lowercase *LongCovid*} these Long COVID symptoms reduce your *{care_person}*'s ability to carry out day-to-day activities compared with the time before they had COVID-19?

| 1 | 2 | 3 | 4 | 5 | 6 |
|---|---|---|---|---|---|
| Not at all | | | | | A lot |

*Page Break*

**Most Recent COVID Caregiving Block**
*Answer the following questions based on your most recent experience caring for your {care_person} when they were most recently sick with COVID-19, which could include care for long-term symptoms (Long COVID).*

*covid_provider* When your *{care_person}* was most recently sick with COVID-19, did you go with them to be seen by a healthcare provider, such as a doctor or nurse, at any of these locations (including telehealth/virtual visits) for their COVID-19?

| | No (1) | Yes (2) |
|---|---|---|
| **Primary Care Clinic** (1) | ○ | ○ |
| **Urgent Care Clinic** (2) | ○ | ○ |
| **Emergency Room (ER)** (3) | ○ | ○ |
| **Specialty Clinic** Examples: Pulmonology (lung) or cardiology (heart) clinic (4) | ○ | ○ |
| **In-Patient Rehabilitation Clinic** (5) | ○ | ○ |

*covid_hosp* Was your *{care_person}* hospitalized as a result of their most recent COVID-19?

No (2) Yes (1)

*covid_home* Did your *{care_person}* receive any home health care services (example: in-home rehabilitation) as a result of their most recent COVID-19?

No (2) Yes (1)

*covid_meds* Did your *{care_person}* take any over-the-counter (non-prescription) or prescription medicines to treat, or as a result of, their COVID-19? *(Examples: cold/cough medicine, decongestants, fever/pain reducers, inhalers or nebulizers, antibiotics for complications of COVID-19. Do not include medications taken during a hospitalization.)*

No (2) Yes (1)

*Page Break*

*Primary Care Visits Block was displayed if the covid_provider response was* "Yes" *to* "Primary Care Clinic".

**Primary Care Visits Block**

*Now we are going to ask you about primary care (non-specialist) visits your {care_person} had for their most recent COVID, which could include care for long-term symptoms (Long COVID). If you don't know or can't remember the exact details, please provide your best answer possible based on what you can remember.*

*primary_num* How many times did your *{care_person}* visit a primary care (non-specialist) clinic (including telehealth/virtual visits) for their most recent COVID-19?

________________________________________________________________

*primary_virtual* How many of these visits were virtual?

________________________________________________________________

*Page Break*

*Answer the following questions based on when you cared for your {care_person} for their most recent COVID-19. If you don't know or can't remember the exact details, please provide your best answer possible based on what you can remember.*

*primary_date* When was the first time you went with your *{care_person}* to a primary care (non-specialist) clinic for their most recent COVID-19?

| Month | Year |
| --- | --- |
| ▼ January (1 ... December (12) | ▼ 2020 (1 ... 2023 (4) |

[*A pulldown menu was displayed with the months in chronological order, and a pulldown menu was displayed with the years in chronological order*.]

*primary_travel* How long did it take to travel to the clinic?

_____ hours _____ minutes

*primary_wait* How much time did you spend at the clinic (including wait and visit times)?

_____ hours _____ minutes

*Page Break*

**Primary Care Part 2 Block**

*Display This Question:*

*If primary_num > primary_virtual*

primary_last When was the last time you went with your *{care_person}* to a primary care (non-specialist) clinic for their most recent COVID-19?

| Month | Year |
|---|---|
| ▼ January (1 ... December (12) | ▼ 2020 (1 ... 2023 (4) |

*Page Break*

*Now we are going to ask you about out-of-pocket costs you might have had related to your first visit with your {care_person} to the primary care clinic for their most recent COVID-19.*

***Out-of-pocket costs** are payments you made that were not reimbursed (paid back) by an insurance company.*

*If you don't know or can't remember the exact details, please provide your best answer possible based on what you can remember.*

*primary_direct was displayed if the care_person response was* "Child".

*primary_direct* How much did you pay out-of-pocket for direct medical costs - such as visit fees, copays, deductibles, lab fees, x-ray - related to the first time your child went to a primary care clinic for their most recent COVID-19? *Round to the nearest $10 value.*

________________________________________________________________

*primary_non* How much did you pay for non-medical costs - such as gas, meals, hotel stays, and other similar costs - related to the first time you went with your *{care_person}* to a primary care clinic for their most recent COVID-19? *Round to the nearest $10 value.*

________________________________________________________________

*primary_desc* Please describe any non-medical costs you had. *Type "None" if you did not have any non-medical costs.*

________________________________________________________________

*Page Break*

*Urgent Care Visits Block was displayed if the covid_provider response was* “Yes” *to* “Urgent Care Clinic”.

**Urgent Care Visits Block**

*Now we are going to ask you about urgent care visits your {care_person} had for their most recent COVID, which could include care for long-term symptoms (Long COVID). If you don't know or can't remember the exact details, please provide your best answer possible based on what you can remember.*

*Page Break*

*urgent_num* How many times did your *{care_person}* visit an urgent care clinic for their most recent COVID-19?

________________________________________________________________

*Page Break*

*Answer the following questions based on when you cared for your {care_person} for their most recent COVID-19. If you don't know or can't remember the exact details, please provide your best answer possible based on what you can remember.*

*urgent_date* When was the first time you went with your *{care_person}* to an urgent care clinic for their most recent COVID-19?

| Month | Year |
| --- | --- |
| ▼ January (1 ... December (12) | ▼ 2020 (1 ... 2023 (4) |

[*A pulldown menu was displayed with the months in chronological order, and a pulldown menu was displayed with the years in chronological order*.]

*urgent_travel* How long did it take to travel to the clinic?

_____ hours _____ minutes

*urgent_wait* How much time did you spend at the clinic (including wait and visit times)?

_____ hours _____ minutes

*Page Break*

*urgent_last was displayed if the urgent_num response was greater than one.*

*urgent_last* When was the last time you went with your *{care_person}* to an urgent care clinic for their most recent COVID-19?

| Month | Year |
|---|---|
| ▼ January (1 ... December (12) | ▼ 2020 (1 ... 2023 (4) |

[*A pulldown menu was displayed with the months in chronological order, and a pulldown menu was displayed with the years in chronological order*.]

*Page Break*

*Now we are going to ask you about out-of-pocket costs you might have had related to your first visit with your {care_person} to an urgent care clinic for their most recent COVID-19.*

***Out-of-pocket costs** are payments you made that were not reimbursed (paid back) by an insurance company.*

*If you don't know or can't remember the exact details, please provide your best answer possible based on what you can remember.*

*urgent_direct was displayed if the care_person response was* "Child".

*urgent_direct* How much did you pay out-of-pocket for direct medical costs - such as visit fees, copays, deductibles, lab fees, x-ray - related to the first time your child went to an urgent care clinic for their most recent COVID-19? *Round to the nearest $10 value.*

________________________________________________________________

*urgent_non* How much did you pay for non-medical costs - such as gas, meals, hotel stays, and other similar costs - related to the first time you went with your *{care_person}* to an urgent care clinic for their most recent COVID-19? *Round to the nearest $10 value.*

________________________________________________________________

*urgent_desc* Please describe any non-medical costs you had. *Type "None" if you did not have any non-medical costs.*

________________________________________________________________

*Page Break*

*Hospitalizations Block was displayed if the covid_hosp response was* "Yes".

**Hospitalizations Block**

*Now we are going to ask you about hospitalizations your {care_person} had when they most recently had COVID-19, which could include hospitalizations related to long-term symptoms (Long COVID).*

*hosp_num* How many times was your *{care_person}* hospitalized because of their most recent COVID-19?

________________________________________________________________

*Page Break*

*Answer the following questions based on when your {care_person} most recently had COVID-19. If you don't know or can't remember the exact details, please provide your best answer possible based on what you can remember.*

*hosp_date* When was the first time your *{care_person}* was hospitalized for their most recent COVID-19?

| Month | Year |
| --- | --- |
| ▼ January (1 ... December (12) | ▼ 2020 (1 ... 2023 (4) |

[*A pulldown menu was displayed with the months in chronological order, and a pulldown menu was displayed with the years in chronological order*.]

*hosp_days* How many days did they stay at the hospital?

________________________________________________________________

*Page Break*

*hosp_last was displayed if the hosp_num response was greater than 1.*
*hosp_last* When was the last time your *{care_person}* was hospitalized for their most recent COVID-19?

| Month | Year |
| --- | --- |
| ▼ January (1 ... December (12) | ▼ 2020 (1 ... 2023 (4) |

[*A pulldown menu was displayed with the months in chronological order, and a pulldown menu was displayed with the years in chronological order*.]

*hosp_ldays* How many days did they stay at the hospital?

________________________________________________________________

*Page Break*

*Now we are going to ask you about out-of-pocket costs you might have had related to the first time your {care_person} was hospitalized as a result of their most recent* COVID-19.

***Out-of-pocket costs** are payments you made that were not reimbursed (paid back) by an insurance company.*

*If you don't know or can't remember the exact details, please provide your best answer possible based on what you can remember.*

*hosp_direct was displayed if the care_person response was* "Child".
*hosp_direct* How much did you pay out-of-pocket for direct medical costs - such as visit fees, copays, deductibles, lab fees, x-ray - related to the first time your child was hospitalized for their most recent COVID-19? *Round to the nearest $10 value.*

________________________________________________________________

*hosp_non* How much did you pay for non-medical costs - such as gas, meals, hotel stays, and other similar costs - related to the first time your *{care_person}* was hospitalized for their most recent COVID-19? *Round to the nearest $10 value.*

________________________________________________________________

*hosp_desc* Please describe any non-medical costs you had. *Type "None" if you did not have any non-medical costs.*

________________________________________________________________

*Page Break*

*Emergency Room Visits Block was displayed if the covid_provider response was "Yes" to* "Emergency Room (ER)".

**Emergency Room Visits Block**

*Now we are going to ask you about emergency room (ER) visits your {care_person} had for their most recent COVID, which could include care for long-term symptoms (Long COVID).*

*er_num* How many times did your *{care_person}* go to an emergency room (ER) for their most recent COVID-19?

________________________________________________________________

*Page Break*

*Answer the following questions based on when your {care_person} most recently had COVID-19. If you don't know or can't remember the exact details, please provide your best answer possible based on what you can remember.*

*er_date* When was the first time you went with your *{care_person}* to an emergency room for their most recent COVID-19?

| Month | Year |
|---|---|
| ▼ January (1 ... December (12) | ▼ 2020 (1 ... 2023 (4) |

[*A pulldown menu was displayed with the months in chronological order, and a pulldown menu was displayed with the years in chronological order*.]

*er_travel* How long did it take you to travel to the emergency room?

_____ hours _____ minutes

*er_wait* How much time did you spend at the emergency room (including wait and visit times)?

_____ hours _____ minutes

*Page Break*

*er_last was displayed if the er_num response was greater than 1.*

er_last When was the last time you went with your *{care_person}* to an emergency room for their most recent COVID-19?

| Month | Year |
| --- | --- |
| ▼ January (1 ... December (12) | ▼ 2020 (1 ... 2023 (4) |

[*A pulldown menu was displayed with the months in chronological order, and a pulldown menu was displayed with the years in chronological order*.]

*Page Break*

*Now we are going to ask you about out-of-pocket costs you might have had related to your {care_person}'s first emergency room visit when they most recently had COVID-19.*

***Out-of-pocket costs** are payments you made that were not reimbursed (paid back) by an insurance company.*

*If you don't know or can't remember the exact details, please provide your best answer possible based on what you can remember.*

*er_direct was displayed if the care_person response was* "Child".

er_direct How much did you pay out-of-pocket for direct medical costs - such as visit fees, copays, deductibles, lab fees, x-ray - related to the first time your child went to the emergency room for their most recent COVID-19? *Round to the nearest $10 value.*

________________________________________________________________

er_non How much did you pay for non-medical costs - such as gas, meals, hotel stays, and other similar costs - related to the first time you went with your *{care_person}* to the emergency room for their most recent COVID-19? *Round to the nearest $10 value.*

________________________________________________________________

er_desc Please describe any non-medical costs you had. *Type "None" if you did not have any non-medical costs.*

________________________________________________________________

*Page Break*

*Medical Specialist Visits Block was displayed if the covid_provider response was* "Yes" *to* "Specialty Clinic Examples: Pulmonology (lung) or cardiology (heart) clinic".

**Medical Specialist Visits Block**

*Now we are going to ask you about visits to a specialty care clinic (examples: pulmonology (lung) or cardiology (heart) clinic) your {care_person} had when they most recently had COVID-19, which could include care for long-term symptoms (Long COVID).*

*spec_type* Did your *{care_person}* have any outpatient visits with any of the following specialists for their most recent COVID-19? *Select all that apply.*

- ☐ Infectious disease specialist (1)
- ☐ Pulmonologist (lung doctor) (2)
- ☐ Cardiologist (heart doctor) (3)
- ☐ Oncologist (cancer doctor) (4)
- ☐ Obstetrician/Gynecologist (OB/GYN) (5)
- ☐ Physical therapist (6)
- ☐ Occupational therapist (7)
- ☐ Other - please specify: (8) ________________________________________________
- ☐ ⊗None of these (9)

*Page Break*

*Infectious Disease Intro Block and Infectious Disease Visits Block were displayed if the spec_type response was* "Infectious disease specialist".

**Infectious Disease Intro Block**

*Answer the following questions based on when your {care_person} most recently had COVID-19.*

*id_num* How many times did your *{care_person}* visit an infectious disease specialist clinic (including telehealth/virtual visits) for their most recent COVID-19?

________________________________________________________________

*id_virtual* How many of these visits were virtual?

________________________________________________________________

*Page Break*

**Infectious Disease Visits Block**

*Answer the following questions based on when your {care_person} most recently had COVID-19. If you don't know or can't remember the exact details, please provide your best answer possible based on what you can remember.*

*id_date* When was the first time you went with your *{care_person}* to an infectious disease specialist clinic for their most recent COVID-19?

| Month | Year |
|---|---|
| ▼ January (1 ... December (12) | ▼ 2020 (1 ... 2023 (4) |

[*A pulldown menu was displayed with the months in chronological order, and a pulldown menu was displayed with the years in chronological order*.]

*id_travel* How long did it take you to travel to the infectious disease specialist clinic?

_____ hours _____ minutes

*id_wait* How much time did you spend at the infectious disease specialist clinic (including wait and visit times)?

_____ hours _____ minutes

*Page Break*

*Display This Question:*
*If id_num > id_virtual*

*id_last* When was the last time you went with your *{care_person}* to an infectious disease specialist clinic for their most recent COVID-19?

| | Month | Year |
|---|---|---|
| (1) | ▼ January (1 ... December (12) | ▼ 2020 (1 ... 2023 (4) |

*Page Break*

*Now we are going to ask you about out-of-pocket costs you might have had related to your {care_person}'s first infectious disease specialist clinic visit when they most recently had COVID-19.*

***Out-of-pocket costs** are payments you made that were not reimbursed (paid back) by an insurance company.*

*If you don't know or can't remember the exact details, please provide your best answer possible based on what you can remember.*

*id_direct was displayed if the care_person response was* "Child".
*id_direct* How much did you pay out-of-pocket for direct medical costs - such as visit fees, copays, deductibles, lab fees, x-ray - related to the first time your child went to the infectious disease specialist clinic for their most recent COVID-19? *Round to the nearest $10 value.*

________________________________________________________________

*id_non* How much did you pay for non-medical costs - such as gas, meals, hotel stays, and other similar costs - related to the first time you went with your *{care_person}* to the infectious disease specialist clinic for their most recent COVID-19? *Round to the nearest $10 value.*

________________________________________________________________

*id_desc* Please describe any non-medical costs you had. *Type "None" if you did not have any non-medical costs.*

________________________________________________________________

*Page Break*

*Pulmonology Intro Block and Pulmonology Visits Block were displayed if the spec_type response was* "Pulmonologist (lung doctor)".

**Pulmonology Intro Block**

*Answer the following questions based on when your {care_person} most recently had COVID-19.*

*pulm_num* How many times did your *{care_person}* visit a pulmonology clinic (including telehealth/virtual visits) for their most recent COVID-19?

________________________________________________________________

*pulm_virtual* How many of these visits were virtual?

________________________________________________________________

*Page Break*

**Pulmonology Visits Block**
*Answer the following questions based on when your {care_person} most recently had COVID-19. If you can't remember the exact details, please provide your best answer possible based on what you can remember.*

*pulm_date* When was the first time you went with your *{care_person}* to a pulmonology clinic for their most recent COVID-19?

| Month | Year |
|---|---|
| ▼ January (1 ... December (12) | ▼ 2020 (1 ... 2023 (4) |

[*A pulldown menu was displayed with the months in chronological order, and a pulldown menu was displayed with the years in chronological order*.]

*pulm_travel* How long did it take you to travel to the pulmonology clinic?

_____ hours _____ minutes

*pulm_wait* How much time did you spend at the pulmonology clinic (including wait and visit times)?

_____ hours _____ minutes

*Page Break*

*Display This Question:*
*If pulm_num > pulm_virtual*

*pulm_last* When was the last time you went with your *{care_person}* to a pulmonology clinic for their most recent COVID-19?

| Month | Year |
|---|---|
| ▼ January (1 ... December (12) | ▼ 2020 (1 ... 2023 (4) |

[*A pulldown menu was displayed with the months in chronological order, and a pulldown menu was displayed with the years in chronological order*.]

*Page Break*

*Now we are going to ask you about out-of-pocket costs you might have had related to your {care_person}'s first pulmonology clinic visit when they most recently had COVID-19.*

***Out-of-pocket costs** are payments you made that were not reimbursed (paid back) by an insurance company.*

*If you can't remember the exact details, please provide your best answer possible based on what you can remember.*

*pulm_direct was displayed if the care_person response was* "Child".

*pulm_direct* How much did you pay out-of-pocket for direct medical costs - such as visit fees, copays, deductibles, lab fees, x-ray - related to the first time your child went to the pulmonology clinic for their most recent COVID-19? *Round to the nearest $10 value.*

________________________________________________________________

*pulm_non* How much did you pay for non-medical costs - such as gas, meals, hotel stays, and other similar costs - related to the first time you went with your *{care_person}* to the pulmonology clinic for their most recent COVID-19? *Round to the nearest $10 value.*

________________________________________________________________

*pulm_desc* Please describe any non-medical costs you had. *Type "None" if you did not have any non-medical costs.*

________________________________________________________________

*Page Break*

*Cardiology Intro Block and Cardiology Visits Block were displayed if the spec_type response was* "Cardiologist (heart doctor)".

**Cardiology Intro Block**

*Answer the following questions based on when your {care_person} most recently had COVID-19.*

*card_num* How many times did your *{care_person}* visit a cardiology clinic (including telehealth/virtual visits) for their most recent COVID-19?

________________________________________________________________

*card_virtual* How many of these visits were virtual?

________________________________________________________________

*Page Break*

**Cardiology Visits Intro Block**
*Answer the following questions based on when your {care_person} most recently had COVID-19. If you can't remember the exact details, please provide your best answer possible based on what you can remember.*

*card_date* When was the first time you went with your *{care_person}* to a cardiology clinic for their most recent COVID-19?

| Month | Year |
|---|---|
| ▼ January (1 ... December (12) | ▼ 2020 (1 ... 2023 (4) |

[*A pulldown menu was displayed with the months in chronological order, and a pulldown menu was displayed with the years in chronological order*.]

*card_travel* How long did it take you to travel to the cardiology clinic?

_____ hours _____ minutes

*card_wait* How much time did you spend at the cardiology clinic (including wait and visit times)?

_____ hours _____ minutes

*Page Break*

*Display This Question:*
*If card_num > card_virtual*

*card_last* When was the last time you went with your *{care_person}* to a cardiology clinic for their most recent COVID-19?

| Month | Year |
|---|---|
| ▼ January (1 ... December (12) | ▼ 2020 (1 ... 2023 (4) |

[*A pulldown menu was displayed with the months in chronological order, and a pulldown menu was displayed with the years in chronological order*.]

*Page Break*

*Now we are going to ask you about out-of-pocket costs you might have had related to your {care_person}'s first cardiology clinic visit when they most recently had COVID-19.*

***Out-of-pocket costs** are payments you made that were not reimbursed (paid back) by an insurance company.*

*If you can't remember the exact details, please provide your best answer possible based on what you can remember.*

*card_direct was displayed if the care_person response was* "Child".

*card_direct* How much did you pay out-of-pocket for direct medical costs - such as visit fees, copays, deductibles, lab fees, x-ray - related to the first time your child went to the cardiology clinic for their most recent COVID-19? *Round to the nearest $10 value.*

________________________________________________________________

*card_non* How much did you pay for non-medical costs - such as gas, meals, hotel stays, and other similar costs - related to the first time you went with your *{care_person}* to the cardiology clinic for their most recent COVID-19? *Round to the nearest $10 value.*

________________________________________________________________

*card_desc* Please describe any non-medical costs you had. *Type "None" if you did not have any non-medical costs.*

________________________________________________________________

*Page Break*

*Oncology Intro Block and Oncology Visits Block were displayed if the spec_type response was* "Oncologist (cancer doctor)".

**Oncology Intro Block**

*Answer the following questions based on when your {care_person} most recently had COVID-19.*

*onc_num* How many times did your *{care_person}* visit an oncology clinic (including telehealth/virtual visits) for their most recent COVID-19?

________________________________________________________________

*onc_virtual* How many of these visits were virtual?

________________________________________________________________

*Page Break*

**Oncology Visits Block**
*Answer the following questions based on when your {care_person} most recently had COVID-19. If you can't remember the exact details, please provide your best answer possible based on what you can remember.*

*onc_date* When was the first time you went with your *{care_person}* to an oncology clinic for their most recent COVID-19?

| Month | Year |
| --- | --- |
| ▼ January (1 ... December (12) | ▼ 2020 (1 ... 2023 (4) |

[*A pulldown menu was displayed with the months in chronological order, and a pulldown menu was displayed with the years in chronological order*.]

*onc_travel* How long did it take you to travel to the oncology clinic?

_____ hours _____ minutes

*onc_wait* How much time did you spend at the oncology clinic (including wait and visit times)?

_____ hours _____ minutes

*Page Break*

*Display This Question:*
*If onc_num = onc_virtual*

*onc_last* When was the last time you went with your *{care_person}* to an oncology clinic for their most recent COVID-19?

| Month | Year |
| --- | --- |
| ▼ January (1 ... December (12) | ▼ 2020 (1 ... 2023 (4) |

[*A pulldown menu was displayed with the months in chronological order, and a pulldown menu was displayed with the years in chronological order*.]

*Page Break*

*Now we are going to ask you about out-of-pocket costs you might have had related to your {care_person}'s first oncology clinic visit when they most recently had COVID-19.*

***Out-of-pocket costs** are payments you made that were not reimbursed (paid back) by an insurance company.*

*If you can't remember the exact details, please provide your best answer possible based on what you can remember.*

*onc_direct was displayed if the care_person response was* "Child".

*onc_direct* How much did you pay out-of-pocket for direct medical costs - such as visit fees, copays, deductibles, lab fees, x-ray - related to the first time your child went to the oncology clinic for their most recent COVID-19? *Round to the nearest $10 value.*

________________________________________________________________

*onc_non* How much did you pay for non-medical costs - such as gas, meals, hotel stays, and other similar costs - related to the first time you went with your *{care_person}* to the oncology clinic for their most recent COVID-19? *Round to the nearest $10 value.*

________________________________________________________________

*onc_desc* Please describe any non-medical costs you had. *Type "None" if you did not have any non-medical costs.*

________________________________________________________________

*Page Break*

*OB/GYN Intro Block and OB/GYN Visits Block were displayed if the spec_type response was* "Obstetrician/Gynecologist (OB/GYN)".

**OB/GYN Intro Block**

*Answer the following questions based on when your {care_person} most recently had COVID-19.*

*ob_num* How many times did your *{care_person}* visit an obstetrics/gynecology clinic (including telehealth/virtual visits) for their most recent COVID-19?

________________________________________________________________

*ob_virtual* How many of these visits were virtual?

________________________________________________________________

*Page Break*

**OB/GYN Visits Block**
*Answer the following questions based on when your* *{care_person}* *most recently had COVID-19. If you can't remember the exact details, please provide your best answer possible based on what you can remember.*

*ob_date* When was the first time you went with your *{care_person}* to an obstetrics/gynecology clinic for their most recent COVID-19?

| Month | Year |
| --- | --- |
| ▼ January (1 ... December (12) | ▼ 2020 (1 ... 2023 (4) |

[*A pulldown menu was displayed with the months in chronological order, and a pulldown menu was displayed with the years in chronological order*.]

*ob_travel* How long did it take you to travel to the obstetrics/gynecology clinic?

_____ hours _____ minutes

*ob_wait* How much time did you spend at the obstetrics/gynecology clinic (including wait and visit times)?

_____ hours _____ minutes

*Page Break*

*Display This Question:*
*If ob_num > ob_num*

*ob_last* When was the last time you went with your *{care_person}* to an obstetrics/gynecology clinic for their most recent COVID-19?

| Month | Year |
| --- | --- |
| ▼ January (1 ... December (12) | ▼ 2020 (1 ... 2023 (4) |

[*A pulldown menu was displayed with the months in chronological order, and a pulldown menu was displayed with the years in chronological order*.]

*Page Break*

*Now we are going to ask you about out-of-pocket costs you might have had related to your {care_person}'s first obstetrics/gynecology clinic visit when they most recently had COVID-19.*

***Out-of-pocket costs** are payments you made that were not reimbursed (paid back) by an insurance company.*

*If you can't remember the exact details, please provide your best answer possible based on what you can remember.*

*ob_direct was displayed if the care_person response was* "Child".
*ob_direct* How much did you pay out-of-pocket for direct medical costs - such as visit fees, copays, deductibles, lab fees, x-ray - related to the first time your child went to the obstetrics/gynecology clinic for their most recent COVID-19? *Round to the nearest $10 value.*

________________________________________________________________

*ob_non* How much did you pay for non-medical costs - such as gas, meals, hotel stays, and other similar costs - related to the first time you went with your *{care_person}* to the obstetrics/gynecology clinic for their most recent COVID-19? *Round to the nearest $10 value.*

________________________________________________________________

*ob_desc* Please describe any non-medical costs you had. *Type "None" if you did not have any non-medical costs.*

________________________________________________________________

*Page Break*

*Physical Therapy Intro Block and Physical Therapy Visits Block were displayed if the spec_type response was* "Physical therapist".
**Physical Therapy Intro Block**
*Answer the following questions based on when your {care_person} most recently had COVID-19.*

*pt_num* How many times did your *{care_person}* visit a physical therapy clinic (including telehealth/virtual visits) for their most recent COVID-19?

________________________________________________________________

*pt_virtual* How many of these visits were virtual?

________________________________________________________________

*Page Break*

**Physical Therapy Visits Block**
*Answer the following questions based on when your {care_person} most recently had COVID-19. If you can't remember the exact details, please provide your best answer possible based on what you can remember.*

*pt_date* When was the first time you went with your *{care_person}* to a physical therapy clinic for their most recent COVID-19?

| Month | Year |
|---|---|
| ▼ January (1 ... December (12) | ▼ 2020 (1 ... 2023 (4) |

[*A pulldown menu was displayed with the months in chronological order, and a pulldown menu was displayed with the years in chronological order*.]

*pt_travel* How long did it take you to travel to the physical therapy clinic?

_____ hours _____ minutes

*pt_wait* How much time did you spend at the physical therapy clinic (including wait and visit times)?

_____ hours _____ minutes

*Page Break*

*Display This Question:*
*If pt_num > pt_virtual*

*pt_last* When was the last time you went with your *{care_person}* to a physical therapy clinic for their most recent COVID-19?

| Month | Year |
|---|---|
| ▼ January (1 ... December (12) | ▼ 2020 (1 ... 2023 (4) |

[*A pulldown menu was displayed with the months in chronological order, and a pulldown menu was displayed with the years in chronological order*.]

*Page Break*

*Now we are going to ask you about out-of-pocket costs you might have had related to your {care_person}'s first physical therapy clinic visit when they most recently had COVID-19.*

***Out-of-pocket costs** are payments you made that were not reimbursed (paid back) by an insurance company.*

*If you can't remember the exact details, please provide your best answer possible based on what you can remember.*

*pt_direct was displayed if the care_person response was* "Child".
*pt_direct* How much did you pay out-of-pocket for direct medical costs - such as visit fees, copays, deductibles, lab fees, x-ray - related to the first time your child went to the physical therapy clinic for their most recent COVID-19? *Round to the nearest $10 value.*

________________________________________________________________

*pt_non* How much did you pay for non-medical costs - such as gas, meals, hotel stays, and other similar costs - related to the first time you went with your *{care_person}* to the physical therapy clinic for their most recent COVID-19? *Round to the nearest $10 value.*

________________________________________________________________

*pt_desc* Please describe any non-medical costs you had. *Type "None" if you did not have any non-medical costs.*

________________________________________________________________

*Page Break*

*Occupational Therapy Intro Block and Occupational Therapy Visits Block were displayed if the spec_type response was* "Occupational therapist".
**Occupational Therapy Intro Block**
*Answer the following questions based on when your {care_person} most recently had COVID-19.*

*ot_num* How many times did your *{care_person}* visit an occupational therapy clinic (including telehealth/virtual visits) for their most recent COVID-19?

________________________________________________________________

*ot_virtual* How many of these visits were virtual?

________________________________________________________________

*Page Break*

**Occupational Therapy Visits Block**
*Answer the following questions based on when your {care_person} most recently had COVID-19. If you can't remember the exact details, please provide your best answer possible based on what you can remember.*

*ot_date* When was the first time you went with your *{care_person}* to an occupational therapy clinic for their most recent COVID-19?

| Month | Year |
|---|---|
| ▼ January (1 ... December (12) | ▼ 2020 (1 ... 2023 (4) |

[*A pulldown menu was displayed with the months in chronological order, and a pulldown menu was displayed with the years in chronological order*.]

*ot_travel* How long did it take you to travel to the occupational therapy clinic?

_____ hours _____ minutes

*ot_wait* How much time did you spend at the occupational therapy clinic (including wait and visit times)?

_____ hours _____ minutes

*Page Break*

*Display This Question:*
*If ot _num> ot_virtual*

*ot_last* When was the last time you went with your *{care_person}* to an occupational therapy clinic for their most recent COVID-19?

| Month | Year |
|---|---|
| ▼ January (1 ... December (12) | ▼ 2020 (1 ... 2023 (4) |

[*A pulldown menu was displayed with the months in chronological order, and a pulldown menu was displayed with the years in chronological order*.]

*Page Break*

*Now we are going to ask you about out-of-pocket costs you might have had related to your {care_person}'s first occupational therapy clinic visit when they most recently had COVID-19.*

***Out-of-pocket costs** are payments you made that were not reimbursed (paid back) by an insurance company.*

*If you can't remember the exact details, please provide your best answer possible based on what you can remember.*

*ot_direct was displayed if the care_person response was* "Child".

*ot_direct* How much did you pay out-of-pocket for direct medical costs - such as visit fees, copays, deductibles, lab fees, x-ray - related to the first time your child went to the occupational therapy clinic for their most recent COVID-19? *Round to the nearest $10 value.*

________________________________________________________________

*ot_non* How much did you pay for non-medical costs - such as gas, meals, hotel stays, and other similar costs - related to the first time you went with your *{care_person}* to the occupational therapy clinic for their most recent COVID-19? *Round to the nearest $10 value.*

________________________________________________________________

*ot_desc* Please describe any non-medical costs you had. *Type "None" if you did not have any non-medical costs.*

________________________________________________________________

*Page Break*

*Other Visits Intro Block and Other Visits Block were displayed if the spec_type response was* "Other - please specify:".

**Other Visits Intro Block**

*Answer the following questions based on when your {care_person} most recently had COVID-19.*

*other_num* How many times did your *{care_person}* visit the *{spec_type Other response}* clinic (including telehealth/virtual visits) for their most recent COVID-19?

________________________________________________________________

*other_virtual* How many of these visits were virtual?

________________________________________________________________

*Page Break*

**Other Visits Block**
*Answer the following questions based on when your {care_person} most recently had COVID-19. If you can't remember the exact details, please provide your best answer possible based on what you can remember.*

*other_date* When was the first time you went with your *{care_person}* to a *{spec_type Other response}* clinic for their most recent COVID-19?

| Month | Year |
|---|---|
| ▼ January (1 ... December (12) | ▼ 2020 (1 ... 2023 (4) |

[*A pulldown menu was displayed with the months in chronological order, and a pulldown menu was displayed with the years in chronological order*.]

*other_travel* How long did it take you to travel to the *{spec_type Other response}* clinic?

_____ hours _____ minutes

*other_wait* How much time did you spend at the *{spec_type Other response}* clinic (including wait and visit times)?

_____ hours _____ minutes

*Page Break*

*Display This Question:*
*If other_num > other_virtual*

*other_last* When was the last time you went with your *{care_person}* to a *{spec_type Other response}* clinic for their most recent COVID-19?

| Month | Year |
|---|---|
| ▼ January (1 ... December (12) | ▼ 2020 (1 ... 2023 (4) |

[*A pulldown menu was displayed with the months in chronological order, and a pulldown menu was displayed with the years in chronological order*.]

*Page Break*

*Now we are going to ask you about out-of-pocket costs you might have had related to your {care_person}'s first {spec_type Other response} clinic visit when they most recently had COVID-19.*

***Out-of-pocket costs** are payments you made that were not reimbursed (paid back) by an insurance company.*

*If you can't remember the exact details, please provide your best answer possible based on what you can remember.*

*other_direct was displayed if the care_person response was* "Child".

*other_direct* How much did you pay out-of-pocket for direct medical costs - such as visit fees, copays, deductibles, lab fees, x-ray - related to the first time your child went to the *{spec_type Other response}* clinic for their most recent COVID-19? *Round to the nearest $10 value.*

________________________________________________________________

*other_non* How much did you pay for non-medical costs - such as gas, meals, hotel stays, and other similar costs - related to the first time you went with your *{care_person}* to the *{spec_type Other response}* clinic for their most recent COVID-19? *Round to the nearest $10 value.*

________________________________________________________________

*other_desc* Please describe any non-medical costs you had. *Type "None" if you did not have any non-medical costs.*

________________________________________________________________

*Page Break*

*In Patient Rehab Block was displayed if the covid_provider response was* "Yes" *to* "In-Patient Rehabilitation Clinic".

**In Patient Rehab Block**

*Now we are going to ask you about inpatient rehabilitation your {care_person} experienced when they most recently had COVID-19, which could include care for long-term symptoms (Long COVID).*

*rehab_num* How many times was your *{care_person}* admitted to an inpatient rehabilitation clinic as a result of their most recent COVID-19?

________________________________________________________________

*Page Break*

*Answer the following questions based on when your {care_person} most recently had COVID-19. If you don't know or can't remember the exact details, please provide your best answer possible based on what you can remember.*

*rehab_date* When was the first time your *{care_person}* was admitted to an inpatient rehabilitation clinic as a result of their most recent COVID-19?

| Month | Year |
|---|---|
| ▼ January (1 ... December (12) | ▼ 2020 (1 ... 2023 (4) |

[*A pulldown menu was displayed with the months in chronological order, and a pulldown menu was displayed with the years in chronological order*.]

*rehab_days* How many days were they at the inpatient rehabilitation clinic?

________________________________________________________________

*Page Break*

*rehab_last was displayed if the hosp_num response was greater than one.*
*rehab_last* When was the last time your *{care_person}* was admitted to an inpatient rehabilitation clinic as a result of their most recent COVID-19?

| Month | Year |
|---|---|
| ▼ January (1 ... December (12) | ▼ 2020 (1 ... 2023 (4) |

[*A pulldown menu was displayed with the months in chronological order, and a pulldown menu was displayed with the years in chronological order*.]

*rehab_ldays was displayed if the hosp_num response was greater than one.*
*rehab_ldays* How many days were they at the inpatient rehabilitation clinic?

________________________________________________________________

*Page Break*

*Now we are going to ask you about out-of-pocket costs you might have had related to the first time your {care_person} was admitted to an inpatient rehabilitation clinic as a result of their most recent COVID-19.*

***Out-of-pocket costs** are payments you made that were not reimbursed (paid back) by an insurance company.*

*If you don't know or can't remember the exact details, please provide your best answer possible based on what you can remember.*

*rehab_direct was displayed if the care_person response was* "Child".

*rehab_direct* How much did you pay out-of-pocket for direct medical costs - such as visit fees, copays, deductibles, lab fees, x-ray - related to the first time your child was admitted to an inpatient rehabilitation clinic as a result of their most recent COVID-19? *Round to the nearest $10 value.*

________________________________________________________________

*rehab_non* How much did you pay for non-medical costs - such as gas, meals, hotel stays, and other similar costs - related to the first time your *{care_person}* was admitted to an inpatient rehabilitation clinic as a result of their most recent COVID-19? *Round to the nearest $10 value.*

________________________________________________________________

*rehab_desc* Please describe any non-medical costs you had. *Type "None" if you did not have any non-medical costs.*

________________________________________________________________

*Page Break*

*Home Health Care Services Block was displayed if the covid_home response was* "Yes".

**Home Health Care Services Block**

*Now we are going to ask you about home health care services your {care_person} received related to their most recent had COVID-19, which could include care for long-term symptoms (Long COVID). If you don't know or can't remember the exact details, please provide your best answer possible on what you can remember.*

*home_date* When did your *{care_person}* first start receiving home health care services (example: in-home rehabilitation) as a result of their most recent COVID-19?

| | Month | Year |
|---|---|---|
| (1) | ▼ January (1 ... December (12) | ▼ 2020 (1 ... 2023 (4) |

*home_last* When was the last time your *{care_person}* received home health care services as a result of their most recent COVID-19?

| | Month | Year |
|---|---|---|
| (1) | ▼ January (1 ... December (12) | ▼ 2020 (1 ... 2023 (4) |

*home_days* How many days per week {*LongCovid*} your *{care_person}* receive home health care services on average as a result of their most recent COVID-19?

________________________________________________________________

*home_hours* How many hours per day {*LongCovid*} your *{care_person}* receive home health care services on average as a result of their most recent COVID-19?

________________________________________________________________

*Page Break*

*Now we are going to ask you about out-of-pocket costs you might have had related to home health care services your child received as a result of their most recent COVID-19.*

***Out-of-pocket costs** are payments you made that were not reimbursed (paid back) by an insurance company.*

*If you don't know or can't remember the exact details, please provide your best answer possible based on what you can remember.*

*home_direct was displayed if the care_person response was* "Child".

*home_direct* How much did you pay out-of-pocket for direct medical costs - such as visit fees, copays, deductibles - related to home health care services your child received as a result of their most recent COVID-19? *Round to the nearest $10 value.*

________________________________________________________________

*Page Break*

*Medications Block was displayed if the covid_meds response was* “Yes”.

**Medications Block**

*Now we are going to ask you about medicines your {care_person} received related to their most recent had COVID-19, which could include medicines for long-term symptoms (Long COVID).*

*med_prescription* How many types of prescription medicines did your *{care_person}* take for their most recent COVID-19?

________________________________________________________________

*prescription_price* How much did your *{care_person}* pay out-of-pocket for these prescription medicines? *Round to the nearest $10 value.*

________________________________________________________________

*med_otc* How many types of over-the-counter (non-prescription) medicines did your *{care_person}* take for their most recent COVID-19?

________________________________________________________________

*otc_price* How much did your *{care_person}* pay out-of-pocket for these over-the-counter medicines? *Round to the nearest $10 value.*

________________________________________________________________

*Page Break*

**Insurance and Reimbursement Block**

*Now we are going to ask you about any health insurance you may have had during your {care_person}'s most recent COVID-19, which could include care for long-term symptoms (Long COVID).*

*ins_coverage* Did you have any health insurance or health coverage plans during your *{care_person}*'s most recent COVID-19?

- ○ No (2)
- ○ Yes (1)
- ○ Unsure/I can't remember (3)

*Participants skipped to the Confidence Estimates Block if the ins_coverage response was NOT* “Yes”.

*Page Break*

_ins_type_ Were you covered by any of the following types of health insurance or health coverage plans during your _{care_person}_'s most recent COVID-19? *Mark "No" or "Yes" for EACH type of insurance.*

| | No (1) | Yes (2) |
|---|---|---|
| Private insurance through a current or former employer or union (of you or another family member) (1) | ○ | ○ |
| Insurance purchased directly from an insurance company (2) | ○ | ○ |
| Insurance purchased directly from HealthCare.gov or a state Health Insurance Marketplace (3) | ○ | ○ |
| Medicare (for people 65 and older, or people with certain disabilities) (4) | ○ | ○ |
| Medicaid, Medical Assistance, or any kind of government-assistance plan for people with low incomes or a disability (5) | ○ | ○ |
| TRICARE or other military health care (6) | ○ | ○ |
| VA (7) | ○ | ○ |
| Indian Health Service (8) | ○ | ○ |
| Any other type of health insurance or health coverage plan - Specify: (9) | ○ | ○ |

_ins_reimburse_ Were you partially or fully reimbursed (paid back) by a health insurance company or public assistance program for any payments you made for care, services, or medicines your _{care_person}_ received related to their most recent COVID-19?

- ○ No (1)
- ○ Yes (2)
- ○ Waiting to find out (3)
- ○ Don't know/Unsure (4)

*Participants skipped to the Confidence Estimates Block if the ins_reimburse response was NOT* "Yes".

*Page Break*

_ins_amount_ How much money were you reimbursed (paid back) by a health insurance company or public assistance program for any payments you made for care, services, or medicines your _{care_person}_ received related to their most recent COVID-19? *Round to the nearest $10 value.*

________________________________________________________________

*ins_time* How much time did you spend talking to your insurance company about reimbursements or other issues related to your *{care_person}*'s most recent COVID-19?

_____ hours _____ minutes

*Page Break*

**Confidence Estimates Block**
*Now we are going to ask you about how confident you were with some of the answers you provided in the previous sections.*

*conf_medical* Overall, how confident are you that the numbers you provided about medical costs are accurate?

| 1 | 2 | 3 | 4 | 5 |
|---|---|---|---|---|
| Not at all confident | | | | Very confident |

*conf_non* Overall, how confident are you that the numbers you provided about non-medical costs are accurate?

| 1 | 2 | 3 | 4 | 5 |
|---|---|---|---|---|
| Not at all confident | | | | Very confident |

*conf_travel* Overall, how confident are you that the numbers you provided about travel time are accurate?

| 1 | 2 | 3 | 4 | 5 |
|---|---|---|---|---|
| Not at all confident | | | | Very confident |

*conf_wait* Overall, how confident are you that the numbers you provided about time at the clinic (wait and visit times) are accurate?

| 1 | 2 | 3 | 4 | 5 |
|---|---|---|---|---|
| Not at all confident | | | | Very confident |

*Page Break*

**Financial Challenges Block**
*Now we are going to ask you about any challenges you may have had when you're your {care_person} most recently had COVID-19, which could include long-term symptoms (Long COVID).*

*Answer the following questions based on when your {care_person} most recently had COVID-19.*

*bills_problem* Did you or anyone else in your household have problems paying, or were not able to pay, any COVID-related medical bills? *Select all that apply.*

- ☐ ⊗No (1)
- ☐ Yes, when I had COVID (2)
- ☐ Yes, after I had COVID (3)
- ☐ Yes, currently (4)

*bills_wait* Did you or anyone else in your household ever wait to get any medical care (COVID-related or otherwise) because of worry about the cost?

- ☐ ⊗No (1)
- ☐ Yes, when I had COVID (2)
- ☐ Yes, after I had COVID (3)
- ☐ Yes, currently (4)

*bills_meds* Did you or anyone else in your household ever take less or skip doses of medicine (COVID-related or otherwise) because of worry about the cost?

- ☐ ⊗No (1)
- ☐ Yes, when I had COVID (2)
- ☐ Yes, after I had COVID (3)
- ☐ Yes, currently (4)

*bills_care* Was there any time you or anyone else in your household needed medical care (COVID-related or otherwise) but did not get it because of worry about the cost?

- ☐ ⊗No (1)
- ☐ Yes, when I had COVID (2)
- ☐ Yes, after I had COVID (3)
- ☐ Yes, currently (4)

*bills_prescription* Was there any time you or anyone else in your household needed a prescription medicine (COVID-related or otherwise) but did not get it because of worry about the cost?

- ☐ ⊗No (1)
- ☐ Yes, when I had COVID (2)
- ☐ Yes, after I had COVID (3)
- ☐ Yes, currently (4)

*Page Break*

*bills_other* Did you or anyone else in your household have problems paying, or were not able to pay, any non-medical bills (examples: mortgage/rent; student loans; car loan; credit cards)? *Select all that apply.*

- ☐ ⊗No (1)
- ☐ Yes, when I had COVID (2)
- ☐ Yes, after I had COVID (3)
- ☐ Yes, currently (4)

*bills_meals* Did you or anyone else in your household ever not eat as much, or skip meals, because there wasn't enough money for food? *Select all that apply.*

- ☐ ⊗No (1)
- ☐ Yes, when I had COVID (2)
- ☐ Yes, after I had COVID (3)
- ☐ Yes, currently (4)

*covid_absent* How many days of school or work did you miss as a result of your *{care_person}*'s most recent COVID-19?

________________________________________________________________

*Page Break*

*sacrifice* Please use the space below to type in anything other “sacrifices” (cost you money or time) you may have made while you were acting as the primary caregiver for during your *{care_person}*’s most recent COVID-19.

________________________________________________________________

*Page Break*

**Vaccination History and Attitudes Block**

*Almost done! Now we are going to ask you about vaccines you and your {care_person} may have received, and for some additional information about you.*

*relative_flu was displayed if the care_person response was* "Child" *OR* "Spouse/Partner".

*relative_flu* Over the last 3 years, how many times has your *{care_person}* gotten an annual flu vaccine?

- Never (1)
- Sometimes (2)
- Every year (3)
- Don't know/Unsure (4)

*relative_covid was displayed if the care_person response was* "Child" *OR* "Spouse/Partner".

*relative_covid* Has your *{care_person}* ever received a COVID-19 vaccine?

No (2) Yes (1) Don't know/Unsure (3)

*relative_booster was displayed if the care_person response was* "Child" *OR* "Spouse/Partner".

*relative_booster* Has your *{care_person}* received a COVID-19 booster shot since September 1, 2022?

No (2) Yes (1) Don't know/Unsure (3)

*self_flu* Over the last 3 years, how many times did you get an annual flu vaccine?

Never (1) Sometimes (2) Every year (3)

*self_covid* Have you ever received a COVID-19 vaccine?

No (2) Yes (1)

*self_booster* Have you received a COVID-19 booster shot since September 1, 2022?

No (2) Yes (1)

*vax_attitudes* How do you feel about {*vaxgroup*}, in general?

| 1 | 2 | 3 | 4 | 5 |
|---|---|---|---|---|
| Very negative | | | | Very positive |

*Page Break*

**Additional Demographics Block**
*state* Which state do you live in?

[*U.S. states and territories were listed in alphabetical order, with* District of Columbia *coded as* “9”, and “40” was skipped (i.e., Rhode Island *was coded as* “41” *instead of* “40”.]

▼ Alabama (1) ... Wyoming (51)

*rurality* How would you describe the area in which you live?

Urban (1) Suburban (2) Rural (3)

*educ* What is the highest level of schooling you have completed?

- Less than high school (1)
- High school (Diploma or GED) (2)
- Bachelor's degree (BS, BA, etc.) (3)
- Master's degree (MA, MPH, etc.) (4)
- Doctoral/Professional degree (PhD, MD, etc.) (5)

*income* Which of the following categories best describes your total household income, from all sources, before taxes?

- Less than $20,000 (1)
- $20,000 - $39,999 (2)
- $40,000 - $59,999 (3)
- $60,000 - $79,999 (4)
- $80,000 - $99,999 (5)
- $100,000 - $149,999 (6)
- $150,000 - $199,999 (7)
- $200,000 - $249,999 (8)
- $250,000 - $299,999 (9)
- $300,000 or more (10)
- Prefer not to answer (11)

*income_people* Including yourself, how many people does that income support?

________________________________________________________________

*Page Break*

*health_conditions* Have you been diagnosed with any of the following conditions? *Select all that apply.*

- ▢ Chronic obstructive pulmonary disease (COPD) (1)
- ▢ Heart disease (2)
- ▢ Diabetes (3)
- ▢ Immunocompromised condition or taking medications that suppress the immune system (4)
- ▢ Kidney disease or need for dialysis (5)
- ▢ Liver disease (6)
- ▢ ⊗None of these (7)

*Page Break*

*comments* Do you have any other comments or thoughts you would like to share with us regarding challenges you've experienced as a result of caring for someone with COVID-19?

________________________________________________________________
