## Appendix 1 for "Medical and non-medical out-of-pocket cost and time burden of COVID-19 illness in U.S. children and adults: a cross-sectional survey study"

Table 1. Respondent demographics, n (%), caregivers of patients ≥18 y

| | Caregiver of patients ≥18 y (n=185) |
|---|---|
| **Female** | 90 (48.7) |
| **Age of respondent, mean (SD)** | 48.4 (13.9) |
| **Age of patient, mean (SD)** | 57.1 (18.8) |
| **Hispanic** | 47 (25.4) |
| **Race*** | |
| White | 140 (75.7) |
| Black | 35 (18.9) |
| Other† | 22 (11.9) |
| **Education level** | |
| Less than high school | 2 (1.1) |
| High school or GED | 60 (32.4) |
| Bachelor's degree | 91 (49.2) |
| Master's degree | 22 (11.9) |
| Doctoral/professional degree | 10 (5.4) |
| **Geographic area** | |
| Urban | 57 (30.8) |
| Suburban | 100 (54.1) |
| Rural | 28 (15.1) |
| **Household income** | |
| <$40,000 | 37 (20.0) |
| $40,000 - <$80,000 | 64 (34.6) |
| $80,000 - <$150,000 | 51 (27.6) |
| ≥$150,000 | 30 (16.2) |
| Missing | 3 (1.6) |
| **Insurance*** | |
| Any insurance | 80 (95.2) |
| Private insurance | |
| Employer | 42 (53.2) |
| From insurance company | 8 (10.1) |
| From insurance marketplace | 8 (10.3) |
| Medicare | 22 (27.5) |
| Medicaid | 15 (19.7) |
| Tricare | 7 (9.0) |
| Veterans Affairs | 2 (2.7) |
| Indian Health Service | 1 (1.3) |
| Other | 4 (6.5) |
| Missing | 1 (1.2) |
| **COVID-19 severity‡ (mean (SD))** | 4.0 (1.0) |
| **Long COVID** | 83 (44.9) |
| **Long COVID severity‡ (mean (SD))** | 4.2 (1.1) |
| **COVID-19 vaccine, ever** | 64 (76.2) |
| **COVID-19 booster** | 41 (48.8) |

* Respondents could select >1 answer

† Includes: Asian, American Indian, Alaskan Native, Native Hawaiian, Other Pacific Islander, none of these, prefer not to answer

‡ 6-point scale where 1 = no symptoms and 6 = very severe symptoms. Hospitalization and ICU care were defined as "very severe"

Table 2. Respondents who reported receiving care for their COVID illness, by setting, n(%), caregivers of patients ≥18 y

| | Caregiver of patients ≥18 y (n=185) |
|---|---|
| Primary care | 108 (58.4) |
| Urgent care | 90 (49.2) |
| Specialist visits | 25 (13.8) |
| Emergency department visits | 56 (30.6) |
| Hospitalization | 41 (22.2) |
| Home health | 38 (20.5) |
| Inpatient rehab | 17 (9.3) |

Table 3. Visit number, time spent, and out of pocket costs by care setting, caregivers of patients ≥18 y

| | Caregiver of patients ≥18 y (n=185) | | | | | |
|---|---|---|---|---|---|---|
| | **N** | **Mean** | **SD** | **Median** | **Min** | **Max** |
| **Primary care** | | | | | | |
| Number of visits | 185 | 1.3 | 1.5 | 1 | 0 | 8 |
| Time spent (min) | 108 | 117.8 | 78.9 | 100 | 0 | 336 |
| OOP medical costs ($) | 2 | 57.5 | 60.1 | 57.5 | 15 | 100 |
| Non-medical costs ($) | 95 | 48.1 | 87.8 | 20 | 0 | 500 |
| **Urgent care** | | | | | | |
| Number of visits | 183 | 0.6 | 0.8 | 0 | 0 | 4 |
| Time spent (min) | 92 | 134.7 | 81.4 | 120 | 0 | 355 |
| OOP medical costs ($) | 8 | 16.9 | 27.3 | 0 | 0 | 75 |
| Non-medical costs ($) | 90 | 37.3 | 50.5 | 17.5 | 0 | 200 |
| **Emergency department** | | | | | | |
| Number of visits | 183 | 0.3 | 0.6 | 0 | 0 | 3 |
| Time spent (min) | 58 | 395.7 | 560.2 | 255.5 | 0 | 3270 |
| OOP medical costs ($) | 1 | 0.0 | . | 0 | 0 | 0 |
| Non-medical costs ($) | 54 | 86.7 | 209.2 | 32.5 | 0 | 1500 |
| **Hospitalization** | | | | | | |
| Number of visits | 185 | 0.3 | 0.6 | 0 | 0 | 4 |
| Length of stay (d) | 41 | 6.7 | 7.5 | 5 | 1 | 45 |
| OOP medical costs ($) | 1 | 80.0 | . | 80 | 80 | 80 |
| Non-medical costs ($) | 40 | 100.6 | 114.9 | 50 | 0 | 400 |

OOP = out of pocket; SD = standard deviation

Table 4. Financial challenges resulting from most recent COVID-19 illness, caregivers of patients ≥18 y

| | Caregiver of patients ≥18 y (n=185) |
|---|---|
| **Problems paying COVID-19-related medical bills** | |
| No | 156 (84.8) |
| Yes, when I had COVID-19 | 17 (9.2) |
| Yes, after I had COVID-19 | 11 (6.0) |
| Yes, currently | 8 (4.4) |
| **Wait to get medical care due to cost** | |
| No | 156 (85.3) |
| Yes, when I had COVID-19 | 17 (9.3) |
| Yes, after I had COVID-19 | 5 (2.7) |
| Yes, currently | 10 (5.5) |
| **Take less or skip doses of medicine due to cost** | |
| No | 165 (89.7) |
| Yes, when I had COVID-19 | 13 (7.1) |
| Yes, after I had COVID-19 | 3 (1.6) |
| Yes, currently | 5 (2.7) |
| **Did not get medical care due to cost** | |
| No | 160 (87.0) |
| Yes, when I had COVID-19 | 12 (6.5) |
| Yes, after I had COVID-19 | 5 (2.7) |
| Yes, currently | 11 (6.0) |
| **Did not get prescription medicine due to cost** | |
| No | 160 (87.0) |
| Yes, when I had COVID-19 | 9 (4.9) |
| Yes, after I had COVID-19 | 10 (5.4) |
| Yes, currently | 11 (6.0 |
| **Problems paying non-medical bills** | |
| No | 148 (80.0) |
| Yes, when I had COVID-19 | 20 (10.8) |
| Yes, after I had COVID-19 | 14 (7.6) |
| Yes, currently | 14 (7.6) |
| **Skip or limit meals due to cost** | |
| No | 159 (86.4) |
| Yes, when I had COVID-19 | 11 (6.0) |
| Yes, after I had COVID-19 | 11 (6.0) |
| Yes, currently | 8 (4.4) |
| **Any financial problem** | |
| Yes, when I had COVID-19 | 42 (22.7) |
| Yes, after I had COVID-19 | 24 (13.0) |
| Yes, currently | 27 (14.6) |
| Yes, at any time | 55 (29.7) |
| None | 130 (70.3) |

Table 5. Exclusion criteria and number of respondents excluded

a. Adult sample

| Criterion | Primary analysis N | Secondary analysis N |
|---|---|---|
| Respondents reporting >10 visits at a care setting | 18 | 18 |
| Respondents with survey duration <1/2 median duration | NA | 55 |
| Respondents reporting low severity COVID-19 illness and a hospitalization | 5 | 5 |
| Respondents reporting on a COVID-19 illness >18 months prior to survey completion | 377 | NA |
| Respondents reporting on a COVID-19 illness >12 months prior to survey completion | NA | 544 |
| Remove respondents reporting high values of time spent receiving care (>97.5 %tile) | 32 | 32 |
| Total excluded | **391** | **568** |
| Total included | **409** | **232** |

b. Child sample, patient age 0-17y

| Criterion | Primary analysis N | Secondary analysis N |
|---|---|---|
| Respondents reporting >10 visits at a care setting | 2 | 2 |
| Respondents with survey duration <1/2 median duration | NA | 9 |
| Respondents reporting low severity COVID-19 illness and a hospitalization | 0 | 0 |
| Respondents reporting on a COVID-19 illness >18 months prior to survey completion | 114 | NA |
| Respondents reporting on a COVID-19 illness >12 months prior to survey completion | NA | 130 |
| Remove respondents reporting high values of time spent receiving care (>97.5 %tile) | 14 | 14 |
| Total excluded | **117** | **133** |
| Total included | **52** | **36** |

Table 6. Percentage of respondents reporting a medical visit and 0 costs (n (%)) and costs for those reporting a medical visit and >0 costs (mean (median))

| | n(%) reporting a medical visit and 0 costs | | | | | | Mean (median) of those reporting a medical visit and >0 costs | | | | | |
|---|---|---|---|---|---|---|---|---|---|---|---|---|
| | Adult sample | | | Child sample | | | Adult sample | | | Child sample | | |
| | No exclusions (n=800) | Primary analysis (n=409) | Secondary analysis (n=232) | No exclusions (n=169) | Primary analysis (n=52) | Secondary analysis (n=36) | No exclusions (n=800) | Primary analysis (n=409) | Secondary analysis (n=232) | No exclusions (n=169) | Primary analysis (n=52) | Secondary analysis (n=36) |
| **Primary care** | | | | | | | | | | | | |
| OOP medical costs ($) | 76 (34.2) | 35 (40.7) | 22 (38.6) | 23 (39.0) | 12 (36.4) | 11 (45.8) | 119.7 (25) | 84.6 (35) | 76.5 (35) | 66.8 (30) | 63.1 (30) | 74.7 (30) |
| Non-medical costs ($) | 75 (33.2) | 37 (42.0) | 26 (44.1) | 18 (27.3) | 11 (31.4) | 6 (25.0) | 101.1 (20) | 36.2 (20) | 34.0 (15) | 38.8 (17.5) | 24.0 (10) | 21.9 (10) |
| **Urgent care** | | | | | | | | | | | | |
| OOP medical costs ($) | 73 (29.0) | 38 (36.5) | 21 (37.5) | 14 (35.9) | 6 (37.5) | 6 (40.0) | 126.2 (30) | 128.1 (40) | 159.9 (30) | 62.0 (40) | 87.5 (50) | 75.0 (50) |
| Non-medical costs ($) | 82 (32.4) | 44 (42.3) | 30 (52.6) | 22 (40.7) | 9 (47.4) | 8 (50.0) | 57.1 (20) | 41.0 (20) | 39.6 (20) | 28.1 (15) | 43.5 (20) | 34.4 (20) |
| **Emergency department** | | | | | | | | | | | | |
| OOP medical costs ($) | 55 (32.5) | 28 (46.7) | 14 (45.2) | 15 (55.6) | 7 (58.3) | 3 (42.9) | 192.9 (25) | 193.5 (80) | 130.6 (20) | 177.5 (85) | 194.0 (100) | 217.5 (150) |
| Non-medical costs ($) | 58 (33.9) | 25 (41.0) | 12 (36.4) | 11 (26.8) | 5 (38.5) | 2 (28.6) | 72.1 (15) | 39.4 (20) | 42.2 (20) | 68.5 (30) | 55.9 (20) | 32.4 (15) |
| **Hospitalization** | | | | | | | | | | | | |
| OOP medical costs ($) | 25 (17.6) | 11 (27.5) | 6 (26.1) | 4 (21.1) | 3 (37.5) | 3 (50.0) | 248.9 (20) | 325.9 (90) | 247.4 (80) | 206.3 (40) | 211.8 (109) | 303.0 (300) |
| Non-medical costs ($) | 24 (16.8) | 12 (29.3) | 6 (25.0) | 3 (9.7) | 3 (37.5) | 2 (33.3) | 116.0 (20) | 163.1 (30) | 164.0 (53) | 51.1 (20) | 67.0 (50) | 80.0 (65) |

Table 7. Healthcare utilization costs, by care setting and demographics, primary analysis

a. By insurance coverage

| | Adult Sample | | | | | | | | Child Sample | | | | | | | |
|---|---|---|---|---|---|---|---|---|---|---|---|---|---|---|---|---|
| | Insured | | | | Uninsured | | | | Insured | | | | Uninsured | | | |
| | N | Mean | SD | Median | N | Mean | SD | Median | N | Mean | SD | Median | N | Mean | SD | Median |
| **Primary care** | | | | | | | | | | | | | | | | |
| OOP medical costs ($) | 78 | 41.4 | 97.3 | 7.5 | 7 | 154.9 | 224.3 | 50.0 | 32 | 40.7 | 66.8 | 20.0 | 1 | 25.0 | . | 25.0 |
| Non-medical costs ($) | 80 | 20.6 | 45.0 | 6.0 | 7 | 28.6 | 27.4 | 20.0 | 31 | 15.0 | 21.0 | 10.0 | 1 | 30.0 | . | 30.0 |
| **Urgent care** | | | | | | | | | | | | | | | | |
| OOP medical costs ($) | 91 | 85.5 | 325.1 | 10.0 | 12 | 56.2 | 141.6 | 8.0 | 15 | 56.7 | 78.1 | 20.0 | 1 | 25.0 | . | 25.0 |
| Non-medical costs ($) | 91 | 24.9 | 66.1 | 5.0 | 12 | 16.0 | 18.9 | 5.0 | 15 | 13.7 | 25.7 | 5.0 | 1 | 0.0 | . | 0.0 |
| **Emergency department** | | | | | | | | | | | | | | | | |
| OOP medical costs ($) | 50 | 78.0 | 175.2 | 0.0 | 10 | 229.2 | 275.0 | 107.5 | 11 | 88.2 | 151.4 | 0.0 | 1 | 0.0 | . | 0.0 |
| Non-medical costs ($) | 51 | 23.9 | 49.8 | 2.0 | 10 | 20.1 | 14.2 | 20.0 | 11 | 18.4 | 23.8 | 12.0 | 1 | 0.0 | . | 0.0 |
| **Hospitalization** | | | | | | | | | | | | | | | | |
| OOP medical costs ($) | 32 | 166.8 | 325.2 | 6.0 | 8 | 514.4 | 665.3 | 400.0 | 7 | 151.3 | 184.5 | 100.0 | 1 | 0.0 | . | 0.0 |
| Non-medical costs ($) | 33 | 97.7 | 348.6 | 5.0 | 8 | 188.4 | 449.8 | 37.5 | 7 | 47.9 | 53.5 | 40.0 | 1 | 0.0 | . | 0.0 |

b. By insurance type

| | Adult Sample | | | | | | | | Child Sample | | | | | | | |
|---|---|---|---|---|---|---|---|---|---|---|---|---|---|---|---|---|
| | Medicaid | | | | Other | | | | Medicaid | | | | Other | | | |
| | N | Mean | SD | Median | N | Mean | SD | Median | N | Mean | SD | Median | N | Mean | SD | Median |
| **Primary care** | | | | | | | | | | | | | | | | |
| OOP medical costs ($) | 19 | 22.6 | 44.2 | 0.0 | 58 | 48.3 | 109.4 | 12.5 | 10 | 25.5 | 41.3 | 0.0 | 21 | 48.9 | 77.1 | 25.0 |
| Non-medical costs ($) | 19 | 14.3 | 19.4 | 7.0 | 60 | 22.9 | 50.7 | 6.0 | 10 | 17.0 | 14.8 | 10.0 | 20 | 14.8 | 24.1 | 10.0 |
| **Urgent care** | | | | | | | | | | | | | | | | |
| OOP medical costs ($) | 31 | 33.1 | 83.0 | 0.0 | 59 | 63.6 | 109.7 | 25.0 | 4 | 50.0 | 100.0 | 0.0 | 11 | 59.1 | 74.2 | 40.0 |
| Non-medical costs ($) | 31 | 21.2 | 48.8 | 5.0 | 59 | 27.3 | 74.3 | 4.0 | 4 | 5.0 | 5.8 | 5.0 | 11 | 16.8 | 29.6 | 5.0 |
| **Emergency department** | | | | | | | | | | | | | | | | |
| OOP medical costs ($) | 26 | 24.5 | 82.9 | 0.0 | 24 | 135.9 | 226.3 | 17.5 | 4 | 0.0 | 0.0 | 0.0 | 7 | 138.6 | 173.4 | 100.0 |
| Non-medical costs ($) | 27 | 17.0 | 42.8 | 0.0 | 24 | 31.6 | 56.6 | 7.5 | 4 | 12.5 | 9.6 | 15.0 | 7 | 21.7 | 29.4 | 12.0 |
| **Hospitalization** | | | | | | | | | | | | | | | | |
| OOP medical costs ($) | 11 | 29.8 | 48.6 | 3.0 | 21 | 238.5 | 383.4 | 11.0 | 2 | 50.0 | 70.7 | 50.0 | 5 | 191.8 | 206.4 | 109.0 |
| Non-medical costs ($) | 12 | 30.1 | 56.6 | 7.0 | 21 | 136.3 | 434.0 | 5.0 | 2 | 47.5 | 46.0 | 47.5 | 5 | 48.0 | 61.4 | 40.0 |

### c. By health conditions

| | Adult Sample | | | | | | | | Child Sample | | | | | | | |
|---|---|---|---|---|---|---|---|---|---|---|---|---|---|---|---|---|
| | Health conditions* | | | | None of these | | | | Health conditions* | | | | None of these | | | |
| | N | Mean | SD | Median | N | Mean | SD | Median | N | Mean | SD | Median | N | Mean | SD | Median |
| **Primary care** | | | | | | | | | | | | | | | | |
| OOP medical costs ($) | 21 | 17.5 | 30.3 | 3.0 | 65 | 60.7 | 129.1 | 10.0 | 5 | 77.2 | 127.5 | 25.0 | 28 | 33.6 | 49.2 | 20.0 |
| Non-medical costs ($) | 21 | 13.4 | 16.9 | 10.0 | 67 | 23.3 | 48.9 | 7.0 | 5 | 19.0 | 15.2 | 10.0 | 30 | 16.0 | 24.3 | 10.0 |
| **Urgent care** | | | | | | | | | | | | | | | | |
| OOP medical costs ($) | 27 | 60.6 | 144.4 | 5.0 | 76 | 88.4 | 350.1 | 20.0 | 3 | 43.3 | 40.4 | 50.0 | 13 | 57.3 | 82.9 | 20.0 |
| Non-medical costs ($) | 27 | 35.6 | 100.3 | 10.0 | 76 | 19.7 | 41.9 | 2.5 | 3 | 5.0 | 5.0 | 5.0 | 16 | 26.2 | 44.6 | 5.0 |
| **Emergency department** | | | | | | | | | | | | | | | | |
| OOP medical costs ($) | 22 | 81.9 | 173.8 | 5.0 | 38 | 115.5 | 216.0 | 0.0 | 1 | 0.0 | - | 0.0 | 11 | 88.2 | 151.4 | 0.0 |
| Non-medical costs ($) | 22 | 21.5 | 44.0 | 5.0 | 39 | 24.2 | 47.4 | 0.0 | 1 | 20.0 | - | 20.0 | 12 | 35.6 | 70.0 | 11.0 |
| **Hospitalization** | | | | | | | | | | | | | | | | |
| OOP medical costs ($) | 12 | 104.7 | 285.3 | 4.0 | 28 | 292.7 | 469.7 | 65.0 | 0 | - | - | - | 8 | 132.4 | 179.0 | 75.0 |
| Non-medical costs ($) | 12 | 35.3 | 85.5 | 4.5 | 29 | 148.5 | 429.7 | 20.0 | 0 | - | - | - | 8 | 41.9 | 52.4 | 27.5 |

* Includes chronic obstructive pulmonary disease (COPD), heart disease, diabetes, immunocompromised, kidney disease, and liver disease

### d. By Long COVID status

| | Adult Sample | | | | | | | | Child Sample | | | | | | | |
|---|---|---|---|---|---|---|---|---|---|---|---|---|---|---|---|---|
| | Yes | | | | No | | | | Yes | | | | No | | | |
| | N | Mean | SD | Median | N | Mean | SD | Median | N | Mean | SD | Median | N | Mean | SD | Median |
| **Primary care** | | | | | | | | | | | | | | | | |
| OOP medical costs ($) | 53 | 72.9 | 140.4 | 15.0 | 33 | 13.5 | 24.5 | 0.0 | 9 | 55.0 | 69.7 | 25.0 | 22 | 36.6 | 67.4 | 20.0 |
| Non-medical costs ($) | 55 | 29.5 | 52.8 | 10.0 | 33 | 6.7 | 11.0 | 0.0 | 9 | 35.6 | 35.4 | 20.0 | 24 | 10.2 | 12.4 | 7.5 |
| **Urgent care** | | | | | | | | | | | | | | | | |
| OOP medical costs ($) | 52 | 76.4 | 136.3 | 15.0 | 52 | 86.2 | 415.4 | 7.0 | 7 | 87.1 | 105.6 | 10.0 | 7 | 34.3 | 29.4 | 40.0 |
| Non-medical costs ($) | 52 | 39.1 | 84.5 | 10.0 | 52 | 8.2 | 13.9 | 0.0 | 8 | 23.8 | 41.4 | 0.0 | 9 | 27.2 | 47.2 | 10.0 |
| **Emergency department** | | | | | | | | | | | | | | | | |
| OOP medical costs ($) | 39 | 124.1 | 202.7 | 10.0 | 21 | 64.4 | 195.7 | 0.0 | 7 | 114.3 | 186.4 | 0.0 | 4 | 42.5 | 50.6 | 35.0 |
| Non-medical costs ($) | 39 | 31.7 | 55.0 | 5.0 | 22 | 8.2 | 12.8 | 0.0 | 7 | 24.3 | 28.1 | 15.0 | 5 | 55.4 | 106.3 | 12.0 |
| **Hospitalization** | | | | | | | | | | | | | | | | |
| OOP medical costs ($) | 31 | 246.5 | 441.9 | 50.0 | 9 | 200.9 | 399.5 | 0.0 | 6 | 168.2 | 196.1 | 104.5 | 1 | 50.0 | . | 50.0 |
| Non-medical costs ($) | 32 | 139.8 | 411.0 | 9.0 | 9 | 28.6 | 65.2 | 0.0 | 6 | 55.8 | 53.9 | 45.0 | 1 | 0.0 | . | 0.0 |

### Table 8. Healthcare utilization costs, by care setting, and demographics, no exclusions

#### a. By insurance coverage

| | Adult Sample | | | | | | | | Child Sample | | | | | | | |
|---|---|---|---|---|---|---|---|---|---|---|---|---|---|---|---|---|
| | Insured | | | | Uninsured | | | | Insured | | | | Uninsured | | | |
| | N | Mean | SD | Median | N | Mean | SD | Median | N | Mean | SD | Median | N | Mean | SD | Median |
| **Primary care** | | | | | | | | | | | | | | | | |
| OOP medical costs ($) | 198 | 80.2 | 314.5 | 10.0 | 20 | 79.9 | 142.5 | 22.5 | 57 | 41.8 | 68.6 | 20.0 | 2 | 12.5 | 17.7 | 12.5 |
| Non-medical costs ($) | 201 | 71.9 | 403.1 | 10.0 | 21 | 38.6 | 67.0 | 10.0 | 56 | 26.2 | 52.7 | 10.0 | 2 | 15.0 | 21.2 | 15.0 |
| **Urgent care** | | | | | | | | | | | | | | | | |
| OOP medical costs ($) | 214 | 93.1 | 383.5 | 14.0 | 32 | 82.5 | 224.1 | 16.0 | 35 | 41.9 | 66.5 | 20.0 | 2 | 32.5 | 10.6 | 32.5 |
| Non-medical costs ($) | 215 | 38.6 | 168.5 | 10.0 | 32 | 36.2 | 63.3 | 12.5 | 38 | 13.8 | 22.3 | 9.0 | 2 | 0.0 | 0.0 | 0.0 |
| **Emergency department** | | | | | | | | | | | | | | | | |
| OOP medical costs ($) | 140 | 138.9 | 448.9 | 10.0 | 27 | 92.5 | 194.3 | 10.0 | 26 | 81.9 | 168.2 | 0.0 | 1 | 0.0 | . | 0.0 |
| Non-medical costs ($) | 141 | 54.6 | 228.7 | 8.0 | 28 | 15.6 | 17.3 | 9.5 | 28 | 47.4 | 114.2 | 10.0 | 1 | 0.0 | . | 0.0 |
| **Hospitalization** | | | | | | | | | | | | | | | | |
| OOP medical costs ($) | 119 | 206.6 | 790.3 | 10.0 | 22 | 205.4 | 454.3 | 15.0 | 18 | 171.9 | 360.6 | 36.5 | 1 | 0.0 | . | 0.0 |
| Non-medical costs ($) | 120 | 99.0 | 379.1 | 10.0 | 22 | 87.4 | 272.7 | 17.5 | 21 | 58.5 | 91.4 | 20.0 | 1 | 0.0 | . | 0.0 |

#### b. By insurance type

| | Adult Sample | | | | | | | | Child Sample | | | | | | | |
|---|---|---|---|---|---|---|---|---|---|---|---|---|---|---|---|---|
| | Medicaid | | | | Other | | | | Medicaid | | | | Other | | | |
| | N | Mean | SD | Median | N | Mean | SD | Median | N | Mean | SD | Median | N | Mean | SD | Median |
| **Primary care** | | | | | | | | | | | | | | | | |
| OOP medical costs ($) | 68 | 79.8 | 380.5 | 8.0 | 128 | 81.7 | 277.4 | 12.5 | 22 | 14.1 | 29.7 | 0.0 | 34 | 60.3 | 80.9 | 25.0 |
| Non-medical costs ($) | 69 | 106.9 | 533.0 | 10.0 | 130 | 54.4 | 318.1 | 10.0 | 21 | 39.0 | 78.1 | 10.0 | 34 | 19.1 | 27.5 | 10.0 |
| **Urgent care** | | | | | | | | | | | | | | | | |
| OOP medical costs ($) | 87 | 51.5 | 183.8 | 10.0 | 123 | 100.8 | 402.8 | 30.0 | 12 | 23.3 | 56.6 | 0.5 | 23 | 51.5 | 70.3 | 30.0 |
| Non-medical costs ($) | 87 | 26.8 | 65.1 | 10.0 | 124 | 47.8 | 215.0 | 8.5 | 13 | 10.8 | 18.9 | 10.0 | 25 | 15.4 | 24.1 | 8.0 |
| **Emergency department** | | | | | | | | | | | | | | | | |
| OOP medical costs ($) | 73 | 96.3 | 405.4 | 3.0 | 66 | 188.1 | 494.1 | 12.5 | 15 | 37.7 | 144.5 | 0.0 | 11 | 142.3 | 185.9 | 70.0 |
| Non-medical costs ($) | 74 | 41.8 | 176.8 | 5.0 | 66 | 69.8 | 277.7 | 10.0 | 16 | 38.0 | 93.4 | 8.5 | 12 | 59.8 | 140.7 | 13.5 |
| **Hospitalization** | | | | | | | | | | | | | | | | |
| OOP medical costs ($) | 56 | 71.4 | 334.1 | 10.0 | 63 | 326.9 | 1028.8 | 10.0 | 7 | 73.1 | 127.2 | 20.0 | 11 | 234.7 | 447.4 | 40.0 |
| Non-medical costs ($) | 57 | 42.2 | 135.1 | 10.0 | 63 | 150.3 | 503.7 | 10.0 | 8 | 58.8 | 56.0 | 50.0 | 13 | 58.3 | 110.0 | 14.0 |

### c. By health conditions

| | Adult Sample | | | | | | | | Child Sample | | | | | | | |
|---|---|---|---|---|---|---|---|---|---|---|---|---|---|---|---|---|
| | Health conditions* | | | | None of these | | | | Health conditions* | | | | None of these | | | |
| | N | Mean | SD | Median | N | Mean | SD | Median | N | Mean | SD | Median | N | Mean | SD | Median |
| **Primary care** | | | | | | | | | | | | | | | | |
| OOP medical costs ($) | 73 | 123.7 | 495.7 | 10.0 | 147 | 57.3 | 116.4 | 10.0 | 9 | 46.2 | 97.8 | 0.0 | 50 | 39.8 | 62.1 | 20.0 |
| Non-medical costs ($) | 73 | 154.0 | 660.5 | 10.0 | 150 | 26.6 | 52.9 | 9.5 | 13 | 56.8 | 96.3 | 20.0 | 53 | 21.2 | 35.3 | 10.0 |
| **Urgent care** | | | | | | | | | | | | | | | | |
| OOP medical costs ($) | 84 | 133.6 | 512.5 | 10.0 | 162 | 68.9 | 259.8 | 15.0 | 7 | 28.6 | 31.8 | 20.0 | 32 | 42.2 | 68.5 | 20.0 |
| Non-medical costs ($) | 84 | 66.6 | 263.4 | 10.0 | 163 | 25.5 | 50.5 | 7.0 | 16 | 10.0 | 9.1 | 10.0 | 38 | 19.5 | 33.0 | 9.0 |
| **Emergency department** | | | | | | | | | | | | | | | | |
| OOP medical costs ($) | 75 | 141.4 | 424.8 | 10.0 | 93 | 122.5 | 412.9 | 8.0 | 6 | 83.3 | 204.1 | 0.0 | 21 | 77.6 | 159.0 | 1.0 |
| Non-medical costs ($) | 76 | 83.1 | 307.3 | 9.0 | 94 | 19.5 | 35.2 | 7.5 | 15 | 55.0 | 125.0 | 20.0 | 26 | 47.3 | 90.5 | 11.0 |
| **Hospitalization** | | | | | | | | | | | | | | | | |
| OOP medical costs ($) | 71 | 234.5 | 979.7 | 10.0 | 71 | 175.6 | 392.2 | 11.0 | 5 | 314.0 | 663.2 | 20.0 | 14 | 108.9 | 158.0 | 36.5 |
| Non-medical costs ($) | 71 | 108.7 | 432.1 | 10.0 | 72 | 84.5 | 280.5 | 17.5 | 13 | 56.5 | 108.6 | 20.0 | 18 | 38.6 | 47.3 | 22.5 |

* Includes chronic obstructive pulmonary disease (COPD), heart disease, diabetes, immunocompromised, kidney disease, and liver disease

### d. By Long COVID status

| | Adult Sample | | | | | | | | Child Sample | | | | | | | |
|---|---|---|---|---|---|---|---|---|---|---|---|---|---|---|---|---|
| | Yes | | | | No | | | | Yes | | | | No | | | |
| | N | Mean | SD | Median | N | Mean | SD | Median | N | Mean | SD | Median | N | Mean | SD | Median |
| **Primary care** | | | | | | | | | | | | | | | | |
| OOP medical costs ($) | 129 | 115.3 | 384.6 | 15.0 | 93 | 28.0 | 77.2 | 0.0 | 16 | 41.2 | 59.2 | 10.0 | 37 | 40.3 | 71.3 | 20.0 |
| Non-medical costs ($) | 132 | 105.9 | 494.7 | 10.0 | 94 | 13.7 | 34.7 | 5.0 | 21 | 28.9 | 30.7 | 20.0 | 39 | 31.2 | 66.3 | 10.0 |
| **Urgent care** | | | | | | | | | | | | | | | | |
| OOP medical costs ($) | 140 | 104.2 | 398.6 | 19.5 | 112 | 71.4 | 311.6 | 10.0 | 12 | 55.8 | 87.3 | 15.0 | 22 | 27.5 | 25.1 | 24.5 |
| Non-medical costs ($) | 140 | 59.2 | 208.3 | 10.0 | 113 | 13.0 | 31.6 | 5.0 | 24 | 15.9 | 24.6 | 10.0 | 25 | 20.3 | 33.6 | 10.0 |
| **Emergency department** | | | | | | | | | | | | | | | | |
| OOP medical costs ($) | 110 | 155.2 | 478.4 | 10.0 | 59 | 83.4 | 260.2 | 0.0 | 16 | 119.3 | 206.1 | 2.0 | 10 | 22.1 | 37.3 | 0.0 |
| Non-medical costs ($) | 111 | 62.3 | 254.0 | 10.0 | 60 | 20.6 | 58.9 | 0.0 | 27 | 52.1 | 114.9 | 15.0 | 13 | 49.8 | 80.7 | 12.0 |
| **Hospitalization** | | | | | | | | | | | | | | | | |
| OOP medical costs ($) | 109 | 239.8 | 837.1 | 11.0 | 33 | 90.2 | 239.2 | 4.0 | 13 | 224.3 | 416.5 | 25.0 | 3 | 43.3 | 5.8 | 40.0 |
| Non-medical costs ($) | 110 | 117.5 | 410.3 | 10.0 | 33 | 26.6 | 61.6 | 6.0 | 24 | 51.4 | 86.3 | 20.0 | 4 | 43.8 | 41.5 | 37.5 |

Table 9. Financial challenges resulting from most recent COVID-19 illness, primary analysis, n(%)

a. By insurance coverage status

| | Adult Sample | | Child Sample | |
|---|---|---|---|---|
| | Insured (n=358) | Uninsured (n=45) | Insured (n=46) | Uninsured (n=2) |
| **Problems paying COVID-19-related medical bills** | | | | |
| No | 318 (89.1) | 36 (80.0) | 41 (89.1) | 2 (100.0) |
| Yes, when I had COVID-19 | 25 (7.0) | 4 (8.9) | 2 (4.3) | 0 (0.0) |
| Yes, after I had COVID-19 | 10 (2.8) | 4 (8.9) | 1 (2.2) | 0 (0.0) |
| Yes, currently | 9 (2.5) | 1 (2.2) | 2 (4.3) | 0 (0.0) |
| **Wait to get medical care due to cost** | | | | |
| No | 306 (85.5) | 29 (64.4) | 42 (91.3) | 2 (100.0) |
| Yes, when I had COVID-19 | 30 (8.4) | 11 (24.4) | 2 (4.3) | 0 (0.0) |
| Yes, after I had COVID-19 | 11 (3.1) | 5 (11.1) | 1 (2.2) | 0 (0.0) |
| Yes, currently | 16 (4.5) | 2 (4.4) | 3 (6.5) | 0 (0.0) |
| **Take less or skip doses of medicine due to cost** | | | | |
| No | 323 (90.5) | 36 (80.0) | 43 (93.5) | 2 (100.0) |
| Yes, when I had COVID-19 | 17 (4.8) | 5 (11.1) | 1 (2.2) | 0 (0.0) |
| Yes, after I had COVID-19 | 13 (3.6) | 2 (4.4) | 1 (2.2) | 0 (0.0) |
| Yes, currently | 7 (2.0) | 2 (4.4) | 3 (6.5) | 0 (0.0) |
| **Did not get medical care due to cost** | | | | |
| No | 304 (84.9) | 35 (77.8) | 41 (89.1) | 1 (50.0) |
| Yes, when I had COVID-19 | 24 (6.7) | 4 (8.9) | 0 (0.0) | 0 (0.0) |
| Yes, after I had COVID-19 | 18 (5.0) | 4 (8.9) | 2 (4.3) | 0 (0.0) |
| Yes, currently | 16 (4.5) | 2 (4.4) | 4 (8.7) | 1 (50.0) |
| **Did not get prescription medicine due to cost** | | | | |
| No | 308 (86.0) | 38 (84.4) | 42 (91.3) | 1 (50.0) |
| Yes, when I had COVID-19 | 23 (6.4) | 3 (6.7) | 1 (2.2) | 0 (0.0) |
| Yes, after I had COVID-19 | 18 (5.0) | 2 (4.4) | 0 (0.0) | 0 (0.0) |
| Yes, currently | 14 (3.9) | 2 (4.4) | 3 (6.5) | 1 (50.0) |
| **Problems paying non-medical bills** | | | | |
| No | 290 (81.2) | 34 (75.6) | 36 (78.3) | 1 (50.0) |
| Yes, when I had COVID-19 | 27 (7.6) | 5 (11.1) | 4 (8.7) | 0 (0.0) |
| Yes, after I had COVID-19 | 19 (5.3) | 2 (4.4) | 2 (4.3) | 0 (0.0) |
| Yes, currently | 40 (11.2) | 4 (8.9) | 7 (15.2) | 1 (50.0) |
| **Skip or limit meals due to cost** | | | | |
| No | 296 (82.7) | 34 (75.6) | 40 (87.0) | 1 (50.0) |
| Yes, when I had COVID-19 | 24 (6.7) | 6 (13.3) | 2 (4.3) | 0 (0.0) |
| Yes, after I had COVID-19 | 18 (5.0) | 3 (6.7) | 2 (4.3) | 0 (0.0) |
| Yes, currently | 34 (9.5) | 3 (6.7) | 4 (8.7) | 1 (50.0) |

b. By insurance type

| | Adult Sample | | Child Sample | |
|---|---|---|---|---|
| | Medicaid (n=79) | Other (n=272) | Medicaid (n=16) | Other (n=29) |
| **Problems paying COVID-19-related medical bills** | | | | |
| No | 64 (82.1) | 247 (90.8) | 14 (87.5) | 26 (89.7) |
| Yes, when I had COVID-19 | 6 (7.7) | 19 (7.0) | 1 (6.2) | 1 (3.4) |
| Yes, after I had COVID-19 | 3 (3.8) | 7 (2.6) | 1 (6.2) | 0 (0.0) |
| Yes, currently | 7 (9.0) | 2 (0.7) | 0 (0.0) | 2 (6.9) |
| **Wait to get medical care due to cost** | | | | |
| No | 61 (77.2) | 240 (88.2) | 14 (87.5) | 27 (93.1) |
| Yes, when I had COVID-19 | 9 (11.4) | 20 (7.4) | 2 (12.5) | 0 (0.0) |
| Yes, after I had COVID-19 | 2 (2.5) | 9 (3.3) | 1 (6.2) | 0 (0.0) |
| Yes, currently | 7 (8.9) | 8 (2.9) | 1 (6.2) | 2 (6.9) |
| **Take less or skip doses of medicine due to cost** | | | | |
| No | 64 (82.1) | 252 (92.6) | 15 (93.8) | 27 (93.1) |
| Yes, when I had COVID-19 | 8 (10.3) | 9 (3.3) | 1 (6.2) | 0 (0.0) |
| Yes, after I had COVID-19 | 3 (3.8) | 10 (3.7) | 1 (6.2) | 0 (0.0) |
| Yes, currently | 5 (6.4) | 2 (0.7) | 1 (6.2) | 2 (6.9) |
| **Did not get medical care due to cost** | | | | |
| No | 62 (78.5) | 237 (87.1) | 15 (93.8) | 25 (86.2) |
| Yes, when I had COVID-19 | 9 (11.4) | 14 (5.1) | 0 (0.0) | 0 (0.0) |
| Yes, after I had COVID-19 | 4 (5.1) | 14 (5.1) | 1 (6.2) | 1 (3.4) |
| Yes, currently | 5 (6.3) | 10 (3.7) | 1 (6.2) | 3 (10.3) |
| **Did not get prescription medicine due to cost** | | | | |
| No | 61 (77.2) | 241 (88.6) | 14 (87.5) | 27 (93.1) |
| Yes, when I had COVID-19 | 9 (11.4) | 14 (5.1) | 1 (6.2) | 0 (0.0) |
| Yes, after I had COVID-19 | 5 (6.3) | 13 (4.8) | 0 (0.0) | 0 (0.0) |
| Yes, currently | 6 (7.6) | 7 (2.6) | 1 (6.2) | 2 (6.9) |
| **Problems paying non-medical bills** | | | | |
| No | 53 (67.9) | 231 (84.9) | 11 (68.8) | 24 (82.8) |
| Yes, when I had COVID-19 | 14 (17.9) | 13 (4.8) | 4 (25.0) | 0 (0.0) |
| Yes, after I had COVID-19 | 9 (11.5) | 10 (3.7) | 2 (12.5) | 0 (0.0) |
| Yes, currently | 16 (20.5) | 23 (8.5) | 2 (12.5) | 5 (17.2) |
| **Skip or limit meals due to cost** | | | | |
| No | 52 (65.8) | 238 (87.5) | 13 (81.2) | 26 (89.7) |
| Yes, when I had COVID-19 | 12 (15.2) | 11 (4.0) | 1 (6.2) | 1 (3.4) |
| Yes, after I had COVID-19 | 9 (11.4) | 9 (3.3) | 2 (12.5) | 0 (0.0) |
| Yes, currently | 18 (22.8) | 16 (5.9) | 2 (12.5) | 2 (6.9) |

#### c. By health conditions

| | Adult Sample | | Child sample | |
|---|---|---|---|---|
| | Health conditions* (n=106) | None of these (n=299) | Health conditions* (n=9) | None of these (n=43) |
| **Problems paying COVID-19-related medical bills** | | | | |
| No | 88 (83.0) | 269 (90.3) | 9 (100.0) | 37 (86.0) |
| Yes, when I had COVID-19 | 10 (9.4) | 18 (6.0) | 0 (0.0) | 3 (7.0) |
| Yes, after I had COVID-19 | 4 (3.8) | 10 (3.4) | 0 (0.0) | 1 (2.3) |
| Yes, currently | 6 (5.7) | 4 (1.3) | 0 (0.0) | 2 (4.7) |
| **Wait to get medical care due to cost** | | | | |
| No | 87 (82.1) | 251 (83.9) | 8 (88.9) | 39 (90.7) |
| Yes, when I had COVID-19 | 8 (7.5) | 32 (10.7) | 1 (11.1) | 2 (4.7) |
| Yes, after I had COVID-19 | 5 (4.7) | 11 (3.7) | 1 (11.1) | 0 (0.0) |
| Yes, currently | 8 (7.5) | 10 (3.3) | 1 (11.1) | 2 (4.7) |
| **Take less or skip doses of medicine due to cost** | | | | |
| No | 90 (84.9) | 272 (91.3) | 8 (88.9) | 40 (93.0) |
| Yes, when I had COVID-19 | 5 (4.7) | 16 (5.4) | 1 (11.1) | 1 (2.3) |
| Yes, after I had COVID-19 | 6 (5.7) | 9 (3.0) | 1 (11.1) | 0 (0.0) |
| Yes, currently | 7 (6.6) | 2 (0.7) | 1 (11.1) | 2 (4.7) |
| **Did not get medical care due to cost** | | | | |
| No | 88 (83.0) | 254 (84.9) | 6 (66.7) | 39 (90.7) |
| Yes, when I had COVID-19 | 10 (9.4) | 18 (6.0) | 0 (0.0) | 1 (2.3) |
| Yes, after I had COVID-19 | 3 (2.8) | 18 (6.0) | 1 (11.1) | 1 (2.3) |
| Yes, currently | 6 (5.7) | 12 (4.0) | 3 (33.3) | 2 (4.7) |
| **Did not get prescription medicine due to cost** | | | | |
| No | 85 (80.2) | 265 (88.6) | 6 (66.7) | 40 (93.0) |
| Yes, when I had COVID-19 | 7 (6.6) | 18 (6.0) | 0 (0.0) | 2 (4.7) |
| Yes, after I had COVID-19 | 10 (9.4) | 9 (3.0) | 0 (0.0) | 0 (0.0) |
| Yes, currently | 7 (6.6) | 9 (3.0) | 3 (33.3) | 1 (2.3) |
| **Problems paying non-medical bills** | | | | |
| No | 78 (74.3) | 247 (82.6) | 5 (55.6) | 35 (81.4) |
| Yes, when I had COVID-19 | 8 (7.6) | 25 (8.4) | 2 (22.2) | 3 (7.0) |
| Yes, after I had COVID-19 | 7 (6.7) | 14 (4.7) | 1 (11.1) | 1 (2.3) |
| Yes, currently | 14 (13.3) | 30 (10.0) | 3 (33.3) | 5 (11.6) |
| **Skip or limit meals due to cost** | | | | |
| No | 79 (74.5) | 253 (84.6) | 7 (77.8) | 36 (83.7) |
| Yes, when I had COVID-19 | 8 (7.5) | 23 (7.7) | 1 (11.1) | 3 (7.0) |
| Yes, after I had COVID-19 | 11 (10.4) | 10 (3.3) | 1 (11.1) | 1 (2.3) |
| Yes, currently | 15 (14.2) | 21 (7.0) | 2 (22.2) | 3 (7.0) |

* Includes chronic obstructive pulmonary disease (COPD), heart disease, diabetes, immunocompromised, kidney disease, and liver disease

#### d. By Long COVID status

| | Adult Sample | | Child Sample | |
|---|---|---|---|---|
| | **Yes (n=144)** | **No (n=264)** | **Yes (n=13)** | **No (n=36)** |
| **Problems paying COVID-19-related medical bills** | | | | |
| No | 107 (74.8) | 252 (95.5) | 11 (84.6) | 32 (88.9) |
| Yes, when I had COVID-19 | 21 (14.7) | 8 (3.0) | 2 (15.4) | 1 (2.8) |
| Yes, after I had COVID-19 | 11 (7.7) | 3 (1.1) | 0 (0.0) | 1 (2.8) |
| Yes, currently | 9 (6.3) | 1 (0.4) | 0 (0.0) | 2 (5.6) |
| **Wait to get medical care due to cost** | | | | |
| No | 96 (66.7) | 244 (92.4) | 11 (84.6) | 33 (91.7) |
| Yes, when I had COVID-19 | 30 (20.8) | 11 (4.2) | 2 (15.4) | 1 (2.8) |
| Yes, after I had COVID-19 | 11 (7.6) | 5 (1.9) | 0 (0.0) | 1 (2.8) |
| Yes, currently | 13 (9.0) | 5 (1.9) | 0 (0.0) | 3 (8.3) |
| **Take less or skip doses of medicine due to cost** | | | | |
| No | 113 (79.0) | 251 (95.1) | 12 (92.3) | 33 (91.7) |
| Yes, when I had COVID-19 | 16 (11.2) | 6 (2.3) | 1 (7.7) | 1 (2.8) |
| Yes, after I had COVID-19 | 12 (8.4) | 3 (1.1) | 0 (0.0) | 1 (2.8) |
| Yes, currently | 5 (3.5) | 4 (1.5) | 0 (0.0) | 3 (8.3) |
| **Did not get medical care due to cost** | | | | |
| No | 101 (70.1) | 242 (91.7) | 12 (92.3) | 30 (83.3) |
| Yes, when I had COVID-19 | 21 (14.6) | 8 (3.0) | 1 (7.7) | 0 (0.0) |
| Yes, after I had COVID-19 | 14 (9.7) | 8 (3.0) | 0 (0.0) | 2 (5.6) |
| Yes, currently | 11 (7.6) | 7 (2.7) | 0 (0.0) | 5 (13.9) |
| **Did not get prescription medicine due to cost** | | | | |
| No | 102 (70.8) | 249 (94.3) | 11 (84.6) | 32 (88.9) |
| Yes, when I had COVID-19 | 21 (14.6) | 5 (1.9) | 2 (15.4) | 0 (0.0) |
| Yes, after I had COVID-19 | 16 (11.1) | 4 (1.5) | 0 (0.0) | 0 (0.0) |
| Yes, currently | 10 (6.9) | 6 (2.3) | 0 (0.0) | 4 (11.1) |
| **Problems paying non-medical bills** | | | | |
| No | 95 (66.0) | 233 (88.6) | 11 (84.6) | 26 (72.2) |
| Yes, when I had COVID-19 | 24 (16.7) | 9 (3.4) | 2 (15.4) | 3 (8.3) |
| Yes, after I had COVID-19 | 14 (9.7) | 7 (2.7) | 0 (0.0) | 2 (5.6) |
| Yes, currently | 23 (16.0) | 21 (8.0) | 0 (0.0) | 8 (22.2) |
| **Skip or limit meals due to cost** | | | | |
| No | 97 (67.4) | 237 (89.8) | 12 (92.3) | 28 (77.8) |
| Yes, when I had COVID-19 | 21 (14.6) | 10 (3.8) | 1 (7.7) | 3 (8.3) |
| Yes, after I had COVID-19 | 14 (9.7) | 7 (2.7) | 0 (0.0) | 2 (5.6) |
| Yes, currently | 22 (15.3) | 15 (5.7) | 0 (0.0) | 5 (13.9) |

Table 10. Financial challenges resulting from most recent COVID-19 illness, n(%) (no exclusions)

a. By insurance coverage status

| | Adult Sample | | Child Sample | |
|---|---|---|---|---|
| | Insured (n=678) | Uninsured (n=107) | Insured (n=138) | Uninsured (n=8) |
| **Problems paying COVID-19-related medical bills** | | | | |
| No | 559 (82.6) | 83 (77.6) | 120 (87.0) | 8 (100.0) |
| Yes, when I had COVID-19 | 76 (11.2) | 14 (13.1) | 10 (7.2) | 0 (0.0) |
| Yes, after I had COVID-19 | 27 (4.0) | 7 (6.5) | 2 (1.4) | 0 (0.0) |
| Yes, currently | 26 (3.8) | 6 (5.6) | 6 (4.3) | 0 (0.0) |
| **Wait to get medical care due to cost** | | | | |
| No | 545 (80.6) | 72 (67.3) | 120 (87.0) | 8 (100.0) |
| Yes, when I had COVID-19 | 75 (11.1) | 20 (18.7) | 7 (5.1) | 0 (0.0) |
| Yes, after I had COVID-19 | 36 (5.3) | 15 (14.0) | 6 (4.3) | 0 (0.0) |
| Yes, currently | 27 (4.0) | 5 (4.7) | 8 (5.8) | 0 (0.0) |
| **Take less or skip doses of medicine due to cost** | | | | |
| No | 567 (83.8) | 79 (73.8) | 123 (89.1) | 7 (87.5) |
| Yes, when I had COVID-19 | 61 (9.0) | 10 (9.3) | 5 (3.6) | 0 (0.0) |
| Yes, after I had COVID-19 | 34 (5.0) | 13 (12.1) | 5 (3.6) | 1 (12.5) |
| Yes, currently | 19 (2.8) | 7 (6.5) | 7 (5.1) | 0 (0.0) |
| **Did not get medical care due to cost** | | | | |
| No | 549 (81.0) | 73 (68.2) | 117 (84.8) | 5 (62.5) |
| Yes, when I had COVID-19 | 62 (9.1) | 14 (13.1) | 4 (2.9) | 1 (12.5) |
| Yes, after I had COVID-19 | 41 (6.0) | 14 (13.1) | 8 (5.8) | 0 (0.0) |
| Yes, currently | 33 (4.9) | 6 (5.6) | 10 (7.2) | 2 (25.0) |
| **Did not get prescription medicine due to cost** | | | | |
| No | 543 (80.1) | 80 (74.8) | 120 (87.0) | 6 (75.0) |
| Yes, when I had COVID-19 | 76 (11.2) | 11 (10.3) | 5 (3.6) | 0 (0.0) |
| Yes, after I had COVID-19 | 34 (5.0) | 8 (7.5) | 7 (5.1) | 1 (12.5) |
| Yes, currently | 33 (4.9) | 9 (8.4) | 7 (5.1) | 1 (12.5) |
| **Problems paying non-medical bills** | | | | |
| No | 515 (76.3) | 70 (65.4) | 102 (73.9) | 5 (62.5) |
| Yes, when I had COVID-19 | 72 (10.7) | 16 (15.0) | 11 (8.0) | 0 (0.0) |
| Yes, after I had COVID-19 | 49 (7.3) | 12 (11.2) | 13 (9.4) | 1 (12.5) |
| Yes, currently | 71 (10.5) | 14 (13.1) | 19 (13.8) | 2 (25.0) |
| **Skip or limit meals due to cost** | | | | |
| No | 522 (77.2) | 75 (70.1) | 112 (81.2) | 4 (50.0) |
| Yes, when I had COVID-19 | 67 (9.9) | 15 (14.0) | 8 (5.8) | 0 (0.0) |
| Yes, after I had COVID-19 | 48 (7.1) | 11 (10.3) | 11 (8.0) | 1 (12.5) |
| Yes, currently | 60 (8.9) | 12 (11.2) | 12 (8.7) | 3 (37.5) |

b. By insurance type

| | Adult Sample | | Child Sample | |
|---|---|---|---|---|
| | Medicaid (n=172) | Other (n=491) | Medicaid (n=52) | Other (n=85) |
| **Problems paying COVID-19-related medical bills** | | | | |
| No | 117 (68.4) | 429 (87.4) | 44 (84.6) | 75 (88.2) |
| Yes, when I had COVID-19 | 27 (15.8) | 47 (9.6) | 4 (7.7) | 6 (7.1) |
| Yes, after I had COVID-19 | 13 (7.6) | 14 (2.9) | 2 (3.8) | 0 (0.0) |
| Yes, currently | 18 (10.5) | 8 (1.6) | 2 (3.8) | 4 (4.7) |
| **Wait to get medical care due to cost** | | | | |
| No | 114 (66.7) | 421 (85.9) | 43 (82.7) | 76 (89.4) |
| Yes, when I had COVID-19 | 29 (17.0) | 42 (8.6) | 3 (5.8) | 4 (4.7) |
| Yes, after I had COVID-19 | 15 (8.8) | 21 (4.3) | 4 (7.7) | 2 (2.4) |
| Yes, currently | 14 (8.2) | 12 (2.4) | 5 (9.6) | 3 (3.5) |
| **Take less or skip doses of medicine due to cost** | | | | |
| No | 117 (68.4) | 437 (89.0) | 44 (84.6) | 78 (91.8) |
| Yes, when I had COVID-19 | 31 (18.1) | 28 (5.7) | 3 (5.8) | 2 (2.4) |
| Yes, after I had COVID-19 | 14 (8.2) | 20 (4.1) | 3 (5.8) | 2 (2.4) |
| Yes, currently | 12 (7.0) | 7 (1.4) | 4 (7.7) | 3 (3.5) |
| **Did not get medical care due to cost** | | | | |
| No | 115 (66.9) | 424 (86.4) | 44 (84.6) | 72 (84.7) |
| Yes, when I had COVID-19 | 28 (16.3) | 31 (6.3) | 3 (5.8) | 1 (1.2) |
| Yes, after I had COVID-19 | 16 (9.3) | 24 (4.9) | 2 (3.8) | 6 (7.1) |
| Yes, currently | 15 (8.7) | 17 (3.5) | 4 (7.7) | 6 (7.1) |
| **Did not get prescription medicine due to cost** | | | | |
| No | 113 (65.7) | 419 (85.3) | 42 (80.8) | 77 (90.6) |
| Yes, when I had COVID-19 | 32 (18.6) | 42 (8.6) | 4 (7.7) | 1 (1.2) |
| Yes, after I had COVID-19 | 13 (7.6) | 20 (4.1) | 4 (7.7) | 3 (3.5) |
| Yes, currently | 16 (9.3) | 16 (3.3) | 3 (5.8) | 4 (4.7) |
| **Problems paying non-medical bills** | | | | |
| No | 100 (59.2) | 404 (82.3) | 32 (61.5) | 69 (81.2) |
| Yes, when I had COVID-19 | 31 (18.3) | 38 (7.7) | 9 (17.3) | 2 (2.4) |
| Yes, after I had COVID-19 | 27 (16.0) | 22 (4.5) | 7 (13.5) | 6 (7.1) |
| Yes, currently | 31 (18.3) | 38 (7.7) | 9 (17.3) | 10 (11.8) |
| **Skip or limit meals due to cost** | | | | |
| No | 99 (58.2) | 412 (83.9) | 36 (69.2) | 75 (88.2) |
| Yes, when I had COVID-19 | 32 (18.8) | 32 (6.5) | 4 (7.7) | 4 (4.7) |
| Yes, after I had COVID-19 | 25 (14.7) | 23 (4.7) | 7 (13.5) | 4 (4.7) |
| Yes, currently | 28 (16.5) | 31 (6.3) | 8 (15.4) | 4 (4.7) |

#### b. By health conditions

| | Adult Sample | | Child Sample | |
|---|---|---|---|---|
| | Health conditions* (n=220) | None of these (n=570) | Health conditions* (n=35) | None of these (n=134) |
| **Problems paying COVID-19-related medical bills** | | | | |
| No | 151 (68.6) | 498 (87.5) | 25 (71.4) | 116 (86.6) |
| Yes, when I had COVID-19 | 42 (19.1) | 47 (8.3) | 4 (11.4) | 11 (8.2) |
| Yes, after I had COVID-19 | 17 (7.7) | 17 (3.0) | 2 (5.7) | 3 (2.2) |
| Yes, currently | 17 (7.7) | 14 (2.5) | 6 (17.1) | 4 (3.0) |
| **Wait to get medical care due to cost** | | | | |
| No | 152 (69.1) | 472 (83.1) | 23 (65.7) | 117 (87.3) |
| Yes, when I had COVID-19 | 34 (15.5) | 60 (10.6) | 2 (5.7) | 7 (5.2) |
| Yes, after I had COVID-19 | 23 (10.5) | 28 (4.9) | 7 (20.0) | 6 (4.5) |
| Yes, currently | 14 (6.4) | 17 (3.0) | 6 (17.1) | 5 (3.7) |
| **Take less or skip doses of medicine due to cost** | | | | |
| No | 155 (70.5) | 499 (87.7) | 23 (65.7) | 119 (88.8) |
| Yes, when I had COVID-19 | 33 (15.0) | 36 (6.3) | 3 (8.6) | 7 (5.2) |
| Yes, after I had COVID-19 | 20 (9.1) | 27 (4.7) | 5 (14.3) | 3 (2.2) |
| Yes, currently | 16 (7.3) | 9 (1.6) | 6 (17.1) | 5 (3.7) |
| **Did not get medical care due to cost** | | | | |
| No | 154 (70.0) | 475 (83.3) | 23 (65.7) | 112 (83.6) |
| Yes, when I had COVID-19 | 35 (15.9) | 41 (7.2) | 4 (11.4) | 6 (4.5) |
| Yes, after I had COVID-19 | 16 (7.3) | 38 (6.7) | 2 (5.7) | 8 (6.0) |
| Yes, currently | 18 (8.2) | 20 (3.5) | 7 (20.0) | 8 (6.0) |
| **Did not get prescription medicine due to cost** | | | | |
| No | 148 (67.3) | 484 (84.9) | 20 (57.1) | 119 (88.8) |
| Yes, when I had COVID-19 | 37 (16.8) | 49 (8.6) | 4 (11.4) | 6 (4.5) |
| Yes, after I had COVID-19 | 19 (8.6) | 21 (3.7) | 3 (8.6) | 7 (5.2) |
| Yes, currently | 20 (9.1) | 20 (3.5) | 8 (22.9) | 3 (2.2) |
| **Problems paying non-medical bills** | | | | |
| No | 136 (62.1) | 452 (79.6) | 17 (48.6) | 102 (76.1) |
| Yes, when I had COVID-19 | 36 (16.4) | 53 (9.3) | 6 (17.1) | 11 (8.2) |
| Yes, after I had COVID-19 | 26 (11.9) | 36 (6.3) | 4 (11.4) | 13 (9.7) |
| Yes, currently | 29 (13.2) | 56 (9.9) | 12 (34.3) | 12 (9.0) |
| **Skip or limit meals due to cost** | | | | |
| No | 144 (65.5) | 458 (80.6) | 18 (51.4) | 109 (81.3) |
| Yes, when I had COVID-19 | 28 (12.7) | 55 (9.7) | 5 (14.3) | 10 (7.5) |
| Yes, after I had COVID-19 | 31 (14.1) | 28 (4.9) | 7 (20.0) | 10 (7.5) |
| Yes, currently | 25 (11.4) | 46 (8.1) | 9 (25.7) | 10 8 (6.0) |

* Includes chronic obstructive pulmonary disease (COPD), heart disease, diabetes, immunocompromised, kidney disease, and liver disease

c. By Long COVID status

| | Adult Sample | | Child Sample | |
|---|---|---|---|---|
| | Yes (n=304) | No (n=495) | Yes (n=46) | No (n=111) |
| **Problems paying COVID-19-related medical bills** | | | | |
| No | 201 (66.3) | 453 (91.5) | 28 (60.9) | 103 (92.8) |
| Yes, when I had COVID-19 | 65 (21.5) | 26 (5.3) | 10 (21.7) | 3 (2.7) |
| Yes, after I had COVID-19 | 27 (8.9) | 8 (1.6) | 3 (6.5) | 2 (1.8) |
| Yes, currently | 23 (7.6) | 9 (1.8) | 7 (15.2) | 3 (2.7) |
| **Wait to get medical care due to cost** | | | | |
| No | 189 (62.4) | 441 (89.3) | 27 (58.7) | 102 (91.9) |
| Yes, when I had COVID-19 | 66 (21.8) | 30 (6.1) | 5 (10.9) | 3 (2.7) |
| Yes, after I had COVID-19 | 35 (11.6) | 16 (3.2) | 11 (23.9) | 2 (1.8) |
| Yes, currently | 23 (7.6) | 9 (1.8) | 5 (10.9) | 6 (5.4) |
| **Take less or skip doses of medicine due to cost** | | | | |
| No | 206 (68.0) | 453 (91.5) | 28 (60.9) | 103 (92.8) |
| Yes, when I had COVID-19 | 49 (16.2) | 23 (4.6) | 7 (15.2) | 2 (1.8) |
| Yes, after I had COVID-19 | 37 (12.2) | 10 (2.0) | 5 (10.9) | 3 (2.7) |
| Yes, currently | 16 (5.3) | 10 (2.0) | 6 (13.0) | 5 (4.5) |
| **Did not get medical care due to cost** | | | | |
| No | 195 (64.1) | 438 (88.5) | 27 (58.7) | 97 (87.4) |
| Yes, when I had COVID-19 | 56 (18.4) | 22 (4.4) | 8 (17.4) | 1 (0.9) |
| Yes, after I had COVID-19 | 34 (11.2) | 22 (4.4) | 6 (13.0) | 4 (3.6) |
| Yes, currently | 25 (8.2) | 14 (2.8) | 5 (10.9) | 10 (9.0) |
| **Did not get prescription medicine due to cost** | | | | |
| No | 189 (62.2) | 447 (90.3) | 27 (58.7) | 101 (91.0) |
| Yes, when I had COVID-19 | 65 (21.4) | 23 (4.6) | 10 (21.7) | 0 (0.0) |
| Yes, after I had COVID-19 | 32 (10.5) | 10 (2.0) | 6 (13.0) | 3 (2.7) |
| Yes, currently | 26 (8.6) | 16 (3.2) | 4 (8.7) | 7 (6.3) |
| **Problems paying non-medical bills** | | | | |
| No | 172 (56.8) | 422 (85.6) | 24 (52.2) | 86 (77.5) |
| Yes, when I had COVID-19 | 62 (20.5) | 28 (5.7) | 11 (23.9) | 6 (5.4) |
| Yes, after I had COVID-19 | 48 (15.8) | 15 (3.0) | 7 (15.2) | 9 (8.1) |
| Yes, currently | 47 (15.5) | 39 (7.9) | 6 (13.0) | 16 (14.4) |
| **Skip or limit meals due to cost** | | | | |
| No | 174 (57.4) | 433 (87.7) | 26 (56.5) | 92 (82.9) |
| Yes, when I had COVID-19 | 59 (19.5) | 25 (5.1) | 10 (21.7) | 4 (3.6) |
| Yes, after I had COVID-19 | 46 (15.2) | 14 (2.8) | 11 (23.9) | 5 (4.5) |
| Yes, currently | 45 (14.9) | 28 (5.7) | 3 (6.5) | 13 (11.7) |

### Table 11. Comparison of demographic characteristics by analysis, n(%)

| | No exclusions | | Primary analysis | | Secondary analysis | |
|---|---|---|---|---|---|---|
| | Adult sample (n=800) | Child sample (n=169) | Adult sample (n=409) | Child sample (n=52) | Adult sample (n=232) | Child sample (n=36) |
| Female | 400 (50.0) | 106 (62.7) | 221 (54.0) | 28 (53.9) | 132 (56.9) | 18 (50) |
| Age of respondent (mean (SD)) | 46.5 (17.6) | 40.3 (10.0) | 49.6 (17.9) | 41.2 (10.4) | 50.4 (17.6) | 40.5 (9.6) |
| Age of patient (mean (SD)) | NA | 8.9 (5.4) | NA | 9 (5.2) | NA | 9.1 (5.1) |
| Hispanic | 169 (20.0) | 33 (19.5) | 76 (18.6) | 15 (28.9) | 41 (17.7) | 12 (33.3) |
| Race* | | | | | | |
| White | 611 (76.4) | 112 (66.3) | 325 (79.5) | 34 (65.4) | 184 (79.3) | 23 (63.9) |
| Black | 137 (17.1) | 41 (24.3) | 58 (14.2) | 12 (23.1) | 36 (15.5) | 8 (22.2) |
| Other† | 73 (9.1) | 28 (16.6) | 38 (9.3) | 9 (17.3) | 18 (7.8) | 5 (13.9) |
| Education level | | | | | | |
| Less than high school | 20 (2.5) | 2 (1.2) | 5 (1.2) | 1 (2.0) | 2 (0.9) | 1 (2.8) |
| High school or GED | 315 (39.4) | 65 (38.7) | 174 (42.7) | 22 (43.1) | 99 (42.9) | 15 (41.7) |
| Bachelor's degree | 294 (36.8) | 64 (38.1) | 150 (36.8) | 18 (35.3) | 87 (37.7) | 12 (33.3) |
| Master's degree | 131 (16.4) | 25 (14.9) | 62 (15.2) | 7 (13.7) | 32 (13.9) | 6 (16.7) |
| Doctoral/professional degree | 38 4.8) | 12 (7.1) | 17 (4.2) | 3 (5.9) | 11 (4.8) | 2 (5.6) |
| Missing | - | - | - | - | - | - |
| Geographic area | | | | | | |
| Urban | 298 (37.3) | 67 (39.6) | 119 (29.2) | 20 (38.5) | 68 (29.4) | 17 (47.2) |
| Suburban | 356 (44.5) | 78 (46.2) | 214 (52.5) | 24 (46.2) | 117 (50.7) | 13 (36.1) |
| Rural | 144 (18.0) | 24 (14.2) | 75 (18.4) | 8 (15.4) | 46 (19.9) | 6 (16.7) |
| Missing | 2 (0.2) | - | - | - | - | - |
| Household income | | | | | | |
| <$40,000 | 188 (23.5) | 39 (23.1) | 98 (24.0) | 8 (15.4) | 51 (22.0) | 4 (11.1) |
| $40,000 - <$80,000 | 254 (31.8) | 53 (31.4) | 145 (35.5) | 20 (38.5) | 92 (39.7) | 14 (38.9) |
| $80,000 - <$150,000 | 208 (26.0) | 40 (23.7) | 89 (21.8) | 14 (26.9) | 49 (21.1) | 10 (27.8) |
| ≥$150,000 | 137 (17.1) | 37 (21.9) | 70 (17.1) | 10 (19.2) | 39 (16.8) | 8 (22.2) |
| Missing | 13 (1.6) | | 7 (1.7) | - | 1 (0.4) | - |
| Insurance* | | | | | | |
| Any insurance | 678 (84.8) | 138 (93.2) | 358 (87.5) | 46 (95.8) | 215 (92.3) | 34 (97.1) |
| Private insurance | | | | | | |
| Employer | 369 (55.1) | 76 (55.5) | 170 (47.8) | 23 (51.1) | 97 (45.3) | 20 (60.6) |
| From insurance company | 141 (21.1) | 21 (15.4) | 53 (15.1) | 5 (11.1) | 31 (14.8) | 3 (9.1) |
| From insurance marketplace | 131 (19.8) | 16 (11.9) | 52 (15.0) | 3 (6.7) | 32 (15.4) | 2 (6.1) |
| Medicare | 240 (35.9) | 8 (5.9) | 127 (36.4) | 1 (2.2) | 78 (37.1) | 1 (2.9) |
| Medicaid | 172 (26.0) | 52 (38.2) | 79 (22.7) | 16 (34.8) | 41 (19.7) | 8 (23.5) |
| Tricare | 62 (9.4) | 9 (6.6) | 14 (4.0) | 1 (2.2) | 10 (4.8) | 1 (2.9) |
| Veterans Affairs | 61 (9.2) | 9 (6.6) | 13 (3.7) | 1 (2.2) | 8 (3.8) | 0 (-) |
| Indian Health Service | 48 (7.3) | 7 (5.3) | 8 (2.3) | 0 (-) | 6 (2.9) | 0 (-) |
| Other | 32 (6.3) | 2 (2.0) | 10 (3.7) | 1 (3.2) | 8 (4.9) | 0 (-) |
| Missing | 15 (1.9) | 2 (1.4) | 6 (1.5) | - | 4 (1.7) | - |
| COVID-19 severity‡ (mean (SD)) | 3.5 (1.4) | 3.6 (1.3) | 3.4 (1.2) | 3.3 (1.2) | 3.4 (1.2) | 3.3 (1.2) |
| Long COVID | 304 (38.1) | 46 (27.2) | 144 (35.3) | 13 (25.0) | 88 (38.1) | 10 (27.8) |
| Long COVID severity‡ (mean (SD)) | 4.2 (1.4) | 4.2 (1.4) | 4.0 (1.4) | 3.7 (1.1) | 4.0 (1.4) | 3.5 (1.2) |
| COVID-19 vaccine, ever | 632 (79.1) | 85 (57.4) | 329 (80.4) | 27 (56.3) | 188 (81.0) | 26 (72.2) |
| COVID-19 booster | 444 (55.6) | 44 (29.9) | 233 (57.0) | 14 (29.2) | 134 (57.8) | 15 (41.7) |

* Respondents could select >1 answer choice.
† Includes: Asian, American Indian, Alaskan Native, Native Hawaiian, Other Pacific Islander, none of these, prefer not to answer
‡ 6-point scale where 1 = no symptoms and 6 = very severe symptoms. Hospitalization and ICU care were defined as "very severe"

Table 12. Respondents who reported receiving care for their COVID illness, by setting, by analysis, n(%)

| | No exclusions | | Primary analysis | | Secondary analysis | |
|---|---|---|---|---|---|---|
| | **Adult sample (n=800)** | **Child sample (n=169)** | **Adult sample (n=409)** | **Child sample (n=52)** | **Adult sample (n=232)** | **Child sample (n=36)** |
| Primary care | 315 (39.4) | 97 (57.4) | 127 (31.1) | 38 (73.1) | 84 (36.2) | 26 (72.2) |
| Urgent care | 257 (32.1) | 54 (32.1) | 106 (25.9) | 19 (36.5) | 57 (24.6) | 16 (44.4) |
| Specialist visits | 106 (13.3) | 26 (15.5) | 25 (6.1) | 5 (9.6) | 16 (6.9) | 3 (8.3) |
| Emergency department visits | 175 (21.9) | 42 (24.9) | 63 (15.4) | 13 (25.0) | 34 (14.7) | 7 (19.4) |
| Hospitalization | 147 (18.4) | 32 (18.9) | 41 (10.0) | 8 (15.4) | 24 (10.3) | 6 (16.7) |
| Home health | 147 (18.4) | 32 (18.9) | 42 (10.3) | 9 (17.3) | 25 (10.8) | 7 (19.4) |
| Inpatient rehab | 100 (12.5) | 19 (11.5) | 24 (5.9) | 1 (2.0) | 14 (6.0) | 1 (2.9) |

Table 13. Visit number, time spent, and out of pocket costs by care setting, by analysis

a. No exclusions

| | Adult sample (n=800) | | | | | | Child sample(n=169) | | | | | |
|---|---|---|---|---|---|---|---|---|---|---|---|---|
| | N | Mean | SD | Median | Min | Max | N | Mean | SD | Median | Min | Max |
| **Primary care** | | | | | | | | | | | | |
| Number of visits | 800 | 6.4 | 101.2 | 0 | 0 | 2020 | 169 | 1.2 | 1.5 | 1 | 0 | 8 |
| Time spent (min) | 166 | 352.8 | 575.3 | 183.5 | 0 | 5800 | 97 | 149.1 | 405.0 | 70 | 0 | 3720 |
| OOP medical costs ($) | 222 | 78.7 | 300.0 | 10 | 0 | 3000 | 59 | 40.8 | 67.7 | 20 | 0 | 301 |
| Non-medical costs ($) | 226 | 67.6 | 380.8 | 10 | 0 | 4000 | 66 | 28.2 | 53.9 | 10 | 0 | 357 |
| **Urgent care** | | | | | | | | | | | | |
| Number of visits | 800 | 5.8 | 100.9 | 0 | 0 | 2020 | 168 | 0.5 | 1.0 | 0 | 0 | 5 |
| Time spent (min) | 171 | 416.0 | 591.4 | 270 | 0 | 6150 | 55 | 310.9 | 461.5 | 170 | 0 | 2704 |
| OOP medical costs ($) | 252 | 89.7 | 362.2 | 10 | 0 | 4300 | 39 | 39.7 | 63.4 | 20 | 0 | 250 |
| Non-medical costs ($) | 253 | 38.6 | 157.8 | 10 | 0 | 2300 | 54 | 16.7 | 28.3 | 10 | 0 | 150 |
| **Emergency department** | | | | | | | | | | | | |
| Number of visits | 797 | 3.1 | 71.6 | 0 | 0 | 2020 | 169 | 0.4 | 0.8 | 0 | 0 | 5 |
| Time spent (min) | 137 | 523.3 | 734.2 | 380 | 0 | 7590 | 42 | 573.5 | 870.1 | 275 | 70 | 4380 |
| OOP medical costs ($) | 169 | 130.2 | 416.0 | 10 | 0 | 3500 | 27 | 78.9 | 165.7 | 0 | 0 | 560 |
| Non-medical costs ($) | 171 | 47.7 | 208.2 | 9 | 0 | 2200 | 41 | 50.1 | 102.9 | 12 | 0 | 500 |
| **Hospitalization** | | | | | | | | | | | | |
| Number of visits | 798 | 5.9 | 101.2 | 0 | 0 | 2020 | 169 | 0.5 | 2.3 | 0 | 0 | 20 |
| Length of stay (d) | 131 | 4.9 | 8.2 | 4 | 0 | 90 | 31 | 3.9 | 4.4 | 2 | 1 | 20 |
| OOP medical costs ($) | 142 | 205.1 | 744.1 | 10 | 0 | 7800 | 19 | 162.8 | 352.7 | 33 | 0 | 1500 |
| Non-medical costs ($) | 143 | 96.5 | 362.7 | 10 | 0 | 3400 | 31 | 46.1 | 77.9 | 20 | 0 | 400 |

#### b. Primary analysis

| | Adult sample (n=409) | | | | | | Child sample (n=52) | | | | | |
|---|---|---|---|---|---|---|---|---|---|---|---|---|
| | N | Mean | SD | Median | Min | Max | N | Mean | SD | Median | Min | Max |
| **Primary care** | | | | | | | | | | | | |
| Number of visits | 409 | 0.7 | 1.4 | 0 | 0 | 10 | 52 | 1.2 | 1.1 | 1 | 0 | 4 |
| Time spent (min) | 58 | 181.6 | 171.5 | 97.5 | 0 | 667 | 38 | 96.4 | 54.2 | 90 | 0 | 210 |
| OOP medical costs ($) | 86 | 50.2 | 114.6 | 8.5 | 0 | 650 | 33 | 40.2 | 65.8 | 20 | 0 | 301 |
| Non-medical costs ($) | 88 | 21.0 | 43.6 | 7.5 | 0 | 300 | 35 | 16.4 | 23.1 | 10 | 0 | 100 |
| **Urgent care** | | | | | | | | | | | | |
| Number of visits | 409 | 0.5 | 1.1 | 0 | 0 | 10 | 52 | 0.5 | 0.8 | 0 | 0 | 4 |
| Time spent (min) | 54 | 218.4 | 168.4 | 135 | 40 | 726 | 19 | 136.0 | 79.6 | 120 | 40 | 320 |
| OOP medical costs ($) | 104 | 81.3 | 307.6 | 10 | 0 | 3000 | 16 | 54.7 | 75.8 | 22.5 | 0 | 200 |
| Non-medical costs ($) | 104 | 23.6 | 62.2 | 5 | 0 | 500 | 19 | 22.9 | 41.5 | 5 | 0 | 150 |
| **Emergency department** | | | | | | | | | | | | |
| Number of visits | 408 | 0.2 | 0.8 | 0 | 0 | 10 | 52 | 0.3 | 0.5 | 0 | 0 | 2 |
| Time spent (min) | 46 | 387.5 | 319.4 | 300 | 0 | 1590 | 13 | 248.7 | 120.6 | 240 | 95 | 563 |
| OOP medical costs ($) | 60 | 103.2 | 200.7 | 5 | 0 | 800 | 12 | 80.8 | 146.6 | 0 | 0 | 500 |
| Non-medical costs ($) | 61 | 23.2 | 45.8 | 5 | 0 | 200 | 13 | 34.4 | 67.1 | 12 | 0 | 245 |
| **Hospitalization** | | | | | | | | | | | | |
| Number of visits | 409 | 0.1 | 0.9 | 0 | 0 | 10 | 52 | 0.2 | 0.5 | 0 | 0 | 2 |
| Length of stay (d) | 40 | 3.7 | 2.4 | 3 | 0 | 14 | 8 | 2.3 | 1.8 | 1.5 | 1 | 5 |
| OOP medical costs ($) | 40 | 236.3 | 428.2 | 8 | 0 | 2000 | 8 | 132.4 | 179.0 | 75 | 0 | 500 |
| Non-medical costs ($) | 41 | 115.4 | 366.0 | 8 | 0 | 2000 | 8 | 41.9 | 52.4 | 27.5 | 0 | 150 |

### c. Secondary analysis

| | Adult sample (n=232) | | | | | | Child sample (n=36) | | | | | |
|---|---|---|---|---|---|---|---|---|---|---|---|---|
| | N | Mean | SD | Median | Min | Max | N | Mean | SD | Median | Min | Max |
| **Primary care** | | | | | | | | | | | | |
| Number of visits | 232 | 0.7 | 1.4 | 0 | 0 | 10 | 36 | 1.3 | 1.2 | 1 | 0 | 4 |
| Time spent (min) | 40 | 139.6 | 136.2 | 72 | 0 | 570 | 26 | 97.1 | 54.1 | 90 | 0 | 210 |
| OOP medical costs ($) | 57 | 47.0 | 118.2 | 10 | 0 | 650 | 24 | 40.5 | 74.6 | 12.5 | 0 | 301 |
| Non-medical costs ($) | 59 | 19.0 | 43.2 | 6 | 0 | 300 | 24 | 16.5 | 25.7 | 10 | 0 | 100 |
| **Urgent care** | | | | | | | | | | | | |
| Number of visits | 232 | 0.4 | 1.0 | 0 | 0 | 8 | 36 | 0.6 | 0.9 | 0 | 0 | 4 |
| Time spent (min) | 27 | 218.3 | 165.4 | 145 | 55 | 654 | 16 | 119.5 | 58.4 | 120 | 40 | 240 |
| OOP medical costs ($) | 56 | 100.0 | 405.1 | 10 | 0 | 3000 | 15 | 45.0 | 67.5 | 20 | 0 | 200 |
| Non-medical costs ($) | 57 | 18.8 | 45.5 | 0 | 0 | 260 | 16 | 17.2 | 30.2 | 2.5 | 0 | 100 |
| **Emergency department** | | | | | | | | | | | | |
| Number of visits | 231 | 0.2 | 0.7 | 0 | 0 | 5 | 36 | 0.2 | 0.5 | 0 | 0 | 2 |
| Time spent (min) | 22 | 420.3 | 321.7 | 323 | 0 | 1340 | 7 | 271.9 | 138.2 | 260 | 140 | 563 |
| OOP medical costs ($) | 31 | 71.7 | 150.5 | 5 | 0 | 560 | 7 | 124.3 | 181.1 | 70 | 0 | 500 |
| Non-medical costs ($) | 33 | 26.9 | 51.1 | 5 | 0 | 200 | 7 | 23.1 | 28.4 | 12 | 0 | 75 |
| **Hospitalization** | | | | | | | | | | | | |
| Number of visits | 232 | 0.2 | 0.8 | 0 | 0 | 5 | 36 | 0.2 | 0.5 | 0 | 0 | 2 |
| Length of stay (d) | 23 | 4.0 | 2.8 | 4 | 0 | 14 | 6 | 2.0 | 1.5 | 1.5 | 1 | 5 |
| OOP medical costs ($) | 23 | 182.8 | 320.3 | 7 | 0 | 1000 | 6 | 151.5 | 207.0 | 54.5 | 0 | 500 |
| Non-medical costs ($) | 24 | 123.0 | 404.1 | 7.5 | 0 | 2000 | 6 | 53.3 | 56.5 | 45 | 0 | 150 |

OOP = out of pocket; SD = standard deviation

### Table 14. Financial challenges, by analysis

| | No exclusions | | Primary analysis | | Secondary analysis | |
|---|---|---|---|---|---|---|
| | Adult sample (n=800) | Child sample (n=169) | Adult sample (n=409) | Child sample (n=52) | Adult sample (n=232) | Child sample (n=36) |
| Problems paying COVID-19-related medical bills | | | | | | |
| No | 655 (82.0) | 141 (83.4) | 360 (88.2) | 46 (88.5) | 204 (88.3) | 33 (91.7) |
| Yes, when I had COVID-19 | 91 (11.4) | 15 (8.9) | 29 (7.1) | 3 (5.8) | 19 (8.2) | 1 (2.8) |
| Yes, after I had COVID-19 | 35 (4.4) | 5 (3.0) | 14 (3.4) | 1 (1.9) | 6 (2.6) | 0 (-) |
| Yes, currently | 32 (4.0) | 10 (5.9) | 10 (2.5) | 2 (3.9) | 6 (2.6) | 2 (5.6) |
| Wait to get medical care due to cost | | | | | | |
| No | 631 (79.1) | 140 (82.8) | 341 (83.4) | 47 (90.4) | 196 (84.5) | 33 (91.7) |
| Yes, when I had COVID-19 | 96 (12.0) | 9 (5.3) | 41 (10.0) | 3 (5.8) | 21 (9.1) | 1 (2.8) |
| Yes, after I had COVID-19 | 51 (6.4) | 13 (7.7) | 16 (3.9) | 1 (1.9) | 7 (3.0) | 0 (-) |
| Yes, currently | 32 (4.0) | 11 (6.5) | 18 (4.4) | 3 (5.8) | 11 (4.7) | 2 (5.6) |
| Take less or skip doses of medicine due to cost | | | | | | |
| No | 660 (82.6) | 142 (84.0) | 365 (89.5) | 48 (92.3) | 209 (90.5) | 33 (91.7) |
| Yes, when I had COVID-19 | 72 (9.0) | 10 (5.9) | 22 (5.4) | 2 (3.9) | 11 (4.8) | 1 (2.8) |
| Yes, after I had COVID-19 | 47 (5.9) | 8 (4.7) | 15 (3.7) | 1 (1.9) | 10 (4.3) | 0 (-) |
| Yes, currently | 26 (3.3) | 11 (6.5) | 9 (2.2) | 3 (5.8) | 4 (1.7) | 2 (5.6) |
| Did not get medical care due to cost | | | | | | |
| No | 634 (79.3) | 135 (79.9) | 344 (84.1) | 45 (86.5) | 196 (84.5) | 32 (88.9) |
| Yes, when I had COVID-19 | 78 (9.8) | 10 (5.9) | 29 (7.1) | 1 (1.9) | 16 (6.9) | 1 (2.8) |
| Yes, after I had COVID-19 | 56 (7.0) | 10 (5.9) | 22 (5.4) | 2 (3.9) | 10 (4.3) | 0 (-) |
| Yes, currently | 39 (4.9) | 15 (8.9) | 18 (4.4) | 5 (9.6) | 11 (4.7) | 3 (8.3) |
| Did not get prescription medicine due to cost | | | | | | |
| No | 637 (79.6) | 139 (82.3) | 352 (86.1) | 46 (88.5) | 199 (85.8) | 33 (91.7) |
| Yes, when I had COVID-19 | 88 (11.0) | 10 (5.9) | 26 (6.4) | 2 (3.9) | 12 (5.2) | 1 (2.8) |
| Yes, after I had COVID-19 | 42 (5.3) | 10 (5.9) | 20 (4.9) | 0 (-) | 13 (5.6) | 0 (-) |
| Yes, currently | 42 (5.3) | 11 (6.5) | 16 (3.9) | 4 (7.7) | 11 (4.7) | 2 (5.6) |
| Problems paying non-medical bills | | | | | | |
| No | 595 (74.7) | 119 (70.4) | 329 (80.6) | 40 (76.9) | 186 (80.5) | 30 (83.3) |
| Yes, when I had COVID-19 | 90 (11.3) | 17 (10.1) | 33 (8.1) | 5 (9.6) | 20 (8.7) | 1 (2.8) |
| Yes, after I had COVID-19 | 63 (7.9) | 17 (10.1) | 21 (5.2) | 2 (3.9) | 11 (4.8) | 0 (-) |
| Yes, currently | 86 (10.8) | 24 (14.2) | 44 (10.8) | 8 (15.4) | 28 (12.1) | 5 (13.9) |
| Skip or limit meals due to cost | | | | | | |
| No | 608 (76.2) | 127 (75.2) | 335 (81.9) | 43 (82.7) | 194 (83.6) | 33 (91.7) |
| Yes, when I had COVID-19 | 84 (10.5) | 15 (8.9) | 31 (7.6) | 4 (7.7) | 14 (6.0) | 1 (2.8) |
| Yes, after I had COVID-19 | 60 (7.5) | 17 (10.1) | 21 (5.1) | 2 (3.9) | 13 (5.6) | 0 (-) |
| Yes, currently | 73 (9.2) | 17 (10.1) | 37 (9.1) | 5 (9.6) | 20 (8.6) | 2 (5.6) |
| Any financial problem | | | | | | |
| Yes, when I had COVID-19 | 218 (27.3) | 30 (17.8) | 86 (21.0) | 7 (13.5) | 47 (20.3) | 1 (2.8) |
| Yes, after I had COVID-19 | 153 (19.1) | 34 (20.1) | 66 (16.1) | 3 (5.8) | 37 (16.0) | 0 (-) |
| Yes, currently | 141 (17.6) | 36 (21.3) | 70 (17.1) | 10 (19.2) | 41 (17.7) | 6 (16.7) |
| Yes, at any time | 312 (39.0) | 64 (37.9) | 139 (34.0) | 15 (28.9) | 74 (31.9) | 7 (19.4) |

### Figure 1. Exclusion criteria flow chart

a. Adult sample

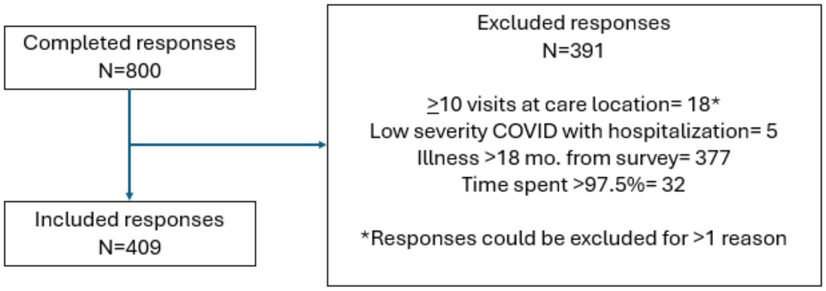

b. Child sample

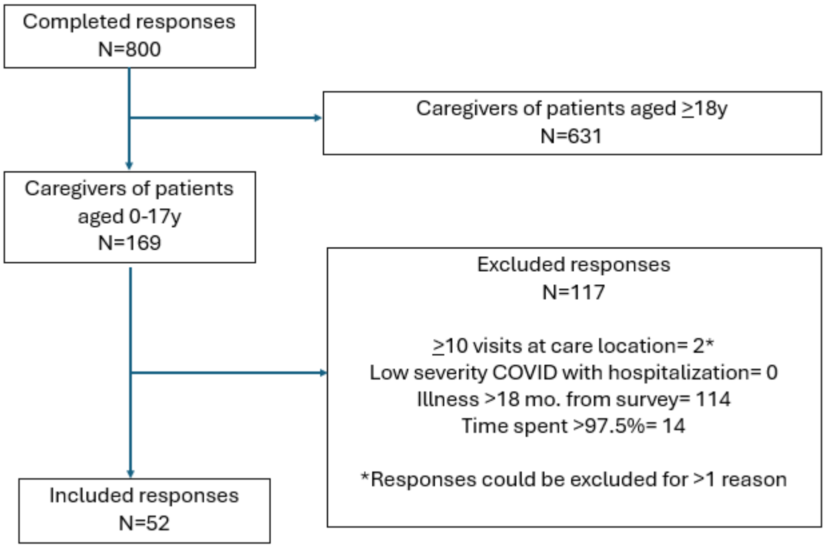
